# Rhinovirus-induced epithelial RIG-I inflammasome activation suppresses antiviral immunity and promotes inflammatory responses in virus-induced asthma exacerbations and COVID-19

**DOI:** 10.1101/2021.11.16.21266115

**Authors:** U Radzikowska, A Eljaszewicz, G Tan, N Stocker, A Heider, P Westermann, S Steiner, A Dreher, P Wawrzyniak, B Rückert, J Rodriguez-Coira, D Zhakparov, M Huang, B Jakiela, M Sanak, M Moniuszko, L O’Mahony, T Kebadze, DJ Jackson, MR Edwards, V Thiel, SL Johnston, CA Akdis, M Sokolowska

## Abstract

Rhinoviruses (RV) and inhaled allergens, such as house dust mite (HDM) are the major agents responsible for asthma onset, exacerbations and progression to the severe disease, but the mechanisms of these pathogenic reciprocal virus-allergen interactions are not well understood. To address this, we analyzed mechanisms of airway epithelial sensing and response to RV infection using controlled experimental in vivo RV infection in healthy controls and patients with asthma and in vitro models of HDM exposure and RV infection in primary airway epithelial cells. We found that intranasal RV infection in patients with asthma led to the highly augmented inflammasome-mediated lower airway inflammation detected in bronchial brushes, biopsies and bronchoalveolar lavage fluid. Mechanistically, RV infection in bronchial airway epithelium led to retinoic acid-inducible gene I (RIG-I), but not via NLR family pyrin domain containing 3 (NLRP3) inflammasome activation, which was highly augmented in patients with asthma, especially upon pre-exposure to HDM. This excessive activation of RIG-I inflammasomes was responsible for the impairment of antiviral type I/III interferons (IFN), prolonged viral clearance and unresolved inflammation in asthma in vivo and in vitro. Pre-exposure to HDM amplifies RV-induced epithelial injury in patients with asthma via enhancement of pro-IL1*β* expression and release, additional inhibition of type I/III IFNs and activation of auxiliary proinflammatory and pro-remodeling proteins. Finally, in order to determine whether RV-induced activation of RIG-I inflammasome may play a role in the susceptibility to severe acute respiratory syndrome coronavirus (SARS-CoV-2) infection in asthma, we analyzed the effects of HDM exposure and RV/SARS-CoV-2 coinfection. We found that prior infection with RV restricted SARS-CoV-2 replication, but co-infection augmented RIG-I inflammasome activation and epithelial inflammation in patients with asthma, especially in the presence of HDM. Timely inhibition of epithelial RIG-I inflammasome activation may lead to more efficient viral clearance and lower the burden of RV and SARS-CoV-2 infections.

## 1. Introduction

Asthma is one of the most common chronic inflammatory lung diseases affecting more than 5% of the global population^1^. Its pathogenesis and clinical presentation is complex, with a common feature of susceptibility to exacerbations leading to loss of disease control, hospitalizations, and in some cases, progressive loss of lung function^2^. Exacerbations of asthma are most often caused by common respiratory viruses^3, 4^, with rhinoviruses (RV) responsible for up to 80% of asthma attacks^3^. RVs that have been initially considered as benign viruses, now are also linked to the early-life development of asthma, severe bronchiolitis in infants and fatal pneumonia in elderly and immunocompromised patients^5, 6^. Likewise, human coronaviruses have not been strongly linked with asthma pathology^7^. However, the current pandemic of severe acute respiratory syndrome coronavirus 2 (SARS-CoV-2) has been challenging this view, resulting in contradictory observations of asthma being considered a risk factor for SARS-CoV-2 infection and coronavirus disease 2019 (COVID-19) severity^8–10^ or constituting a protection from the disease^11, 12^. Another important factor for asthma development and exacerbations is exposure to inhaled allergens. House dust mite (HDM) is the most significant source of perennial allergens worldwide. HDM sensitization is found in around 50%-85% of patients with asthma, and HDM exposure correlates with asthma severity^13^. There are strong epidemiological links between RV infections, allergen exposure and sensitization on the risk of asthma development and the rates of exacerbations^6, 14^. Children with early life RV-induced wheezing and aeroallergen sensitization have an extremely high incidence of asthma in later years^6^. Combination of virus detection in the airways with the high allergen exposure markedly increases the risk of hospital admission^15^. In line with this, HDM immunotherapy significantly reduces risk of asthma exacerbations^16^. It has been also recently suggested that allergen exposure might influence SARS-CoV-2 infection patterns in the general population^17, 18^. However, the underlying mechanisms of these noxious, reciprocal allergen-virus effects in asthma pathogenesis are incompletely understood.

The host response to the RV infection encompasses its RNA recognition by the endosomal toll-like receptor 3 (TLR) 3, TLR7/8 and cytoplasmic RNA helicases: retinoic acid-inducible gene I (RIG-I) and melanoma-differentiation-associated gene 5 (MDA5)^19^, whereas its capsid might interact with the cell surface TLR2 and initiate myeloid differentiation primary response 88 (MyD88)-dependent nuclear factor ‘kappa-light-chain-enhancer’ of activated B cells (NF-kB) activation^20^. RIG-I in its monomeric form binds to the 5’ end of viral RNA, undergoes conformational changes and interacts with mitochondrial antiviral signaling protein (MAVS)^21^. MAVS recruits tumor necrosis factor receptor-associated factor 3 (TRAF3) to activate TRAF family member-associated nuclear factor kappa B activator (TANK)-binding kinase (TBK)-1 and I*κ*B kinase *ε* (IKK*ε*) complex. TBK1 complex mediates phosphorylation of interferon (IFN) response factors and subsequent induction of type I and type III (I/III) IFNs^21^. Interferons further signal via their respective receptors which leads to the broad expression of interferon-stimulated genes (ISGs)^21^. Epithelial antiviral response is sufficient to clear RV infection in healthy airways^22^. In asthma, however, we and others demonstrated several alternations in RV-induced type I/III IFN responses^23–25^, but the pathomechanisms of those changes still remain elusive, suggesting greater complexity than previously anticipated, and potential involvement of additional factors such as allergens and other viruses adding to this complexity. SARS-CoV-2 is also sensed by RIG-I and MDA5, but due to several evasion mechanisms induction of IFNs by SARS-CoV-2 is reduced or delayed^26, 27^. There it still limited understanding of epithelial response to SARS-CoV-2 in asthma or in the presence of underlying allergic inflammation or other viral infection.

Other important host factors involved in sensing viruses, bacteria and other noxious agents are inflammasomes. Inflammasomes are supramolecular complexes, composed of a sensor protein, such as NLR Family Pyrin Domain Containing 3 (NLRP3), RIG-I, MDA5 and others, adaptor protein ASC, and caspase-1^28–30^. They are responsible for cleavage and release of the mature, active forms of IL-1*β* and IL-18, and induction of the proinflammatory cell death called pyroptosis^28^. Activation of RIG-I and NLRP3 inflammasomes has been demonstrated in macrophages and dendritic cells after infection with some respiratory RNA viruses, including RV ^31, 32^, influenza A (IAV) ^32–34^, SARS-CoV-1 ^35,36^and most recently SARS-CoV-2 ^37, 38^. However, activation of any epithelial inflammasomes by these viruses in vivo in human airways and their involvement in pathology of asthma remain poorly understood. It is not known whether they are necessary to clear infection or in contrast, whether they initiate mucosal hyperinflammation delaying virus clearance ^39, 40^, especially in the scenario when the same sensor protein, such as RIG-I or MDA5 can be involved in type I/III IFN response or in inflammasome activation. Likewise, an involvement of NLRP3 inflammasome in HDM-models of asthma and in severe asthma in humans has been demonstrated, however data are conflicting and remain poorly understood ^41–43^. Finally, airway epithelial response in health or during the preexisting allergic inflammation in asthma and combined infection with RV and SARS-CoV-2 is unknown.

Therefore, we analyzed mechanisms of airway epithelial sensing and response to RV infection using controlled experimental in vivo RV infection in healthy controls and patients with asthma and in vitro models of HDM exposure and RV/SARS-CoV-2 coinfection in primary airway epithelial cells from both groups. We found that RV infection in patients with asthma led to overactivation of RIG-I inflammasomes which diminished RIG-I accessibility for type I/III IFN responses, leading to their functional impairment, prolonged viral clearance and unresolved inflammation in vivo and in vitro. Pre-exposure to HDM augmented RIG-I inflammasome activation and additionally inhibited IFN-I/III responses. Prior infection with RV restricted SARS-CoV-2 replication, but co-infection augmented RIG-I inflammasome activation and epithelial inflammation in patients with asthma, especially in the presence of HDM.

## 2. Results

### 2.1 Intranasal infection with rhinovirus induced inflammasome-mediated immune responses in the epithelium of lower airways in asthma

First, we aimed to investigate mechanisms of airway epithelial sensing in bronchial epithelium in the rhinovirus (RV)-induced asthma exacerbations in vivo in humans in the controlled, experimental settings. Therefore, we analyzed bronchial brushings, bronchial biopsies and bronchoalveolar lavage fluid (BAL) from the controlled, experimental RV infection of patients with asthma and healthy controls. Samples were collected two weeks before (d-14, baseline) and 4 days after infection (Fig. 1A), as reported previously ^44^. Both groups were seronegative for anti-RV antibodies prior to infection and only individuals without recent natural respiratory infection underwent the experimental infection. An unbiased analysis of the gene ontologies and networks revealed significant differences in antigen presentation, interferon signaling and innate immune responses to RNA viral infection pathways in bronchial brushings from patients with asthma as compared to control individuals in response to RV infection (Fig. 1B). Importantly, we also noted a significant enrichment of genes in the inflammasome-mediated immune responses in asthma (Fig.1B, Supplementary Table S1). Many of these genes have been also included in the first ranked categories (Supplementary Table S1). Once investigated in detail, quite strikingly, we found a strong upregulation of transcriptome profiles of inflammasome-mediated immune responses in bronchial brushings from patients with asthma, in a sharp contrast to the downregulation or no change of similar genes in control individuals (Fig. 1C, D). We further validated the expression of IL-1*β* and caspase-1 proteins in bronchial biopsies of the same patients. We found higher expression of IL-1*β*in the epithelial areas of the bronchial biopsies of patients with asthma at baseline as compared to healthy controls (Fig. 1E, F). In line with its gene expression in bronchial brushings, we noted a decrease in IL-1*β* protein expression following RV infection in healthy controls, in contrast to the continuous upregulation of IL-1b in patients with asthma (Fig. 1E, F). Additionally, we assessed concentrations of mature IL-1*β* protein secreted into the BAL. In agreement with the epithelial mRNA and protein expression of IL-1*β*, we found significantly decreased IL-1*β* protein concentration in BAL fluid from control individuals 4 days after infection, whereas in patients with asthma IL-1*β* protein concentrations in BAL fluid tended to be increased, though this increase was not statistically significant (Fig. 1G, H). Likewise, we also noted downregulation of epithelial expression of caspase-1 in bronchial biopsies from healthy controls 4 days after infection, whereas it did not change in patients with asthma (Fig. 1I, J). Altogether, we demonstrated here that inflammasome is activated and there is an upregulation of inflammasome-mediated immune responses in bronchial epithelium of patients with asthma after in vivo infection with RV, whereas it is either being actively suppressed or already resolved 4 days after infection in healthy controls.

**Figure 1.**
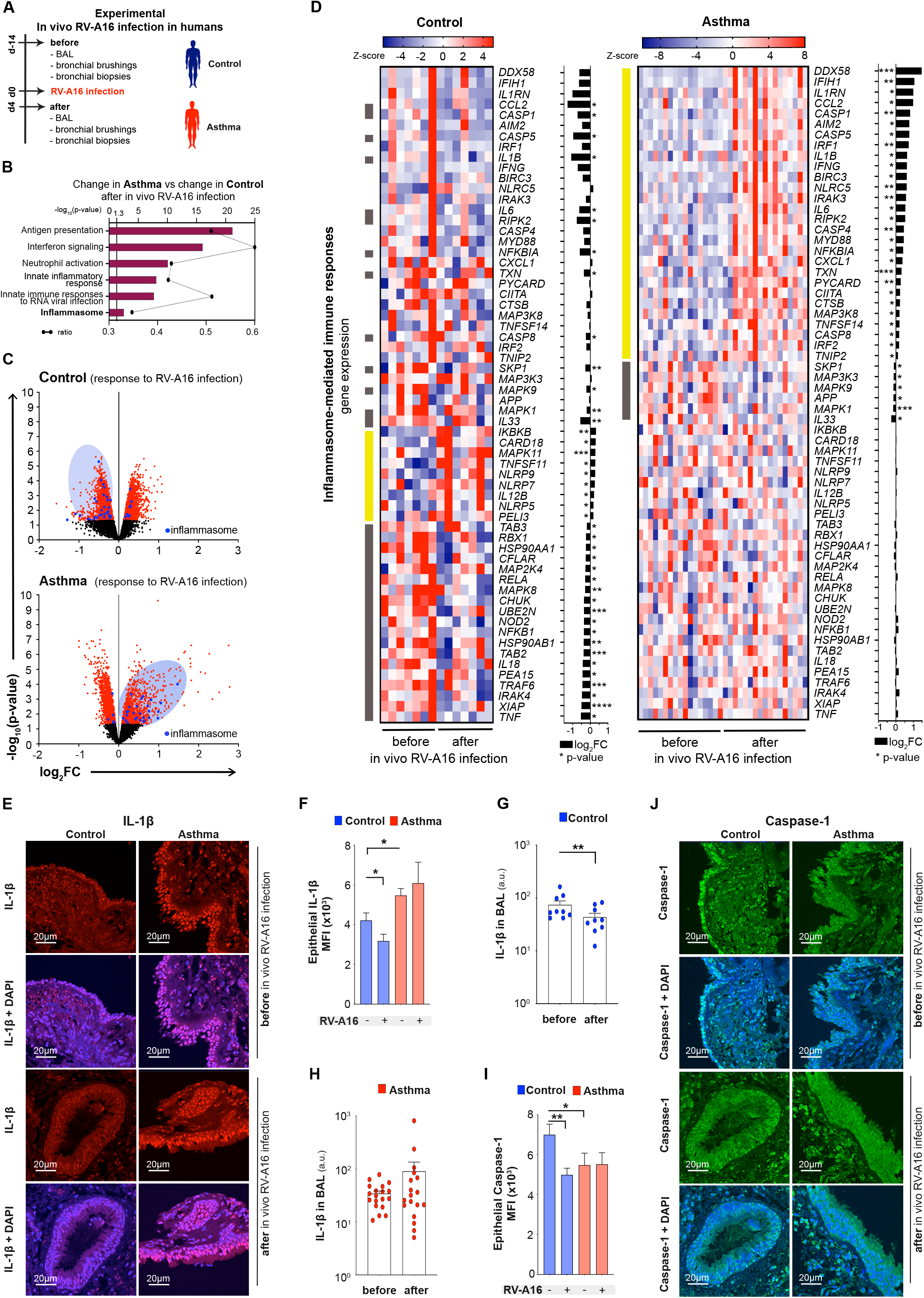
Intranasal infection with rhinovirus induced inflammasome-mediated immune responses in the epithelium of lower airways in asthma. **A)** Overview of the experimental RV-A16 infection in humans. Bronchoalveolar lavage (BAL) fluid, bronchial brushings, and bronchial biopsies from healthy controls and patients with asthma were collected two weeks before (day-14) and four days after (day 4) experimental infection with RV-A16 (100 TCID_50_) on day 0 in vivo. **B)** Top significantly enriched pathways within genes changed after RV-A16 infection in bronchial brushings from patients with asthma compared to genes changed after RV-A16 infection in control individuals in vivo (control n=7, asthma n=17). Black line represents a ratio of genes in the experiment over the whole pathway set. **C)** Volcano plots of all (black), significant (red), and significant inflammasome-mediated immune response (blue) genes in bronchial brushings from controls (upper panel) and patients with asthma (lower panel) after experimental in vivo RV-A16 infection (control n=7, asthma n=17). X axis presents log_2_ fold change (FC) of upregulated (right) and downregulated (left) genes. Y axis presents level of significance. **D)** Heatmap of genes encoding inflammasome-mediated immune responses after in vivo RV-A16 infection in controls (left panel) and patients with asthma (right panel) presented together with the corresponding log_2_ fold change (FC) expression changes (black bars) (control n=7, asthma n=17). Yellow and grey left side color bars represent genes upregulated and downregulated, respectively. **E-F**) Representative confocal images of IL-1β in bronchial biopsies at baseline and after in vivo RV-A16 infection, scale bars: 20μm. Quantification based on the mean fluorescence intensity (MFI) x10^3^ (10 equal epithelial areas from each biopsy of control subjects (n=3) and patients with asthma (n=3-4)). **G-H)** Secretion of IL-1β to BAL fluid before and after RV-A16 infection in vivo in **G)** control (n=9) and **H)** patients with asthma (n=18-19). Data are presented as arbitrary units (a.u.). **I-J)** Representative confocal images of caspase-1 in bronchial biopsies at baseline and after in vivo RV-A16 infection (control n=3, asthma n=3-4), scale bars: 20μm. Quantification based on the mean fluorescence intensity (MFI) x10^3^ (10 equal epithelial areas from each biopsy of control subjects (n=3) and patients with asthma (n=3-4)). (*) represents a significant difference between indicated conditions. Heatmap displays normalized gene expression across the groups (row normalization). Inflammasome-mediated immune responses gene set was curated based on databases and ontologies listed in the Molecular Signatures Database (MSigDB, Broad Institute, Cambridge). Bar graph data present mean ± SEM analyzed with one-way ANOVA (Kruskal-Wallis test), RM one-way ANOVA (Friedman test), mixed-effects model, or paired T-test or Wilcoxon test, as appropriate, depending on the data relation and distribution, p-value: *<0.05; **<0.005; ***<0.0005, ****<0.00005. *RV-A16*, rhinovirus A16; *BAL*, Bronchoalveolar lavage; *MFI*, mean fluorescence intensity*; a.u.*; arbitrary units; *TCID_50_*, median tissue culture infectious dose.

### 2.2 Augmented rhinovirus-induced RIG-I, but not MDA5 or NLRP3 inflammasome activation in bronchial epithelium in asthma

Having demonstrated rhinovirus-induced inflammasome activation in patients with asthma in vivo, we aimed to further characterize mechanisms, sensors and timelines of this phenomenon. First, we analyzed publicly available next-generation sequencing (NGS) data of RV-infected differentiated primary human bronchial epithelial cells (HBECs) from healthy controls and patients with asthma 24 h after infection ^45^. Confirming our in vivo results, an unbiased analysis of pathways and ontologies, revealed upregulated interferon signaling and innate immune responses to RNA viral infections (Fig. S1A, Supplementary Table S2). We also noted significant enrichment of inflammasome-mediated immune responses, in this early time point happening in control and asthma samples, yet still ranking slightly higher in patients with asthma (Fig. S1A). Accordingly, epithelium from both patients with asthma and control individuals showed increased inflammasome-mediated immune responses after RV infection (Fig. S1B, C), but the inflammasome-related molecules such as *CASP1* (caspase-1), *IL6*, NLR family CARD domain containing 5 (*NLRC5)*, *CXCL1* and others were significantly more upregulated in asthma (Fig. S1D). Next, we investigated mechanisms of the release of mature IL-1*β* in HBECs upon RV infection in a dose and time-dependent manner (Fig. 2A-C, S2A-C). RV infection at a multiplicity of infection (MOI) 0.1, but not UV-inactivated RV (UV-RV), induced secretion of mature IL-1*β* 24h after infection (Fig. 2A, B, S2B, C), which was significantly increased in patients with asthma (Fig 2B). Inflammasome activation was accompanied by a higher virus replication at this time point (Fig. S2D). We also demonstrated that mature IL-1*β* release was paired with formation of ASC specks in HBECs from control individuals and patients with asthma infected with RV (Fig. 2C). Again, complementary to IL-1*β* secretion, ASC specks count was higher in patients with asthma (Fig. 2D). We did not observe ASC-speck formation after UV-RV alone. Once we demonstrated that RV-induced inflammasome activation is augmented in patients with asthma, we also looked at the baseline status of IL-1*β* expression and the influence of infection on the inflammasome priming step in both groups. We observed higher expression of pro-IL-1*β* protein in HBECs from patients with asthma at baseline in vitro (Fig. 2A, S2E), in bronchial biopsies in vivo (Fig 1E, F), and a moderate upregulation of IL-1*β* concentrations in bronchoalveolar lavage (BAL) fluid in vivo (Fig. S2F). RV and UV-RV stimulation further increased expression of pro-IL-1*β* mRNA and protein, especially in epithelium in asthma (Fig. 2A, S2E, G). RV infection did not affect protein expression of ASC or pro-caspase-1 (Fig. 2A and S2H, I). Next, we noted significant inhibition of RV-induced inflammasome activation upon caspase-1 inhibitor (YVAD) treatment (Fig. 2E), while, as expected, it did not affect inflammasome priming (Fig. S2J). To investigate whether active RV infection is necessary for inflammasome activation in HBECs, we blocked RV entry to the cells using monoclonal antibodies, blocking its receptor ICAM-1. Indeed, significantly diminished RV infection (Fig. 2F) led to decreased mature IL-1*β* secretion (Fig. 2G). Using targeted proteomics, we further noted that in addition to IL-1*β*, also IL-18, IL-1*α*, tumor necrosis factor (TNF) and TNF-related activation-induced cytokine (TRANCE) were released 24h after RV infection in HBECs from patients with asthma (Fig. S2K) indicating, that RV-induced epithelial inflammasome activation participated in the heightened proinflammatory responses at the bronchial barrier sites in asthma. Finally, we investigated which of the pattern recognition receptors (PRRs) expressed in human bronchial epithelium acts as a sensor and activator of inflammasome assembly. Based on the abundant expression after RV infection in vivo and in vitro (Fig. 1D, S1B, C), and their capability to form inflammasomes in response to other RNA viruses in hematopoietic cells ^30, 31^ or in epithelium ^32^, we focused on RIG-I (*DDX58*) and MDA5 (*IFIH1*) receptors. *DDX58* (RIG-I) and *IFIH1* (MDA5) mRNA was expressed in HBECs from both controls and patients with asthma at baseline (Fig. S3A, B). We observed further increases in *DDX58* (RIG-I) (Fig. S3C, D) and *IFIH1* (MDA5) mRNA (Fig. S3E, F) expression in HBECs upon RV infection. Interestingly, RV-induced upregulation of *DDX58* (RIG-I) was more enhanced in asthma as compared to control (Fig. S3D). Additionally, we observed increased expression of RIG-I protein (Fig. 2H, S3G, H) upon RV infection, accompanied by formation of RIG-I speck-like structures (Fig. 2I). Indeed, coprecipitation of ASC with RIG-I confirmed RIG-I binding to ASC upon RV infection (Fig. 2J). Notably, MDA5 was not bound to ASC upon RV infection (Fig. 2K). Lastly, taking into account various reports regarding NLRP3 inflammasome activation in airway epithelium upon viral infections^31, 32, 46, 47^, we also assessed its importance in human bronchial epithelium. We found extremely low expression of *NLRP3* mRNA in fully differentiated, mature bronchial epithelial cells from patients with asthma and control individuals at baseline (Fig. S3I) or after RV infection (Fig. S3J). In line with that, we did not detect the expression of NLRP3 protein in differentiated HBECs from controls and patients with asthma at baseline or after RV infection (Fig. 2L, M, S3K, L). Finally, a specific NLRP3 inflammasome inhibitor (MCC950) did not affect the secretion of mature IL-1*β*upon RV infection (Fig. 2N). In summary, we demonstrated here that RV infection led to activation of RIG-I inflammasome in the differentiated primary human bronchial epithelial cells, which was augmented in patients with asthma. NLRP3 and MDA5 inflammasomes were not activated by RV infection.

**Figure 2.**
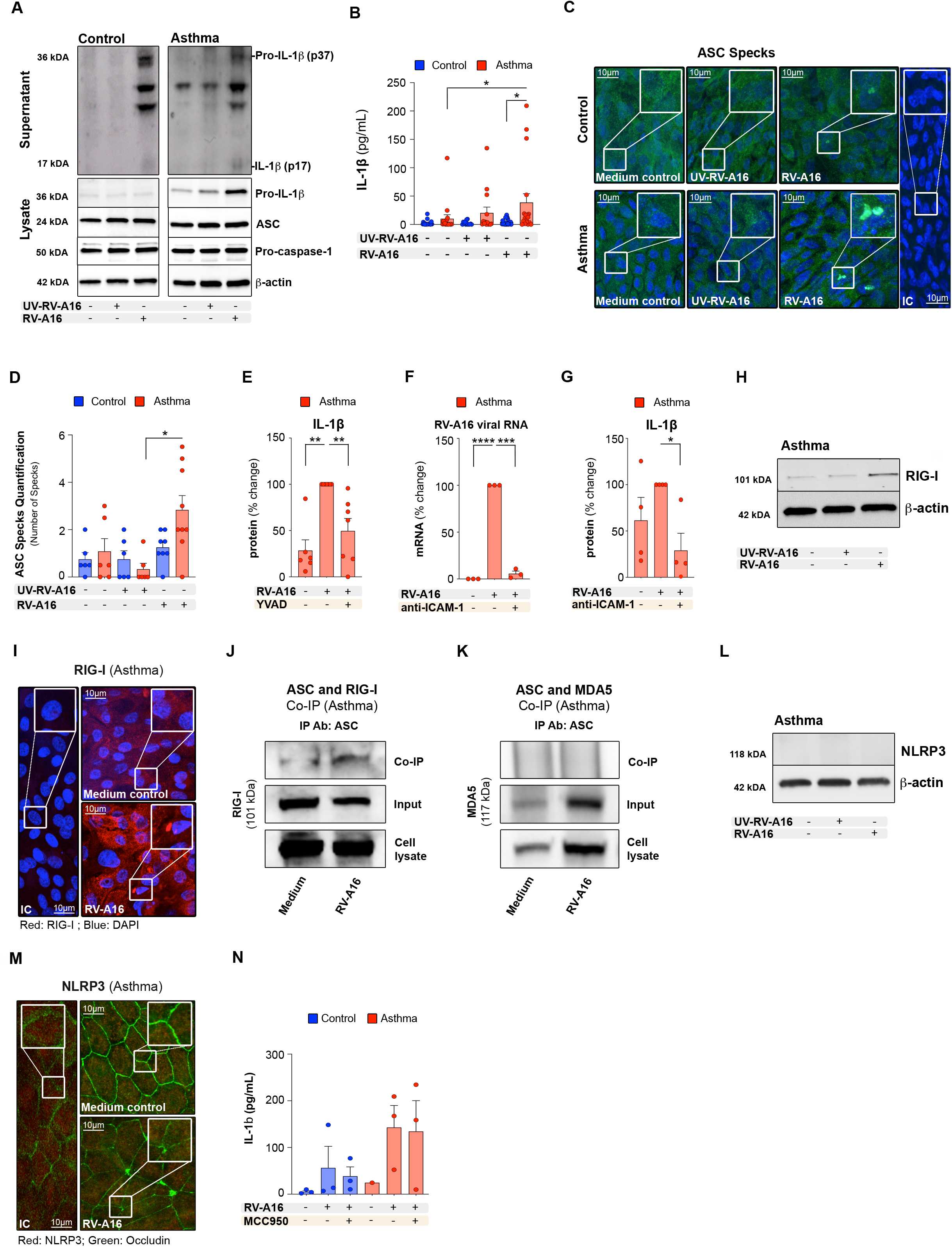
Augmented rhinovirus-induced RIG-I, but not MDA5 or NLRP3 inflammasome activation in bronchial epithelium in asthma. **A)** Representative Western Blot images of secreted IL-1β (apical compartment), and pro-IL-1β, ASC, pro-caspase-1 and β-actin (cell lysates) from HBECs from control subjects (left panel) and patients with asthma (right panel). **B)** IL-1β release to the apical compartment assessed by ELISA (control n=14-22; asthma n=14-17). **C)** Representative confocal images of ASC speck formation in HBECs from control individuals and patients with asthma (control n=3, asthma n=3); scale bars: 10μm. **D)** Quantification of ASC specks, presented as a number of specks (5-11 equal epithelial areas from control n=3, asthma n=3). **E)** IL-1β release to the apical compartment assessed by ELISA (n=6-7) in HBECs from patients with asthma in the presence or absence of caspase-1 inhibitor (YVAD). Data are presented as the percentage of the response after HDM+RV-A16 treatment. **F)** Expression of *RV-A16 positive strand* (RV-A16 viral RNA) in the HBECs from patients with asthma after anti-ICAM-1 stimulation (n=3) was assessed using RT-PCR, and presented as a relative quantification (RQ=2^-ΔΔCt^) as compared to the vehicle condition from patients with asthma. Data are presented as the percentage of the response after HDM+RV-A16 treatment. **G)** IL-1β release to the apical supernatants assessed by ELISA (n=4) in HBECs (n=3) from patients with asthma in the presence or absence of anti-ICAM-1 combined with HDM+RV-A16 stimulation. Data are presented as the percentage of the response after HDM+RV-A16 treatment. **H)** Representative Western Blot images of RIG-I protein expression in HBECs from patients with asthma (n=4). **I)** Representative confocal images of RIG-I in HBECs (n=3) from patients with asthma; scale bars: 10μm. **J)** Co-immunoprecipitation (co-IP) of ASC/RIG-I complex using anti-ASC antibodies followed by RIG-I detection in the presence of HDM (n=4). **K)** Co-immunoprecipitation (co-IP) of ASC/MDA5 complex using anti-ASC antibodies followed by MDA5 detection. **L)** Representative Western Blot images of NLRP3 protein in the HBECs from patients with asthma (n=4). **M)** Representative confocal images of NLRP3 and Occludin in HBECs from patients with asthma in the presence of HDM (n=3), scale bars: 10μm. **N)** IL-1β release to the apical compartment with/without RV-A16 and NLRP3 inflammasome inhibitor (MCC950) (control n=3, asthma n=3). HBECs from patients with asthma are presented in red, HBECs from control individuals are presented in blue. (*) represents a significant difference as indicated. Bar graph data show mean ± SEM analyzed with one-way ANOVA (Kruskal-Wallis test), RM one-way ANOVA (Friedman test) or mixed-effects model, as appropriate, depending on the data relation (paired or unpaired) and distribution, *p-value≤0.05, **p-value≤0.01, ***p-value≤0.001. *ALI*; Air-liquid interface cultures; *anti-ICAM-1*, anti-ICAM-1 antibody; *IC*; Isotype control; *HDM*, house dust mite; *RV-A16*, rhinovirus A16; *UV-RV-A16*, UV-treated rhinovirus A16; *YVAD*, ac-YVAD-cmk (caspase-1 inhibitor); *MOI*, multiplicity of infection; *NPX*, normalized protein expression; *MCC950*, NLRP3 inflammasome inhibitor; *Co-IP*, Co-immunoprecipitation; *IP Ab*, antibodies used for co-precipitation.

### 2.3 Activation of the RIG-I inflammasome impaired RIG-I dependent interferon signaling in bronchial epithelium of patients with asthma

Since the major function of RIG-I is recognition of RNA viruses ^21^, we also analyzed the status of antiviral genes and proteins involved in in vivo responses to RV infection. In line with inflammasome-mediated immune responses, the majority of genes encoding antiviral pathways were still upregulated 4 days after in vivo RV infection in patients with asthma while they were either downregulated or not changed in healthy controls (Fig. 3A, B, Supplementary Table S3). These data suggest less effective resolution of RV infection and delayed clearance of the virus in asthma. Indeed, RV load in the bronchoalveolar lavage fluid in asthma was around 100-fold higher than in controls and the peak nasal lavage virus load was 25-fold higher in patients with asthma than in healthy controls, though this difference was not statistically significant (Fig. 3C, D). Thus, we showed here that bronchial epithelium from healthy individuals can efficiently respond to RV infection which leads to rapid virus clearance and subsequent resolution of antiviral responses. In contrast, in asthma, the lack of resolution of antiviral responses and delayed virus clearance suggest that there is an ongoing process in epithelium, which impairs the effectiveness of antiviral mechanisms. Therefore, we hypothesized that excessive RIG-I inflammasome activation and subsequent IL-1*β*secretion in response to RV infection in asthma, resulted in persistent, but less efficient RIG-I-mediated anti-RV response. Hence, we further studied whether RIG-I activation of MAVS/TBK1/IKK*ε* and downstream interferon signaling inhibits RIG-I inflammasome activation and conversely if formation of RIG-I inflammasome inhibits interferon signaling. First, we used BX795, a chemical inhibitor of TBK1 and IKK*ε*. As expected, it blocked expression of *IFNL2/3* (IFN-*λ*) in HBECs of patients with asthma (Fig. 3E) and subsequently decreased expression of *DDX58* (RIG-I) (Fig. 3F). BX795 treatment also reduced expression of interferon-stimulated chemokines: CXCL10, CXCL11 and CCL3 (Fig. 3G). It also led to a trend to increased RV infection (Fig. 3H), as well as to significantly augmented inflammasome priming (Fig. 3I) and activation (Fig. 3J). Next, we blocked IL-1*β* processing by RIG-I inflammasome with the use of the caspase-1 inhibitor YVAD and investigated interferon signaling and RV infection. Inhibition of RIG-I inflammasome activation and subsequent IL-1*β* signaling indeed led to trends of increasing expression of *IFNB* (IFN-*β*) (Fig. 3K) and *DDX58* (RIG-I) mRNA (Fig. 3L) and it further increased the production of CXCL10, CXCL11, CCL3, and CCL4 (Fig. 3M). Since ASC specs and assembled inflammasome complexes are often released from the cells together with mature IL-1*β* ^48^, we analyzed RIG-I protein expression also in the supernatants of the cells. Accordingly, we found that activation of RIG-I inflammasome led to increased release of RIG-I protein from the epithelial cells (Fig. 3N), which may translate to the decreased expression of RIG-I protein assessed in bronchial biopsies in vivo (Fig. 3O). In summary, these data suggest that increased RV-dependent RIG-I inflammasome activation in bronchial epithelium disturbed the effectiveness of RIG-I dependent anti-RV responses in asthma. Therefore, a timely blockade of the excessive RIG-I inflammasome activation and IL-1*β* signaling may lead to more efficient viral clearance and lower burden of infection.

**Figure 3.**
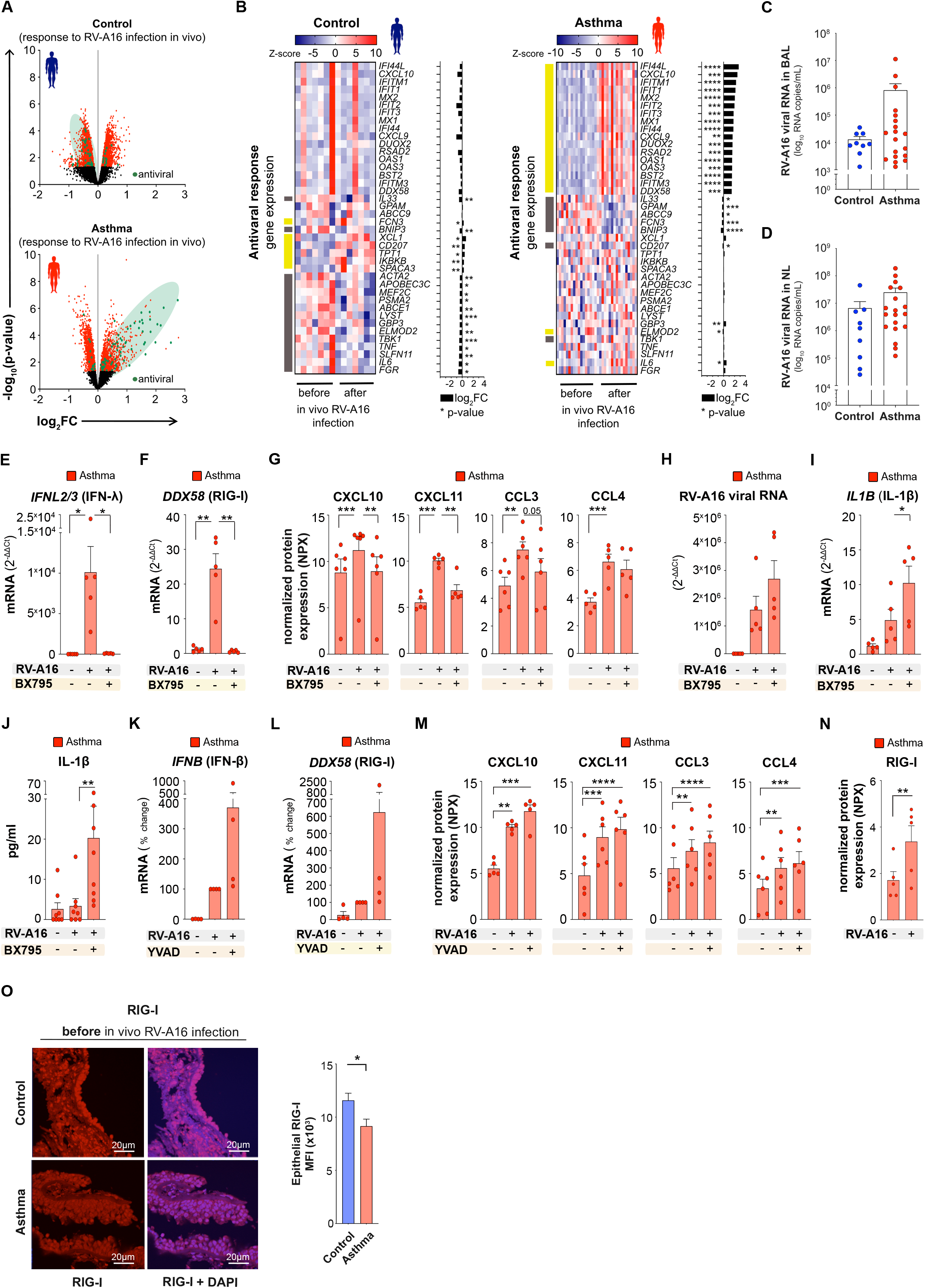
Activation of the RIG-I inflammasome impaired RIG-I dependent interferon signaling in bronchial epithelium of patients with asthma. **A)** Volcano plots of all (black), significant (red), and significant antiviral (green) genes in bronchial brushings from controls (upper panel) and patients with asthma (lower panel) after experimental in vivo RV-A16 infection (control n=7, asthma n=17). X axis presents log_2_ fold change (FC) of upregulated (right) and downregulated (left) genes. Y axis presents level of significance. **B**) Heatmap of antiviral genes significantly changed four days after experimental RV-A16 infection in healthy controls (left panel) and/or in patients with asthma (right panel) presented together with the log_2_ fold change (FC) (black bars) (control n=7, asthma n=17). Yellow and grey left side color bars represent genes upregulated and downregulated, respectively. **C-D)** RV-A16 virus load in **C)** the bronchoalveolar lavage (BAL) fluid and **D)** the nasal lavage (NL) fluid in control individuals and patients with asthma four days after experimental RV-A16 infection (control n=9, asthma n=19). Data presented as log_10_ viral RNA copies per 1 mL of BAL/NL. **E-J)** HBECs from patients with asthma were infected with RV-A16 in the presence or absence of BX795, a chemical inhibitor of TBK1 and IKKε, or vehicle. mRNA expression of **E)** *IFNL2/3* (IFNλ) and **F)** *DDX58* (RIG-I) assessed using RT-PCR and presented as relative quantification (RQ=2^-ΔΔCt^) as compared to the vehicle condition (n=5). **G)** Secretion of CXCL10, CXCL11, CCL3, and CCL4 proteins into the apical compartment assessed with the PEA targeted proteomics (n=6). Data are presented as normalized protein expression (NPX). Expression of **H)** *RV-A16 positive strand* (RV-A16 viral RNA) and **I)** *IL1B* (IL-1β) assessed using RT-PCR and presented as relative quantification (RQ=2^-ΔΔCt^) (n=5). **J)** IL-1β release to the apical compartment assessed by ELISA (n=8). **K-N)** HBECs from patients with asthma were infected with RV-A16 in the presence or absence of YVAD, a caspase-1 inhibitor or vehicle. mRNA expression of **K)** *IFNB* (IFNβ) and **L)** *DDX58* (RIG-I) (n=4). Data are demonstrated as the percentage of the expression normalized to the RV-A16 condition. **M)** Secretion of CXCL10, CXCL11, CCL3, and CCL4 into apical compartment assessed with the PEA proteomics (n=6). Data are presented as normalized protein expression (NPX). **N)** RIG-I release to the apical compartment assessed by the Proximity Extension Assay proteomics (PEA) in HBECs from patients with asthma (n=5). Data are presented as normalized protein expression (NPX). **O**) Representative confocal images of RIG-I expression in bronchial biopsies at baseline and after experimental in vivo RV-A16 infection, scale bars: 20μm. Quantification based on the mean fluorescence intensity (MFI) x10^3^ (10 equal epithelial areas from each biopsy of control subjects (n=3) and patients with asthma (n=3-4)). (*) represents a significant difference between indicated conditions. Graph data present mean ± SEM analyzed with one-way ANOVA (Kruskal-Wallis test), RM one-way ANOVA (Friedman test) or mixed-effects model, as appropriate, depending on the data relation and distribution, p-value: *<0.05; **<0.005; ***<0.0005, ****<0.00005. Antiviral response gene set was curated based on databases and ontologies listed in the Molecular Signatures Database (MSigDB, Broad Institute, Cambridge). *HBECs*, differentiated human bronchial epithelial cells; *RV-A16*, rhinovirus A16; *MFI*; mean fluorescent intensity*; NPX*, normalized protein expression; *YVAD*, ac-YVAD-cmk (caspase-1 inhibitor); *BX795,* TBK1/IKKε inhibitor; *BAL*, bronchoalveolar lavage; *NL*, nasal lavage.

### 2.4 House dust mite enhanced rhinovirus-induced inflammasome activation in bronchial epithelium in asthma

Knowing that house dust mite (HDM) exposure combined with rhinovirus infection have an especially detrimental impact on severity of asthma exacerbations in children and adults^6, 15^, we investigated the effect of HDM on rhinovirus-induced RIG-I inflammasome activation in bronchial epithelium in a dose, time and lot-dependent manner (Fig. S4A-C). HDM stimulation combined with RV infection, significantly increased mature IL-1*β* secretion in bronchial epithelium of controls and patients with asthma, yet this effect was much more pronounced in asthma (Fig. 4A, B). HDM alone did not lead to release of inflammasome-processed mature IL-1*β*, but it induced the abundant release of non-mature pro-IL-1*β*in both groups (Fig. 4A). HDM pre-exposure followed by RV infection further increased expression of pro-IL-1*β* protein, especially in epithelium in asthma (Fig. 4A, C, S4D), suggesting combined effects of HDM and RV-replication independent and dependent mechanisms on pro-IL-1*β* expression. Expression of ASC and pro-caspase-1 proteins was stable and comparable between controls and patients with asthma (Fig. 4A, S4E, F). HDM prestimulation also increased RV-induced ASC specks formation in HBECs from controls and patients with asthma (Fig. 4D, E), confirming HDM involvement in the enhancement of inflammasome activation. In line with the lack of the release of processed IL-1*β*, we did not observe formation of ASC-specks after HDM stimulation alone (Fig. 4D, E). Additionally, HDM pre-exposure increased RV-induced release of RIG-I from the epithelial cells of patients with asthma (Fig. 4F). In summary, we demonstrated here, that while HDM alone did not activate any epithelial inflammasome in human bronchial epithelium, it led to the release of pro-IL-1*β*, as well as HDM pre-exposure increased RV-induced RIG-I inflammasome activation in bronchial epithelium, which was augmented in patients with asthma.

**Figure 4.**
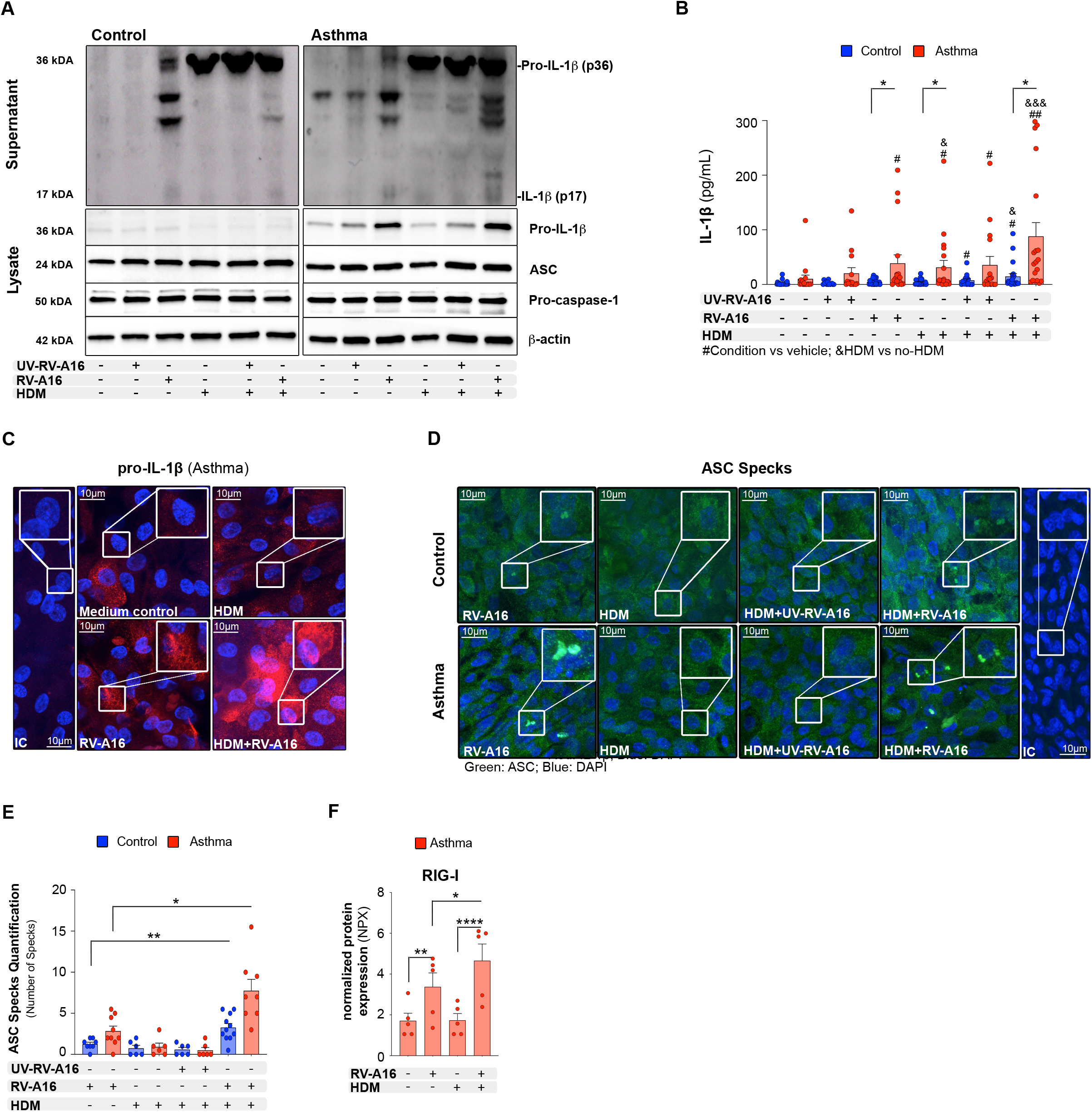
House dust mite enhanced rhinovirus-induced inflammasome activation in bronchial epithelium in asthma. **A)** Representative Western Blot images of secreted IL-1β (apical compartment), and pro-IL-1β, ASC, pro-caspase-1 and β-actin (cell lysates) from HBECs from control subjects (left panel) and patients with asthma (right panel). **B)** IL-1β release to the apical compartment assessed by ELISA (control n=14-22; asthma n=14-17). **C)** Representative confocal images of IL-1β in HBECs from patients with asthma (n=3); scale bars: 10μm. **D)** Representative confocal images of ASC speck formation in HBECs from control individuals and patients with asthma (control n=3, control n=3); scale bars: 10μm. **E)** Quantification of ASC specks, presented as a number of specks (5-11 equal epithelial areas from control n=3, asthma n=3). **F)** RIG-I release to the apical compartment assessed by the Proximity Extension Assay proteomics (PEA) in HBECs from patients with asthma (n=5). Data are presented as normalized protein expression (NPX). HBECs from patients with asthma are presented in red, HBECs from control individuals are presented in blue. (*) represents a significant difference as indicated. (#) represents a significant difference of the designated condition as compared to the vehicle from the same group. (&) represents a significant difference upon HDM treatment as compared to the corresponding condition without HDM. Bar graph data show mean ± SEM analyzed with one-way ANOVA (Kruskal-Wallis test), RM one-way ANOVA (Friedman test) or mixed-effects model, as appropriate, depending on the data relation (paired or unpaired) and distribution, *p-value≤0.05, **p-value≤0.01, ***p-value≤0.001. *ALI*; Air-liquid interface cultures; *anti-ICAM-1*, anti-ICAM-1 antibody; *IC*; Isotype control; *HDM*, house dust mite; *RV-A16*, rhinovirus A16; *UV-RV-A16*, UV-treated rhinovirus A16; *YVAD*, ac-YVAD-cmk (caspase-1 inhibitor); *MOI*, multiplicity of infection; *NPX*, normalized protein expression.

### 2.5 House dust mite impaired interferon responses in rhinovirus-infected bronchial epithelium of patients with asthma

Having demonstrated that HDM increased RV-induced RIG-I inflammasome activation, we continued to explore the effect of HDM pre-exposure on the timing and strength of antiviral responses. We found that HDM pre-treatment decreased RV-induced mRNA expression of *IFNB* (IFN-*β*) and *DDX58* (RIG-I) only in HBECs from patients with asthma after RV infection in vitro (Fig. 5A, S5A, B). Notably, HDM stimulation did not influence RV infection (Fig. S5C). When we analyzed protein expression and enriched biological pathways by targeted proteomics in the same conditions, we observed decreased cell- and IFNs-mediated antiviral responses in HBECs from control individuals and patients with asthma at 24h post infection (Fig. 5B). In addition, HDM in the presence of RV stimulated the release of epithelial to mesenchymal transition factors such as interleukin 15 receptor subunit alpha (IL-15RA) ^49, 50^, artemin (ARTN) ^51^, tolerance inducing TNF receptor superfamily member 9 (TNFRSF9) ^52^ (also called 4-1BB and CD137), extracellular newly identified RAGE-binding protein (EN-RAGE) alarmin ^53^ in both studied groups. However, only in asthma, complementary with all our previous data, HDM simultaneously increased the activation status and release of proinflammatory and pro-remodeling proteins, such as IL-1*α*, signaling lymphocytic activation molecule family member 1 (SLAMF1) ^54^, cluster of differentiation (CD) 40 ^55, 56^ and TRANCE (RANKL) ^57, 58^ (Fig. 5B, Supplementary Table S4). Importantly, HDM pre-stimulation had a slightly additive effect to the antiviral RIG-I pathway inhibitor (BX795) and further reduced BX795-decreased protein expression of the ISGs: CXCL10, CXCL11, CCL3 and CCL4 (Fig. 5C). It all suggests that pre-exposure to HDM, before RV infection decreases IFN type I response and this way it further contributes to the enhanced inflammasome-mediated impairment of antiviral responses in patients with asthma.

**Figure 5.**
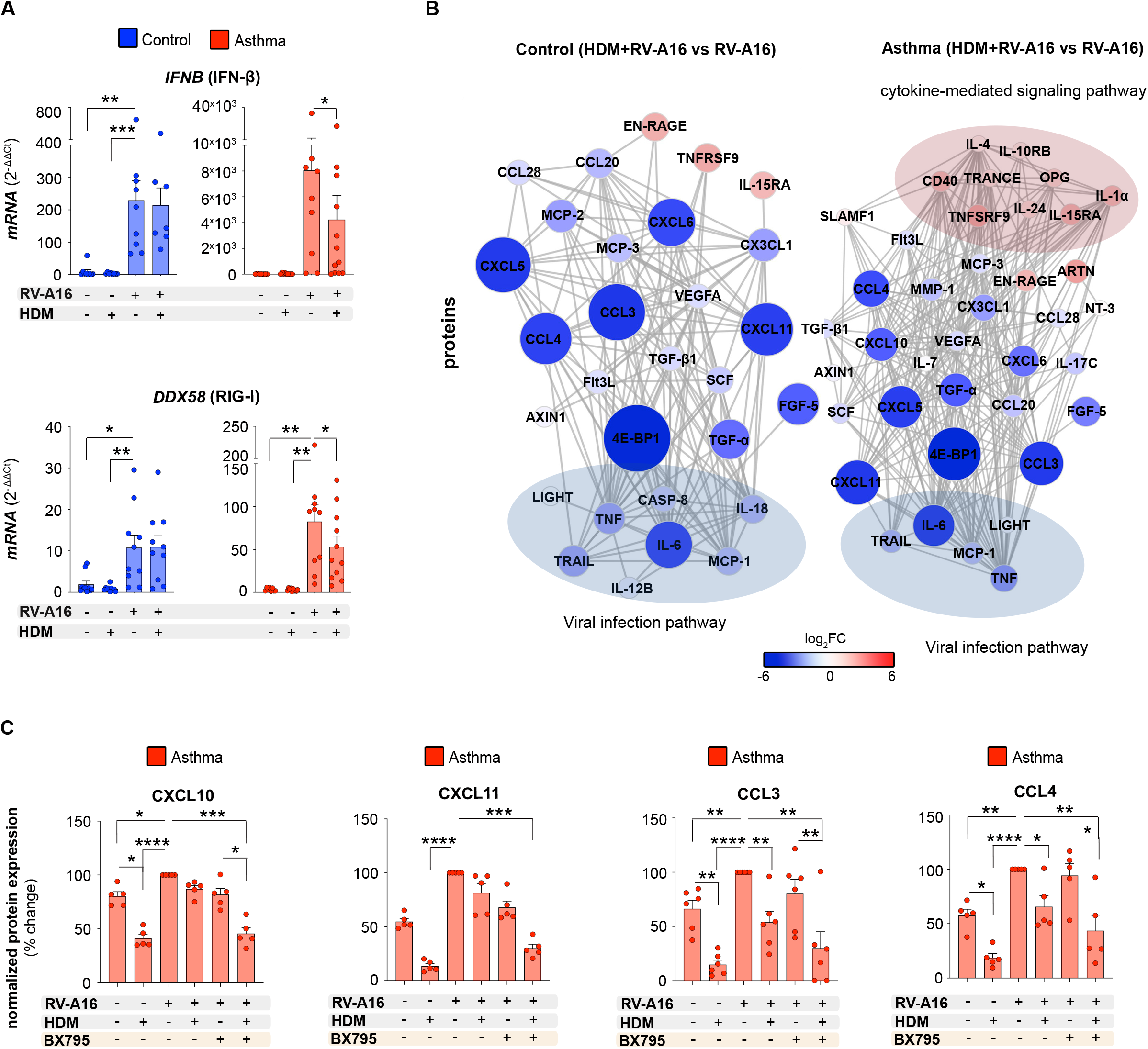
House dust mite impaired interferon responses in rhinovirus-infected bronchial epithelium of patients with asthma. mRNA expression of **A)** *IFNB* (IFN-β) (upper panel), and *DDX58* (RIG-I) (lower panel) (control n=7-9, asthma n=7-12) **B)** Visualization of interaction network of significant proteins secreted to the apical compartment from HBECs from control individuals (left panel) and patients with asthma (right panel) after in vitro treatment with HDM+RV-A16, when compared to RV-A16 infection alone assessed with PEA targeted proteomics (control n=5, asthma n=8). Network nodes represent log_2_FC of significantly upregulated (red), and downregulated (blue) proteins; proteins not interacting with each other are not shown. Edges represent protein-protein interactions. Proteins enriched in viral infection or cytokine-mediated signaling pathway are marked with blue and red eclipses, respectively. Prepared with STRING (https://string-db.org) and Cytoscape (https://cytoscape.org). **C)** Expression of CXCL10, CXCL11, CCL3, and CCL4 in the apical compartment of HBECs from patients with asthma pre-treated with HDM or vehicle, followed by the infection with RV-A16 in the presence of BX795 or vehicle assessed with PEA proteomics (n=6). Data presented as the percentage of the response to the RV-A16 condition. (*) represents a significant difference between indicated conditions. Graph data present mean ± SEM analyzed with one-way ANOVA (Kruskal-Wallis test), RM one-way ANOVA (Friedman test) or mixed-effects model, as appropriate, depending on the data relation and distribution, *p-value≤0.05, **p-value≤0.01, ***p-value≤0.001, ****p-value≤0.0001. *HBECs*, differentiated human bronchial epithelial cells; *RV-A16*, rhinovirus A16; *p.i.*, post-infection; *HDM,* house dust mite; *BX795*, TBK1/IKKε inhibitor; *NPX*, normalized protein expression.

### 2.6 Pre-existing rhinovirus infection attenuated SARS-CoV-2 infection, but augmented epithelial inflammation in asthma

Finally, facing the current pandemic and noting the contradictory results about asthma as a risk factor for COVID-19 in different populations ^8–12^, we investigated if RV-induced RIG-I inflammasome activation and HDM-mediated decrease of IFN responses may affect SARS-CoV-2 infection. We first treated primary HBECs from healthy controls and patients with asthma with or without HDM, next after 24h we infected them with RV, and after a further 24h we infected them with SARS-CoV-2 for 48h (Fig. 6A). We confirmed infection with SARS-CoV-2 by the detection of its nucleocapsid protein N (Fig. 6B) and the increase of SARS-CoV-2 viral RNA (Fig. 6C). In patients with asthma, but not in healthy controls, we observed lower infection with SARS-CoV-2 in samples pre-infected with RV (Fig.6C). Individual samples with high RV virus loads (Fig. 6D) had lower SARS-CoV-2 infection, and vice versa. This effect was diminished upon HDM pre-stimulation in samples from patients with asthma, suggesting that HDM pre-stimulation tended to increase SARS-CoV-2 infection in RV+SARS-CoV-2 infected epithelium in asthma (Fig. 6C). To understand these different patterns of infection, we analyzed the expression of *DDX58* (RIG-I), type I/III IFNs and *IL1B* (IL-1*β*). In line with our previous data, RV infection increased the expression of *DDX58* (RIG-I) (Fig. 6E, S6A), *IFNB* (IFN-*β*) (Fig. 6F, S6B)*, IFNL1* (IFN-*λ*1) (Fig. 6G, S6C) and *IFIH1* (MDA5) (Fig. 6H, S6D). However, we noted that infection with SARS-CoV-2 alone did not induce expression of *DDX58* (RIG-I)*, IFIH1* (MDA5)*, IFNB* (IFN-*β*)*, IFNL1* (IFN-*λ*1) (Fig. 6E-H, S6A-D) or secretion of proinflammatory proteins (Fig. 6I-J, Supplementary Table S5) at this timepoint. In contrast, HDM prestimulation increased the release of IL-18 in SARS-CoV-2-infected HBECs from patients with asthma (Fig. 6I-J). Importantly though, we observed that infection with SARS-CoV-2 on top of the infection with RV resulted in the tendency to increase IL-18 (Fig. 6I-J, Supplementary Table S5) and *IL1B* (IL-1*β*) (Fig. 6K, S6E). Notably, HDM pre-stimulation resulted in an amplified secretion of proinflammatory proteins, such as TRAIL, IL-15, CXCL9, IL-17C and CCL8 only in epithelium of patients with asthma infected with both viruses, whereas these conditions induced IL-18 secretion both in patients with asthma and control individuals (Fig. 6I-J, Supplementary Table S5). In summary, we found here that pre-existing RV infection restricted SARS-CoV-2 replication in asthma, but not in controls, which was in line with the sustained type I/III IFNs at this time-point, induced by RV in asthma. SARS-CoV-2 infection alone did not induce *DDX58* (RIG-I)*, IFIH1* (MDA5)*, IFNB* (IFN-*β*)*, IFNL1* (IFN-*λ*1) at 48h after infection. However, especially in the presence of HDM, we observed enhanced proinflammatory responses in patients with asthma after SARS-CoV-2 co-infection with RV, in spite of the reduced viral load of SARS-CoV-2.

**Figure 6.**
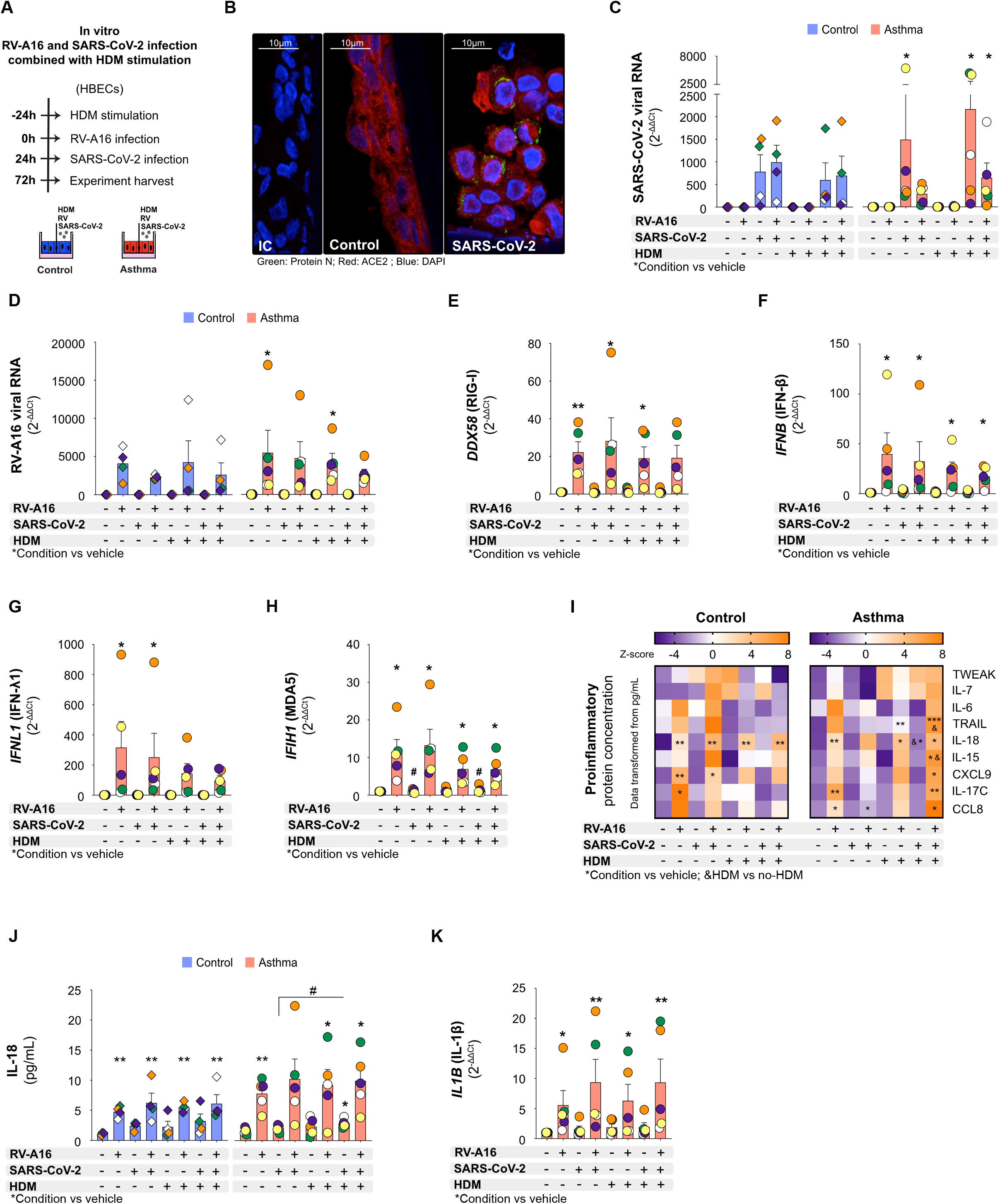
Pre-existing rhinovirus infection attenuated SARS-CoV-2 infection, but augmented epithelial inflammation in asthma. **A)** Overview of the experimental RV-A16 and severe acute respiratory syndrome coronavirus 2 (SARS-CoV-2) co-infection with/without HDM pre-treatment. Primary human bronchial epithelial cells (HBECs) from control subjects (n=4) and patients with asthma (n=5), differentiated in the Air-liquid Interface (ALI) conditions, were treated with house dust mite (HDM) (200 μg/mL) or vehicle for 24h, infected with rhinovirus A16 (RV-A16) in the multiplicity of infection (MOI) 0.1 for 24h, and then co-infected with SARS-CoV-2 in the MOI 0.1 for the following 48h. **B)** Representative confocal images of SARS-CoV-2 N protein and ACE2 in HBECs from patients with asthma after SARS-CoV-2 infection; scale bars: 10μm. Expression of **C)** SARS-CoV-2 virus load (average expression of *N protein*, *S protein* and *ORF1AB*) **D)** *RV-A16 positive strand* (RV-A16 viral RNA), **E)** *DDX58* (RIG-I), **F)** *IFNB* (IFN-β), **G)** *IFNL1* (IFN-λ1) **H)** *IFIH1* (MDA5) was assessed using RT-PCR and presented as relative quantification (RQ=2^-ΔΔCt^) compared to medium condition separately for HBECs from controls and patients with asthma. **I)** Heatmap of proinflammatory proteins assessed in the apical compartments of HBECs from controls (left panel) and patients with asthma (right panel) after in vitro HDM pre-stimulation and RV-A16 and SARS-CoV-2 co-infection analyzed with the quantitative Proximity Extension Assay (PEA) targeted proteomics. Data are transformed from the concentrations in pg/mL. **J)** IL-18 secreted to the apical compartment analyzed with the quantitative PEA. Data are presented as pg/mL. **K)** mRNA expression of *IL1B* (IL-1β) in HBECs from patients with asthma. Patients with asthma are presented in transparent red, control individuals in transparent blue. (*) represents a significant difference as compared to the vehicle from the same group. (&) represents a significant difference between HDM-pre-treated and HDM-naïve condition. (#) represents significant difference as indicated. Bars depict the mean ± SEM, whereas color-coded diamonds (control subjects) or circles (patients with asthma) show individual data from the same donor. Data are present as mean analyzed with one-way ANOVA (Kruskal-Wallis test), RM one-way ANOVA (Friedman test) or mixed-effects model, as appropriate, depending on the data relation and distribution, *p-value≤0.05, **p-value≤0.01, ***p-value≤0.001. *RV-A16*, rhinovirus A16; *HDM*, House Dust Mite; *HBECs*, Human Bronchial Epithelial Cells; *SARS-CoV-2*, Severe Acute Respiratory Syndrome Coronavirus 2; *ALI*, air-liquid interface; *MOI*, multiplicity of infection.

## 3. Discussion

In the current study we demonstrated in vivo and in vitro that recognition of replicating RV by RIG-I helicase in bronchial epithelial cells of patients with asthma led to the augmented ASC recruitment, oligomerization, activation of caspase-1, processing and release of mature IL-1*β* via formation of RIG-I inflammasome, independently of NLRP3 or MDA5. This excessive activation of RIG-I inflammasome by RV in patients with asthma compromised RIG-I-dependent type I/III IFNs and ISG responses, leading to less effective virus clearance and to unresolved airway inflammation. In addition, we found that pre-exposure to HDM amplified RV-induced epithelial injury in patients with asthma via enhancement of pro-IL1*β* expression and release, additional inhibition of type I/III IFNs and activation of auxiliary pro-inflammatory and pro-remodeling proteins. Finally, we showed that prior infection with RV restricted SARS-CoV-2 replication, but co-infection augmented RIG-I inflammasome activation and epithelial inflammation in patients with asthma, especially in the presence of HDM.

There are four main inflammasomes described to date to be involved in innate antiviral immunity against RNA viruses – the NLRP3, RIG-I and in some cases MDA5 and absent in melanoma 2 (AIM2) inflammasomes ^30, 34, 59, 60^. They are activated by several stimuli involved in the viral infection, such as viral nucleic acids, viroporins, RNA-modulating proteins, reactive oxygen species (ROS) and others^40^. Here, we found, that in vivo in humans and in fully differentiated primary human bronchial epithelium, infection with RV, a single stranded RNA virus, leads to increased priming of pro-IL-1*β* in a replication independent and dependent manner, and to assembly of RIG-I/ASC inflammasome, in a replication dependent manner. We did not see involvement of NLRP3 inflammasome upon RV infection or HDM exposure or even significant expression of NLRP3 in airway epithelium at baseline in any of our in vivo, in vitro or data mining approaches. We also did not see MDA5 forming inflammasome. Other groups observed that infection of human peripheral blood mononuclear cells, human macrophages and mouse bone-marrow derived cells with other single stranded RNA viruses, vesicular stomatitis virus (VSV) and IAV, activates RIG-I/MAVS-dependent pro-IL-1*β* transcription and RIG-I/ASC-dependent, but NLRP3-independent, inflammasome activation and mature IL-1*β* and IL-18 production^30, 46^. In contrast, in undifferentiated, submerged cultures of primary human airway epithelial cells infection with IAV revealed RIG-I and NLRP3-inflammasome-dependent mature IL-1*β* release^32^, while infection with RV in a similar model led to activation of NLRP3/NLRC5/ASC complexes and mature IL-1*β* release^47^. These differences might come from i) the undifferentiated state of the cells in the previous studies, as in non-differentiated epithelium, lacking ciliated cells, viral infection might engage different pathways than in human airways in vivo^61, 62^; ii) from different expression of inflammasome components in undifferentiated and differentiated mature epithelium lining human airways, as we showed previously^63^; as well as iii) from the differences in the virus strains and serotypes^64^. Importantly, our in vitro and in vivo data consistently showed the same results that RIG-I is engaged as inflammasome in bronchial epithelium upon RV infection, which constitutes an important early time-point event, triggering subsequent airway inflammation. It is also possible, that both inflammasomes are engaged in different cellular compartments: RIG-I/ASC in airway epithelium and NLRP3 in the infiltrating inflammatory cells in the airways. Indeed, RV infection in mice leads to partly macrophage-derived, NLRP3 inflammasome-dependent airway inflammation^31^. However, neither depletion of macrophages nor NLRP3 knockout leads to the complete blockade of mature caspase-1 and IL-1*β*processing upon RV infection, underlining that RV induces other inflammasomes in airway bronchial epithelium^31^.

An appropriate balance between activation of RIG-I epithelial inflammasome with RIG-I-dependent type I/III IFN responses lead to the limitation of viral replication, efficient virus clearance and timely resolution of airway inflammation in case of IAV infection^32^. Likewise, we observed here that in the bronchial epithelium of healthy subjects at early time points during RV infection there was an activation of RIG-I inflammasome and inflammasome-mediated immune responses, together with efficient type I/III IFN and ISG-responses. Importantly all of these responses were actively inhibited or went back to the pre-infection state, already 4 days after in vivo infection. In contrast, in epithelium of patients with asthma, there was enhanced RIG-I inflammasome activation starting early after infection and still non-resolved in vivo 4 days after infection. Overactivation of epithelial RIG-I inflammasome and subsequent increases in mature IL-1*β* release were at least partially responsible for the impairment of type I/III IFN/ISG responses. We demonstrated this here by blocking caspase-1 with YVAD which led to an increase in IFN-*β (IFNB)* and RIG-I (*DDX58*) together with IFN-responsive chemokines such as CXCL10, CXCL11, CCL3, and CCL4. Our findings are in line with early observations showing that IL-1β is able to attenuate transcription and translation of type I IFNs and excessive IFNα/β-induced effects^65, 66^. Interestingly, we also noted here that this cross-talk between IL-1*β* and type I IFNs is reciprocal, as blocking phosphorylation of TBK1 and IKK*ε* by their inhibitor BX795 and thus reducing RIG-I-induced type I interferons, significantly increased pro-IL-1*β* transcription and its processing by RIG-I inflammasome in airway epithelium. With this finding, we added and important point to the long-lasting discussion about the underlying origins and mechanisms of the frequent viral infections and exacerbations in patients with asthma^67, 68^.

Presence of other airway barrier-damaging and/or activating factors, such as exposure to allergens in addition to the viral infection, worsens the clinical outcomes, leads to a more severe exacerbation, hospitalization or respiratory failure^7^. Mechanistically, it might be connected with the multiplication of activated pathways and/or with the additive effects of different triggers on the same pathway. HDM activates airway epithelium via, among others, TLR2/4, C-type lectins, and protease-activated receptors (PARs) in an allergen-non-specific way to initiate allergen sensitization, but also to perpetuate already developed allergic and probably non-allergic airway inflammation in the absence of the sufficiently developed inhibitory signals^42, 69, 70^. It was recently observed in the clinical trials that one of the strongest effects of HDM-specific immunotherapy in patients with asthma is the reduction of the rate of asthma exacerbations^16^. However, it is not well understood if and how HDM-induced signaling in the airway epithelium interferes with the effectiveness of early antiviral response. Here, we found that HDM contributes significantly to RV-induced pathologic responses in human airway epithelium in patients with asthma by i) enhancement of non-mature pro-IL-1*β* expression and release, ii) overactivation of RIG-I inflammasome and subsequent release of mature caspase-1, mature IL-1*β*, and RIG-I, iii) inhibition of type I/III IFNs and ISG-responses and iv) activation of extra proinflammatory and pro-remodeling proteins, such as IL-1*α*, SLAMF1^54^, CD40^55^ and TRANCE (RANKL)^58^. In the presence of both HDM and RV, we noted that increased usage of RIG-I protein engaged in inflammasome formation and its subsequent expulsion is paired with HDM-dependent inhibition of RIG-I, type I/III IFNs and several ISGs. At an early stage of RV infection, it might explain disturbance of the antiviral response dynamics observed in asthma by us and others^71^. Due to this functional reduction of RIG-I availability, the following type I/III IFN-response is less effective, not able to quickly and efficiently clear the infection and thus it is sustained up to later time points, together with the enhanced inflammasome-related proinflammatory response.

Co-infections with two or more respiratory viruses occur often and likely acts as additional factors increasing airway epithelial damage. Patients with asthma are at a greater risk of developing respiratory failure as shown in the case of H1N1 influenza infection^72^. Thus, it has been somewhat surprising that so far during the current SARS-CoV-2 pandemic, epidemiological cohorts of COVID-19 patients from different geographical locations have resulted in partially contradictory observations that asthma is (USA, United Kingdom, Australia) or is not (Europe, China) a risk factor for SARS-CoV-2 infection and/or severity of COVID-19^11, 73, 74^. We and others demonstrated that the expression of angiotensin-converting enzyme 2 (ACE2), the main SARS-CoV-2 receptor and other plausible points of entry, are not changed in patients with asthma, even though different types of airway inflammation or inhaled steroids might modulate their expression and as such it seems unlikely that it would be the main reason for observed discrepancies^75^. Different geographical locations might represent variable levels and quality of environmental exposures such as viruses or allergens, which may interfere with the rate of SARS-CoV-2 infections and COVID-19 severity^18^. SARS-CoV-2, an enveloped, positive-sense, single-stranded (ss)RNA virus has been shown to be sensed, depending on the cell type, by MDA5^76^, RIG-I^76^ and NLRP3^77, 78^. However, due to several evasion properties and encoding by non-structural (Nsp) and accessory proteins, such as Nsp1,6,12,13, various open read frames (ORFs), protein M, protein N and others, which antagonize interferon pathways on many levels, induction of IFNs by SARS-CoV-2 itself is reduced or delayed^26, 27, 79^ with the augmented proinflammatory mediator release. Several in vitro and in vivo animal studies revealed that SARS-CoV-2, similarly to SARS-CoV-1, is very sensitive to pre-treatment with type I/III IFNs^80–82^, which inhibit its replication. Patients with disrupted IFN gene expression and production or patients with autoantibodies against type I IFNs have been shown to be at a greater risk of severe COVID-19. In accordance with these studies, we observed here that infection of epithelium from patients with asthma at the moment of heightened, but functionally impaired IFNs type I/III response induced by RV, results in reduced SARS-CoV-2 viral load, in contrast to the healthy epithelium, where the RV-induced IFN-I/III response has been already actively resolved. However, even if restricted, SARS-CoV-2 infection in combination with RV and HDM led to an increase in activation of epithelial inflammasome and release of higher amounts of IL-18 and other proinflammatory cytokines. In context of timing and possible clinical relevance it may mean that patients with asthma with pre-existing RV-infection might have a slightly restricted SARS-CoV-2 infection at first, but due to excessive inflammasome-related damage and proinflammatory signaling, together with SARS-CoV-2-induced inhibition of type I/III IFNs, they may in fact succumb eventually to more severe COVID-19. Importantly, in the presence of HDM, these potentially adverse effects are further heightened, meaning that HDM reduces RV-induced IFN response, which leads to higher SARS-CoV-2 replication and it enhances RIG-I inflammasome activation, inflammation and tissue damage.

All in all, we showed here in vivo and in vitro that the lack of balance between activation of RIG-I inflammasome and the RIG-I-IFNs-axis in response to a common respiratory viruses is an important driving factor of epithelial damage, lack of viral clearance and sustained airway inflammation in patients with asthma (Fig. S7). Timely targeting of this abnormal response by the yet-to-be developed early therapeutics or even prophylactic approaches might provide in the future a beneficial strategy to prevent RV-induced exacerbations of asthma and potentially severe COVID-19.

## 4. Material and Methods

### Reagents

House dust mite (HDM) extract (HDM) (Allergopharma, Reinbek, Germany) and house dust mite extract B (HDM B) (Citeq, York, UK) were diluted in sterile 0.9% saline (NaCl) and stored in −20°C. The concentration of the HDM used for the experiments was calculated according to the total protein content. Detailed description of the HDM extracts including protein, main allergens and endotoxin content is presented in the Supplementary Table S6.

All commercially available antibodies and reagents used in the manuscript are described in the Supplementary Table S7.

Mouse IgG2a monoclonal anti-human ICAM-1 antibody (antibody R6.5) was produced from the hybridoma cells (ATCC HB-9580, mouse hybridoma). The hybridoma cells were expanded in RPMI 1640 medium supplemented with 10% (v/v) IgG depleted fetal bovine serum, 2 mM L-glutamine, 1.25 g/L D-(+)-Glucose, 1 mM Sodium Pyruvate, 10 mM HEPES, 100 U/mL Penicillin-Streptomycin at 37°C in 5% CO_2_. The IgG in culture media were affinity purified from the cell culture supernatant using a 1 mL HiTrap™ Protein G HP column (GE Healthcare, 29-0485-81). Eluted fractions were immediately neutralized and buffer exchanged into PBS using dialysis. The antibody was then filtered through a 0.22 µm filter and stored at 4°C.

### Viruses

Rhinovirus A16 (RV-A16) was purchased from Virapur (San Diego, USA). UV-light inactivated RV-A16 (UV-RV-A16) was used as a control after an exposure to UV-light of the 254 nm at a 2 cm distance for 60 min. Cells were infected with RV-A16 or UV-RV-A16 at the multiplicity of infection (MOI) 0.1, 0.01 and 0.001 as determined by plaque assay. Briefly, HeLa cells were infected with the virus serial dilutions from 10^-2^ to 10^-8^ in duplicates. Seven days after infection, cells were fixed with formaldehyde solution and stained with 1% crystal violet in 20% ethanol and dH_2_0. Visible plaques were counted under a microscope.

SARS-CoV-2 viral genome was generated from the synthetic DNA fragments produced by GenScript (Piscataway, USA) using the in-yeast transformation-associated recombination (TAR) cloning method, as previously described^83^. In-vitro transcription was performed for the linearized Yeast artificial chromosome (YAC), containing the cDNA of the SARS-CoV-2 genome, as well as a PCR amplified SARS-CoV-2 N gene using theT7 RiboMAX Large Scale RNA production system (Promega, Madison, USA) with m7G(5ʹ)ppp(5ʹ)G cap provided as described previously^84^. Transcribed, capped mRNA was subsequently electroporated into baby hamster kidney cells (BHK-21) expressing SARS-CoV N protein. Co-culture of electroporated BHK-21 cells with susceptible Vero E6 cells produced passage 0 of SARS-CoV-2 virus. Passage 0 was used to infect Vero E6 cells to generate passage 1 working stocks, which were used for all experiments. Titers were determined using standard plaque assay, as described previously^83^.

### Patients groups and experimental in vivo RV-A16 infection in humans

Experimental in vivo rhinovirus infection in 11 control individuals and 28 patients with asthma was performed as reported previously^44^. Briefly, non-smoking, non-atopic control individuals, and non-smoking mild/moderate patients with asthma without any recent viral illness and without serum neutralizing antibodies towards RV-A16, who passed inclusion criteria, underwent infection on day 0 with RV-A16 at the dose of 100 TCID_50._ Bronchial brushings, bronchial biopsies and bronchoalveolar lavage (BAL) fluid were collected around 2 weeks before and at 4 days after RV-A16 infection. Additionally, nasal lavage (NL) samples at the peak of RV-A16 infection were collected to assess RV-A16 infection rates. Only subjects who had sufficient remaining samples to be analyzed in this study and/or subjects who had successful infection in the lungs, as assessed by viral RNA copies by qPCR, were included in the BAL, NL, and biopsies analyses (n=9 healthy control, n=19 patients with asthma), and bronchial brushing microarray analysis (n=7 healthy controls, n=17 patients with asthma). The study received ethical approval from the St. Mary’s Hospital Research Ethics Committee (09/H0712/59). All participants gave written, informed consent. The clinical characteristics of the 9 control and 19 asthma study participants who had sufficient remaining samples to be analyzed in this study is presented in Supplementary Table S8.

Control individuals and patients with asthma were enrolled in the ALL-MED Medical Research Institute, Wroclaw, Poland; the Pulmonary Division, University Hospital of Zurich, Switzerland (cohort SIBRO)^85^, or at the University Hospital, Jagiellonian University Medical College, Cracow, Poland (cohort A)^86^, as described previously. Briefly, bronchoscopy with epithelial cells brushings and BAL fluid collection was performed. The study was granted ethical permission from Switzerland and Poland (KEK-ZH-Nr. 20212-0043 – Kantonale Ethik-Kommission Zürich; KB-70/2013 and KB-567/2014 – Bioethical Committee, Wroclaw Medical University) or the Jagiellonian University Bioethics Committee (KBET/68/B/2008 and KBET/209/B/2011). Asthma diagnosis and severity were assessed according to the GINA guidelines^2^. All participants gave written, informed consent. Clinical characteristics of the study participants is presented in the Supplementary Table S8.

Primary Human Bronchial Epithelial cells (HBECs) were obtained from the above-listed cohorts or from the doctor-diagnosed asthma and control individuals from two independent commercial sources: Lonza (Basel, Switzerland), and Epithelix (Plan-les-Ouates, Switzerland). Characteristics of the HBECs used in the manuscript are presented in Supplementary Table S9.

### Air-liquid interface (ALI) cultures of bronchial epithelium from control individuals and patients with asthma

HBECs from the control subjects and patients with asthma were cultured and differentiated in the air-liquid interface (ALI) conditions as described previously, with minor alterations of the previous protocol^86^. Briefly, cells from passage 2 were grown in 20 mL of bronchial epithelial basal medium (Lonza, Basel, Switzerland) supplemented with the SingleQuot Kit (Lonza, Basel, Switzerland) placed in 150cm^2^ T-flask in humidified incubator at 37°C with 5% CO_2_ for maximum 10 days, or until 80%-90% confluency. Next, cells were trypsinized (ThermoFisher Scientific, Waltham, USA) and seeded at a density of 1.5×10^5^ cells/well on the 6.5-mm-diameter polyester membranes with the 0.4 μm pore size and growth area of 0.33 cm^2^ (Costar, Corning, NY, USA; Oxyphen, Wetzikon, Switzerland) in 24-well culture plates. Bronchial Epithelial Growth Medium (BEGM) (Lonza, Basel, Switzerland) supplemented with the SingleQuot kit (Lonza, Basel, Switzerland), with an exception of the retinoic acid (ATRA, Sigma-Aldrich, St. Louis, USA) and triiodothyronine, was mixed in the 1:1 ratio with the Dulbecco modified Eagle medium (DMEM, Gibco, Thermofisher Scientific, Waltham, USA). Fresh all-trans ATRA (Sigma-Aldrich, Merck, Kenilworth, USA) was supplemented at a concentration of 15 ng/mL. Cells were grown submerged for 3-5 days in the apical medium and were in contact with the basolateral medium. After they obtained a full confluence, the apical medium was removed and cells were kept in the air-liquid interface (ALI) cultures for at least 21 days. BEGM/DMEM/ATRA medium was maintained only basolaterally to differentiate the HBECs. During the cell culture process, medium was exchanged every 2-3 days and, periodically, excess of produced mucus was removed from the wells. All experiments were performed on the fully differentiated HBECs from the same passage, between 21 and 28 days of ALI culture (Fig. S2A, S4A).

### House dust mite stimulation and rhinovirus A16 in vitro infection model in the primary HBECs

House dust mite (HDM) stimulation, followed by rhinovirus A16 (RV-A16) infection experiments were performed in the OptiMEM medium (LifeTechnologies, ThermoFisher Scientific, Waltham, USA). ALI-differentiated HBECs from control individuals and patients with asthma were treated apically with the HDM extract (Allergopharma, Reinbek, Germany) at a dose of 200 μg/mL of the total protein (or vehicle) in 200 μl OptiMEM on the apical side, and 600 μl of clear OptiMEM on the basolateral side (Fig. S2A, S4A), in the humidified incubator at 37°C with 5% CO_2_. After 24h of HDM stimulation cells, were apically infected with RV-A16 at the MOI of 0.1 or as otherwise specified, or stimulated with UV-RV-A16 at the same MOI, and cultured in the humidified incubator at 34.5°C with 5% CO_2_ for the next 24h (Fig. S2A, S4A). Next, cell supernatants (apical and basolateral), RNA, and protein cellular lysates were collected and stored in −80°C. Some cells were fixed with 4% PFA (Fluka/Sigma Aldrich Buch, Switzerland) and were stored wet at 4°C for 1-2 weeks before the subsequent confocal analyses. All doses and time-points used for the final experiments were based on the preliminary dose-dependent and time-course experiments. Briefly, two different HDM extracts: main HDM extract used in the manuscript (Allergopharma, Reinbek, Germany) at the dose of 200 μg/mL, and HDM extract B (Citeq, York, UK) at the dose of 200 and 100 μg/ml were investigated (Fig. S4B, S5A-B). RV-A16-infection of HBECs from patients with asthma was performed in 6h and 24h time-points (Fig. S2B). Lastly, RV-A16 infection at the MOI 0.001, MOI 0.01, and MOI 0.1 was investigated (Fig. S2C). Based on the secretion of the mature IL-1*β*, HDM extract from the Allergopharma, Reinbek, Germany at the dose of 200 μg/mL, RV-A16 MOI 0.1 and the 24h time-point were chosen, and are presented through the manuscript, if not mentioned differently. For the experiments with inhibitors, 40 μM of the caspase-1 inhibitor: ac-YVAD-cmk (Acetyl-tyrosine-valine-alanine-aspartate-chloromethyl ketone, Invivogen, San Diego, USA), 1μM of the IKK*ε*/TBK1 inhibitor: BX795 (N-[3-[[5-iodo-4-[[3-[(2-thienylcarbonyl)amino]propyl]amino]-2-pyrimidinyl]amino]phenyl]-1-Pyrrolidinecarboxamide hydrochloride, Sigma Aldrich, Merck, Kenilworth, USA), or 1μM of the NLRP3 inflammasome inhibitor: MCC950 (C20H23N2NaO5S, Avistron, Bude, UK) or appropriate vehicle controls were used apically and basolaterally, 24h prior RV-A16 infection. To block ICAM-1, a receptor responsible for RV-A16 infection of HBECs, anti-ICAM-1 antibodies were added to the apical and basolateral compartment, 3h prior RV-A16 infection at the dose of 10ug/mL (Fig. S2A).

### Rhinovirus and severe acute respiratory syndrome coronavirus-2 (SARS-COV-2) in vitro co-infection model in primary HBECs from control individuals and patients with asthma

The ALI-differentiated MucilAir cultures (Epithelix, Plan-les-Ouates, Switzerland) from primary human bronchial epithelium obtained from 4 control individuals and 5 patients with asthma (Supplementary Table S9) were cultured for 7 days in the MucilAir Medium (Epithelix, Plan-les-Ouates, Switzerland) in ALI conditions, with basolateral medium changed every other day. At the day of the experiment, performed in a biosafety level 3 (BSL3) laboratory, cells were washed with warm PBS to remove an excess of mucus. The experiment was performed in the OptiMEM medium (LifeTechnologies, ThermoFisher Scientific, Waltham, USA) in the volume of 250 μl on the apical, and 600 μl on the basolateral side. Through the whole experiment cells were kept in the humidified incubator at 37°C with 5% CO_2._ First, HBECs were stimulated apically with 200 μg/mL of protein content of HDM extract or vehicle. 24h after HDM stimulation, cells were apically infected with/without RV-A16 at the MOI of 0.1. After next 24h, HBECs were apically infected with/without SARS-CoV-2 at the MOI of 0.1. Finally, 48h after SAR-CoV-2 infection experiment was harvested (Fig. 6A). In order to inactivate SARS-CoV-2, all collected supernatants were treated with 65°C for 30 min. Cells were fixed in 4% PFA for at least 20 min. Inactivated supernatants were frozen in −80°C until further analyses. For RNA analyses, insert with the fixed cells were preserved in RNAlater (Qiagen, Hilden, Germany), left overnight in 4°C, and stored in −20°C in the new, dry tube. In order to perform confocal staining, inserts with the fixed cells were snap frozen in the Clear Frozen Section Compound (FSC22, Leica, Wetzlar, Germany).

### THP-1 cell culture

THP-1-XBlue cells (Invivogen, San Diego, USA) were defrosted in 32 mL of RPMI-1640 medium (Sigma-Aldrich, St. Louis, USA) supplemented with the Penicillin/Streptomycin/Kanamycin, MEM vitamins, Na-Pyruvate/MEM Non-essential Amino Acid Solution and heat-inactivated FCS (cRPMI medium) in the 75cm^2^ T-flask, and cultured for 1 day in the humidified incubator at 37°C with 5% CO_2_. In the following day, cells were counted, checked for viability (98%) and transferred to the 12-well cell cultures plate (0.5 mio cells/well in 1 mL of cRPMI medium). Next day, cells were stimulated with LPS (100 ng/mL, Invivogen, San Diego, USA) or vehicle for 4h followed by 2 mM ATP or vehicle (Invivogen, San Diego, USA) for 20 min. Cytospins (250xg, 3 mins, Shandon Cytospin 2, Marshall Scientific, Hampton, USA) were prepared, and cells were immediately fixed with 4% PFA (Fluka/Sigma Aldrich, Buchs, Switzerland), and stored in wet chamber before the confocal staining.

### Immunoassays

#### ELISA and MSD multiplex

Secreted IL-1*β* in majority of in vitro experiments was measured with the ELISA kit (R&D Systems, McKinley Place, USA), according to the manufacturers instruction. Detection limit for the kit is 3.91 pg/mL. IL-1*β* in BAL fluid from control individuals and patients with asthma was measured using mesoscale discovery platform (MSD, Kenilworth, USA) kits as described previously^85^. Additionally, BAL samples from controls and patients with asthma before and after experimental intranasal RV-A16 infection in vivo were analyzed with V-PLEX human IL-1*β* Kit (MSD, Kenilworth, USA), according to the manufacturer’s instructions. V-PLEX IL-1*β* is presented as log_10_ arbitrary units (au) corresponding to percentage of total protein concentration measured by BCA (ThermoFisher Scientific, Waltham, USA) and multiplied by factor 1000000.

#### Proximity extension assay (PEA) targeted proteomics

Protein expression in the apical compartments of the HBECs were measured using the proximity extension assay targeted proteomics technology (Olink, Stockholm, Sweden). Targeted 96-proteins Inflammation, Immune Response and Immuno-Oncology panels were used according to the manufacturer’s instructions with the suggested adaptations to the cell cultures conditions. Apical compartments from RV-A16+SARS-CoV-2 model were analysed with the human Target 48 Cytokine Panel (Olink, Uppsala, Sweden). Final results for 96-plex assay are reported as Normalized Protein eXpression (NPX), an arbitrary unit in log_2_-scale. Results from the Target 48 Cytokine Panel are in pg/mL.

#### Western Blotting

Western Blotting experiments from the cell lysates and the apical supernatants were performed as previously described^86, 87^. Briefly, cells were lysed in RIPA Lysis and Extraction buffer (ThermoFisher Scientific, Waltham, USA) supplemented with the protease inhibitor (Roche, Merck, ThermoFisher Scientific, Waltham, USA) for 15 minutes on ice, centrifuged (15 min, full speed, 4°C), and debris-free protein lysates were frozen in −80°C for further analyses. Protein concentration was assessed with the BCA kit (ThermoFisher Scientific, Waltham, USA), according to the manufacturer’s instructions. Protein from the apical supernatants was precipitated with 1 volume of methanol (Fisher Scientific, Reinach, Switzerland) and ¼ volume of chloroform (Merck Millipore, Burlington, USA) as described previously^87^. Equal amounts of cell lysate proteins (10-20μg) were loaded on the 4-20% Mini-PROTEAN TGX Gel (Bio-Rad, Hercules, USA) or 4-20% SuperPAGE gel (GenScript, Leiden, Netherlands) in Tris/Glycine/SDS buffer (Bio-Rad Lab, Hercules, USA) or MOPS buffer (GenScript, Leiden, Netherlands). After electrophoresis, the proteins were transferred to the nitrocellulose membranes (Bio-Rad, Hercules, USA or Advansta, San Jose USA) using the Trans-Blot Turbo Blotting System (Bio-Rad, Hercules, USA) or eBlot L1 Protein Transfer System (GenScript, Leiden, Nederlands). The membranes were blocked with 5% nonfat milk in 0.1% Tween20 PBS (PBST) for 1h, washed with 10x PBST, and incubated with the primary antibodies for overnight in 4°C. Dilutions of primary antibodies used for the analyses of the cell lysates: 1:100 IL-1*β* (R&D Systems, McKinley Place, USA), 1:200 RIG-I (Santa Cruz Biotechnology, Dallas, USA), 1:1000 ASC (Santa Cruz Biotechnology, Dallas, USA), 1:1000 caspase-1 (Cell Signaling, Danvers, USA), and 1:1000 NLRP3 (Adipogen, San Diego, USA). The membranes were subsequently washed in 10xPBST and incubated with an appropriate secondary antibody conjugated with the horseradish peroxidase (HRP) (1:10,000 dilution) (Jackson ImmunoResearch, West Grove, USA; Santa Cruz Biotechnology, Dallas, USA) for 1h at room temperature. *β*-actin expression was determined with HRP-conjugated antibodies in 1:25,000 dilution (Abcam, Cambridge, UK). Protein samples precipitated from the apical supernatants were analyzed with the use of goat anti-IL-1*β* antibodies (1:1000, R&D Systems, Minneapolis, USA) and HRP conjugated anti-goat (1:10,000, Santa Cruz Biotechnology, Santa Cruz, USA) antibodies. After washing with 10xPBST, the blots were developed with the SuperSignal West Femto Kit (ThermoFisher Scientific, Waltham USA) or WesternBright Quantum/Sirius HRP substrate (Advansta, San Jose, USA) and visualized on the Luminescent Image Analyzed LAS-1000 (Fujifilm, Tokyo, Japan) or the Fusion FX (Vilber, Collegien, France). To assess more than one protein, the membranes were stripped with the Restore PLUS Western Blot Stripping Buffer (ThermoFisher Scientific, Waltham, USA). Quantification of the protein expression was performed in Fiji Software. ^88^ Briefly, an area of the peak of the protein of interest was measured in triplicates, and average was used to calculate the ratio between expression of the protein of interest and *β*-actin (protein/*β*-actin). Protein/*β*-actin ratio from HBECs from control individuals and patients with asthma upon HDM stimulation, RV-A16 infection, or both was further normalized to the vehicle control condition from control individuals, and log transformed with Y=log(Y) function.

#### Co-immunoprecipitation

For co-immunoprecipitation cells were lysed with the Lysing Buffer (1μM DTT + 10% Triton X100 in ddH_2_0 supplemented with the protease inhibitor (Roche, Merck, ThermoFisher Scientific, Waltham, USA) for 15 minutes on ice, centrifuged (15 min, full speed, 4°C), and the debris-free protein lysates were frozen in −80°C for further analyses. Protein concentration was assessed with the BCA kit (ThermoFisher Scientific, Waltham, USA) according to the manufacturer’s recommendation. 100 μg of the pooled protein was pre-cleared with 100 μl of Protein A beads (Bio-Rad, Hercules, USA), magnetized, and incubated with 10 μg of anti-ASC antibodies (Santa Cruz Biotechnology, Santa Cruz, USA) overnight at 4°C, followed by ASC immunoprecipitation with 100 μl of Protein A beads (Bio-Rad, Hercules, USA) for 2h in RT. Co-IP samples and input (protein not bound to the beads) were collected and together with the cell lysates were further analyzed with the Western Blot protocol with the use of mouse anti-human RIG-I antibodies (1:200, Santa Cruz Biotechnology, Santa Cruz, USA) or rabbit anti-human MDA5 antibodies (1:1000, Abcam, Cambridge, UK) and HRP conjugated anti-mouse or anti-rabbit antibodies (1:10,000, Jackson ImmunoResearch, West Grove, USA).

### mRNA isolation and RT-PCR

#### HDM and RV-A16 model

Cells were lysed on ice with the RLT buffer (Qiagen, Hilden, Germany) supplemented with *β*-mercaptoetanol (Sigma-Aldrich, St. Louis, USA), and stored at −80°C until further analyses. mRNA isolation was performed with the RNeasy Micro Kit (Qiagen, Hilden, Germany) according to the manufacturer’s instructions. Quality and quantity of isolated RNA was assessed by the Nanodrop 2000 (ThermoFisher Scientific, Waltham, USA). Reverse transcription was performed with RevertAid RT kit (ThermoFisher Scientific, Waltham, USA) with random hexamers, according to the manufacturer’s recommendations. Gene expression (5-10 ng of cDNA/well) was assessed by RT-PCR using the SYBR Green/ROX qPCR Master Mix (ThermoFisher Scientific, Waltham, USA), performed on the QuantStudio 7 Flex Real-Time PCR System (ThermoFisher Scientific, Waltham, USA). The sequences of used primers (5μM) are summarized in the Supplementary Table S10. Gene expression was normalized to the housekeeping gene, elongation factor 1*α* (EEF1A), and presented as a relative quantification calculated with the *ΔΔ*Ct formula, as described previously^42^. Depending on the analyses, data were calibrated according to the vehicle condition from HBECs from control individuals, or vehicle condition calculated separately for control individuals and patients with asthma. Data are presented as 2^-^*^ΔΔ^*^Ct^ values, or percentage change normalized to the specified condition.

#### HDM, RV-A16 and SARS-CoV-2 model

Samples preserved in the RNAlater, as described above, were immersed in the increasing concentrations of ethanol (30% up to 100%, increasing every 10%). After this initial step, RNA was isolated with use of RecoverAll kit (Invitrogen, Waltham, USA) according to the manufacturer’s protocol. Isolated mRNA was concentrated with the use of SpeedVac (DNA Speed Vac, DNA110, Savant, Hayanis, USA) for 1h in the ambient temperature. Sample concentration did not affect its quality, as measured with use of NanoDrop One^C^ (ThermoFisher Scientific, Waltham, USA). Reverse transcription was performed with use of the SuperScript IV VILO Master Mix (Thermofisher Scientific, Waltham USA), according to the manufacturer’s recommendations. Gene expression (5ng of cDNA/well) was assessed by RT-PCR using i) SYBR Green PCR Master Mix (ThermoFisher Scientific, Waltham, USA) for *DDX58*, *IFNB*, *IFNL1*, *IL1B*, *MDA5*, and ii) TaqMan assays for RV-A16 and SARS-CoV-2 *Protein N*, *Protein S*, *ORF1AB* detection (ThermoFisher Scientific, Waltham, USA) and was performed on the QuantStudio 7 Flex Real-Time PCR System (ThermoFisher Scientific, Waltham, USA). The sequences of used primers are summarized in the Supplementary Table S10. Gene expression was normalized to the elongation factor 1*α* (EEF1A) (SYBR Green analyses), or RNase P (TaqMan assays), and presented as a relative quantification calculated with -*ΔΔ*Ct formula, compared to the vehicle condition separately for controls and patients with asthma, as described previously^42^. SARS-CoV-2 viral RNA in presented as 2^-^*^ΔΔ^*^Ct^ values averaged from the expression of *N protein*, *S protein* and *ORF1AB* in each condition.

#### Rhinovirus infection quantification in the nasal and bronchoalveolar lavage fluid in humans

Rhinovirus infection in control individuals and patients with asthma after experimental RV-A16 infection in vivo was performed in the nasal lavages (peak of infection) and BAL fluid (4 days post infection) with use of qPCR, as previously described. ^44^ Results are presented as log_10_ of viral RNA copies in 1 mL.

#### Confocal microscopy

Cells were fixed on the inserts with 4% paraformaldehyde (Fluka/Sigma Aldrich, Buch, Switzerland) for 10 minutes, permeabilized with detergent (PBS + 0.1% TritonX100 + 0.02% SDS) for 5 minutes and blocked with 10% goat serum (Dako, Agilent, Santa Clara, USA) in 1% BSA/PBS for 60 min at room temperature (RT). All antibodies were diluted in 4% goat serum + 1% BSA/PBS, and cells were stained from apical and basolateral sides with 100 μl of antibodies working solution. Samples were stained for ASC (2μg/mL, mouse anti-ASC, Santa Cruz Biotechnology, Santa Cruz, USA), IL-1*β* (10μg/mL, mouse anti-IL-1*β*, Abcam, Cambridge, UK), and RIG-I (2μg/mL, mouse anti-RIG-I, Santa Cruz Biotechnology, Santa Cruz, USA) for 60 minutes at RT. Proper mouse isotype controls, in the corresponding concentrations were used to control for unspecific binding. (Dako, Agilent, Santa Clara, USA). Subsequently, samples were incubated with the goat anti-mouse Alexa Fluor 488 (for ASC), and the goat anti-mouse Alexa Fluor 546 (for IL-1*β* and RIG-I) secondary antibodies at the concentrations of 1:2000 (Invitrogen, Waltham, USA) for 60 minutes at RT. Samples were mounted in the ProLong Gold mounting medium containing DAPI (Life Technologies, Carlsbad, USA) according to the manufacturer’s instructions, analyzed with a Zeiss LSM780 confocal microscope (Zeiss, Oberkochen, Germany) and Zen Software (Zeiss, Oberkochen, Germany). All pictures were taken at the 40x magnification and are presented as maximal projection (orthogonal projection) from z-stacks (3-22 for ASC, 4 for IL-1*β* and RIG-I). ¼ of the original picture is shown on the figures, with appropriate scale bar and further magnification of the area of interest. Additionally, to quantify ASC speck formation specks from 3-5 pictures per condition were counted in duplicates and averaged.

Differentiated HBECs upon HDM+ RV-A16 stimulation and THP-1 cells stimulated with LPS+ATP were used for NLRP3 and Occludin visualization, whereas HBECs from RV-A16+SARS-CoV-2 model were used for ACE2 and SARS-CoV-2 Protein N staining. Following fixation in 4% (wt/vol) PFA in PBS (Fluka, Fluka/Sigma Aldrich, Buchs, Switzerland), THP-1 cytospins were lined with the PAP pen (Sigma Aldrich, St. Louis, USA), HBECs were stained on the insert, whereas ¼ of the ALI insert from RV-A16+SARS-CoV-2 model were prepared for cryosections by freezing in Clear Frozen Section Compound (FSC22, Leica, Wetzlar, Germany), cut at 6 µm in a cryostat (LEICA CM3050S, Leica Microsystems, Wetzlar, Germany) and mounted on *SuperFrost Plus^TM^* glass slides (Menzel, ThermoFisher, Waltham, USA). Samples were incubated in the blocking solution (10% normal goat serum, 1% bovine serum albumin and 0.2% TritonX-100 in PBS) (Dako, Agilent, Santa Clara, USA) for 1h at RT. Primary antibodies for NLRP3 (5 µg/mL, mouse anti-NLRP3, Adipogen, San Diego, USA), Occludin (2.5 µg/mL, mouse anti-Occludin, ThermoFisher, Waltham, USA), ACE-2 (2 µg/mL, rabbit anti-ACE2, Abcam, Cambridge, UK), and SARS-CoV-2 Protein N (1 µg/mL, mouse anti-Protein N, ThermoFisher, Waltham, USA) diluted in blocking solution (1:1 in PBS) and incubated at 4°C overnight. Proper isotype controls, in the concentrations corresponding to the antibodies, were used (Dako, Agilent, Santa Clara, USA). Following three washing steps in 0.05% Tween20 in PBS, secondary antibodies with DAPI (1 µg/mL, Sigma Aldrich, St. Louis, USA) in diluted blocking solution (1:1 in PBS) were applied for 2h at RT in the dark. Sections have been washed three times in 0.05%Tween20 in PBS before mounting with Fluoromount (Sigma Aldrich, St. Louis, USA). Image acquisition was performed with Zeiss LSM780 (Zeiss, Oberkochen, Germany), by using 40x objective and ZEN software (Zeiss, Oberkochen, Germany). ImageJ/Fiji^88^ was used for tale scan stitching and image analysis.

Bronchial biopsies were collected from the control individuals and patients with asthma at baseline and 4 days after in vivo RV-A16 infection. Biopsies were fixed and embedded in the paraffin blocks, sections were cut, and placed on the glass slides as described previously^89^. Prepared slides were baked 30 minutes in 65°C, followed by the deparaffinization with xylol (2×10 min), graded isopropanol (2×3 mins 100%, 2×3 mins 96% and 3 mins 70%), and rehydration (2×5 mins in H_2_0_2_). Samples were boiled in the sodium citrate buffer (10mM sodium citrate with 0.05% Tween 20 in PBS at pH6) in the pressure cooker for 4 mins, as described previously^90^. Samples were permeabilized and blocked with the Perm/Block Buffer (1%BSA+0.2%TritonX100+10% goat serum in PBS) for 25 minutes in RT. All antibodies were diluted in 4% goat serum + 0.05% Tween20 in PBS, and 50 μl of antibodies per sample were used. Primary antibodies for IL-1*β* (10 μg/mL, mouse anti-IL1*β*, Abcam, Cambridge, UK), caspase-1 (1:50, rabbit anti-caspase 1, Cell Signaling, Danvers, USA), and RIG-I (1:250, mouse anti RIG-I, Santa Cruz Biotechnology, Santa Cruz, USA) were incubated in the wet chamber overnight at 4°C. Appropriate mouse and rabbit isotype controls, in the concentrations corresponding to the antibodies, were used (Dako, Agilent, Santa Clara, USA). Subsequently, samples were incubated with the goat anti-rabbit Alexa Fluor 488 (for caspase-1), and the goat anti-mouse Alexa Fluor 546 (for IL-1*β* and RIG-I) secondary antibodies for 60 minutes in RT in the concentration of 1:1000 (Invitrogen, Waltham, USA). After 3 minutes’ incubation in 1% PFA in room temperature, samples were mounted in the ProLong Gold mounting medium containing DAPI (Life Technologies, Carlsbad, USA) according to the manufacturer’s instructions, analyzed with a Zeiss LSM780 (Zeiss, Oberkochen, Germany) and Zen Software (Zeiss, Oberkochen, Germany). All pictures were taken at the 40x magnification and are presented as the maximal projection (orthogonal projection) from 4 z-stacks, with appropriate scale bar. For the quantification, 10 equal squares from epithelial areas of the tissue (assessed using Haematoxylin and Eosin staining of the adjacent slice) from stained and isotype control samples were measured for signal intensity and averaged. Isotype control signal was further subtracted from the intensity of the stained samples, and values from 10 squares per sample were presented as the mean fluorescent intensity (MFI) of the protein expression.

#### Transcriptome analyses

Next generation sequencing (NGS) from the differentiated human bronchial epithelial cells (HBECs) from control individuals and patients with asthma (cohort A) was performed as previously described^75^. Briefly, total RNA was isolated with a RNeasy Plus Micro Kit (Qiagen, Hilden, Germany). Library was prepared with the TruSeq Stranded mRNA Sample Prep Kit (Illumina, San Diego, USA), and sequenced on the Illumina HiSeq 4000 platform. Description of the study subjects is presented in the Supplementary Table S8.

HBECs from 6 control individuals and 6 patients with asthma, infected with RV-A16 in the MOI 10 for 24h were harvested and sequenced with the use of Illumina HiSeq 2000 platform, as described previously^45^. The mRNA expression data are publicly available at the Gene Expression Omnibus platform (https://www.ncbi.nlm.nih.gov/) under the accession number: GSE61141^45^.

Bronchial brushings from control individuals and patients with asthma before and after experimental RV-A16 infection in vivo were analyzed by Affymetrix HuGene 1.0 array according to the manufacturer’s instructions and Transcriptome Analysis Console v4.0 (Santa Clara, United States).

#### Data analysis

Distribution (normality) of the data was assessed with Shapiro-Wilk test. One-way ANOVA (Kruskal-Wallis test), RM one-way ANOVA (Friedman test) or mixed-effects model tests were performed for more than three groups comparisons depending on the data relation (paired/not-paired) and distribution (normal/not-normal). Two-tailed paired/not-paired t-test or Wilcoxon/U-Mann-Whitney tests were performed for two groups comparisons depending on the data relation and distribution. The data are presented as the mean ± SEM, with the number of samples in each experiment indicated in the figure description. IL-1*β* expression in BAL fluid from cohort SIBRO was analyzed with the Welsh’s test. All differences were considered significant when p≤0.05 and defined as *p-value≤0.05, **p-value≤0.01, ***p-value≤0.001, ****p-value≤0.0001. Statistical analysis was performed with the Prism 9 software (Redmond, USA).

Transcriptome data were processed with the workflow available at https://github.com/uzh/ezRun, with the significance threshold for differentially expressed genes set to p-value<.05 calculated for the entire gene lists in each project using the edgeR R package^91^. Microarray data was analyzed by the following Bioconductor microarray analysis workflow https://www.bioconductor.org/packages/release/workflows/vignettes/arrays/inst/doc/arrays.html.

Differentially expressed probe was identified by the limma R package with empirical Bayes estimation. Threshold for significance for transcriptome data presented on the figures are as follows: p-value: *<0.05; **<0.005; ***<0.0005, ****<0.00005. Heatmaps display normalized gene expression across the gene in the groups (row normalization). Additionally, enrichment analysis of the most significant process networks in bronchial brushings after in vivo RV-A16 infection in patients with asthma when compared with bronchial brushings from control individuals after in vivo RV-A16 infection (Asthma (RV-A16 infection vs baseline) VS Control (RV-A16 infection vs baseline)) was performed with Metacore software version 19.2.69700 (Thomson Reuters, Toronto, Canada) (Supplementary Table S1). Top 100 genes upregulated after RV-A16 infection in the HBECs from control individuals and patients with asthma from GSE61141^45^ were analyzed for the enriched pathways using Metacore software version 20.3.70200 (Thomson Reuters, Toronto, Canada) (Supplementary Table S2). Inflammasome-mediated immune responses and antiviral responses gene sets were curated from GSEA and MSigDB Database (Broad Institute, Massachusetts Institute of Technology, and Reagent of the University of California, USA). Full sets of analyzed genes are described in the Supplementary Table S11.

Proximity Extension Assay (PEA) normalizes protein expression (NPX) data were analyzed with the use of the internal Shiny App Olink data analysis toolkit http://46.14.201.130:3838/OlinkR_shinyApp. The statistical comparison of protein expression between groups was performed with the Bioconductor limma package^92^. The fold change and p-value were estimated by fitting a linear model for each protein. Proteins with p-value<0.05 were considered significant. Additionally, for Target 96 Inflammation panel data are presented as: i) heatmaps of curated signatures of inflammasome-mediated immune responses and antiviral responses (Supplementary Table S11) and ii) protein interactions and pathways analysis prepared using the STRING (version 11.0)^93^, and further processed with the Cytoscape software (version 3.8.2)^94^ (Supplementary Table S4). The list of all proteins available for PEA measurements at the moment of the current analysis, that were used as a background reference for STRING analyses for targeted proteomics data is presented in Supplementary Table S12.

## Abbreviations

*ASC*: apoptosis-associated speck-like protein containing CARD

*BAL*: bronchoalveolar lavage

*BX795*: inhibitor of TBK1 and IKK*ε*

*COVID-19*: coronavirus disease 2019

*HBECs*: human bronchial epithelial cells

*HDM*: house dust mite

*IAV*: influenza A virus

*ICAM-1*: intracellular adhesin molecule-1

*IFN*: interferon

*IKKε*: inhibitor-κB kinase epsilon

*ISG*: interferon-stimulated genes

*MDA5*: melanoma differentiation-associated protein 5

*MOI*: multiplicity of infection

*NLRP3*: NLR family pyrin domain containing 3

*RV*: rhinovirus

*RV-A16*: rhinovirus A16

*RIG-I*: retinoic acid-inducible gene I

*SARS-CoV-2*: severe acute respiratory syndrome virus 2

*TBK1*: TANK binding kinase 1 UV-light-inactivated rhinovirus A16

*YVAD*: caspase-1 inhibitor.

## 5. Data Availability

Transcriptome data from bronchial brushings from control individuals and patients with asthma experimentally infected with RV-A16 has been submitted to the NCBI GEO: GSE185658 and will be publicly available at the time of publication. All other data are included in the Online Supplement or are available from the corresponding author upon request. The codes for transcriptome data analysis are available here: https://github.com/uzh/ezRun (NGS), https://github.com/ge11232002/p1688-Ula (microarray). Code for Proximity Extension Assay data analysis is available from the corresponding author upon request. All codes will be publicly available at the time of publication.

## 6. Code availability

The codes for transcriptome data analysis are available here: https://github.com/uzh/ezRun (NGS), https://github.com/ge11232002/p1688-Ula (microarray). Code for Proximity Extension Assay data analysis is available here: https://github.com/ge11232002/OlinkR. All codes will be publicly available at the time of publication.

## Acknowledgements

The Authors are grateful to all patients, clinicians and research staff involved in securing and processing patients samples. We would like to thank David Mirer for rhinovirus plaque assay analyses and Dr Anna Globinska for help with graphical abstract design. HDM extract was a gift from Allergopharma AG. This work was supported by the Swiss National Science Foundation (SNSF) grant nr 310030_189334/1, European Respiratory Society Long-term research fellowships, European Academy of Allergy and Clinical Immunology long-term research fellowship and GSK research grant (to MS), and the SNF grant nr 320030_176190 and GSK (to CA). The in vivo work was supported by the European Research Council (ERC FP7 grant number 233015); a Chair from Asthma UK (CH11SJ); the Medical Research Council Centre (grant number G1000758); National Institute of Health Research (NIHR) Biomedical Research Centre (grant number P26095); Predicta FP7 Collaborative Project (grant number 260895); and the NIHR Biomedical Research Centre at Imperial College London. SLJ is an Emeritus NIHR Senior Investigator. The in vivo transcriptome work was supported by GSK. UR, AE and MM were supported from funds from the Leading National Research Centre (KNOW) in Bialystok, Poland. JRC was supported by an FPI-CEU predoctoral fellowship and the Swiss-European Mobility Programme.

## 7. Authors contributions

Concept design: MiSo. Data collection: UR, AE, MiSo, NS, AH, PaWe, SS, AD, PaWa, BR, JRC, DZ, MH, JB, MaSa, LOM, DJJ, MRE, TK. Analysis/interpretation of data: UR, AE, MiSo, GT, CA. Write/intellectual contribution: UR, AE, MiSo, CA, MM, TV, SJ. All authors approved the final version of the manuscript.

## 8. Ethics declaration

CA reports research grants from Allergopharma, Idorsia, Swiss National Science Foundation, Christine Kühne-Center for Allergy Research and Education, European Commission’s Horison’s 2020 Framework Programme “Cure”, Novartis Research Institutes, Astrazeneca, SciBase, Stanford University SEAN Parker Asthma and Allergy Center; advisory board of Sanofi/Regeneron, GSK and Novartis, consulting fees from Novartis; Editor-in-Chief Allergy, Co-Chair EAACI Environmental Science in Allergic Diseases and Asthma Guidelines. AE reports National Science Centre Grant No. 2020/37/N/NZ5/04144, National Centre for Research and Development No. STRATEGMED2/269807/14/NCBR/2015, National Centre for Research and Development (POLTUR3/MT-REMOD/2/2019). DJJ reports advisory board and speaker fees from AstraZeneca, GSK and Sanofi and research grants from AstraZeneca. SLJ reports grants/contracts from European Research Council ERC FP7 grant number 233015, Chair from Asthma UK CH11SJ, Medical Research Council Centre grant number G1000758, NIHR Biomedical Research Centre grant number P26095, Predicta FP7 Collaborative Project grant number 260895, NIHR Emeritus NIHR Senior Investigator; consulting fees from Lallemand Pharma, Bioforce, resTORbio, Gerson Lehrman Group, Boehringer Ingelheim, Novartis, Bayer, Myelo Therapeutics GmbH; patents issued/licensed: Wark PA, Johnston SL, Holgate ST, Davies DE. Anti-virus therapy for respiratory diseases. UK patent application No. GB 0405634.7, 12March 2004. Wark PA, Johnston SL, Holgate ST, Davies DE. Interferon-Beta for Anti-Virus Therapy for Respiratory Diseases. International Patent Application No. PCT/GB05/50031, 12 March 2004. Davies DE, Wark PA, Holgate ST, JohnstonSL. Interferon Lambda therapy for the treatment of respiratory disease. UK patent application No. 6779645.9, granted15th August 2012; Participation on a data safety monitory board or advisory board: Enanta Chair of DSMB, Virtus Respiratory Research Board membership. MM reports personal payments from Astra Zeneca, GSK, Berlin-Chemie/Menarini, Lek-AM, Takeda, Celon and support for attending meetings from Astra Zeneca, GSK, Berlin-Biochemie/Menarini. UR reports board secretary position of Working Group of Genomics and Proteomics of the European Academy of Allergy and Clinical Immunology (EAACI). JRC reports Pre-doctoral grant FPI from Universidad CEU San Pablo, Swiss European Mobility Program grant from University of Zurich, EAACI Mid-term Fellowship. MiSo reports research grants from Swiss National Science Foundation, GSK, Novartis and speakers fee from AstraZeneca and board secretary position of the Basic and Clinical Immunology Section of the European Academy of Allergy and Clinical Immunology (EAACI). SS reports funding from National Center of Competence in Research (NCCR) on RNA and Disease to VT (https://nccr-rna-and-disease.ch/). VT reports grant from Swiss National Science Foundation. All other authors report no conflict of interest regarding this work.

## Notes

### Competing Interest Statement

CA reports research grants from Allergopharma, Idorsia, Swiss National Science Foundation, Christine Kuhne-Center for Allergy Research and Education, European Commissions Horisons 2020 Framework Programme Cure, Novartis Research Institutes, Astrazeneca, SciBase, Stanford University SEAN Parker Asthma and Allergy Center; advisory board of Sanofi/Regeneron, GSK and Novartis, consulting fees from Novartis; Editor-in-Chief Allergy, Co-Chair EAACI Environmental Science in Allergic Diseases and Asthma Guidelines. AE reports National Science Centre Grant No. 2020/37/N/NZ5/04144, National Centre for Research and Development No. STRATEGMED2/269807/14/NCBR/2015, National Centre for Research and Development (POLTUR3/MT-REMOD/2/2019). DJJ reports advisory board and speaker fees from AstraZeneca, GSK and Sanofi and research grants from AstraZeneca. SLJ reports grants/contracts from European Research Council ERC FP7 grant number 233015, Chair from Asthma UK CH11SJ, Medical Research Council Centre grant number G1000758, NIHR Biomedical Research Centre grant number P26095, Predicta FP7 Collaborative Project grant number 260895, NIHR Emeritus NIHR Senior Investigator; consulting fees from Lallemand Pharma, Bioforce, resTORbio, Gerson Lehrman Group, Boehringer Ingelheim, Novartis, Bayer, Myelo Therapeutics GmbH; patents issued/licensed: Wark PA, Johnston SL, Holgate ST, Davies DE. Anti-virus therapy for respiratory diseases. UK patent application No. GB 0405634.7, 12March 2004. Wark PA, Johnston SL, Holgate ST, Davies DE. Interferon-Beta for Anti-Virus Therapy for Respiratory Diseases. International Patent Application No. PCT/GB05/50031, 12 March 2004. Davies DE, Wark PA, Holgate ST, JohnstonSL. Interferon Lambda therapy for the treatment of respiratory disease. UK patent application No. 6779645.9, granted15th August 2012; Participation on a data safety monitory board or advisory board: Enanta Chair of DSMB, Virtus Respiratory Research Board membership. MM reports personal payments from Astra Zeneca, GSK, Berlin-Chemie/Menarini, Lek-AM, Takeda, Celon and support for attending meetings from Astra Zeneca, GSK, Berlin-Biochemie/Menarini. UR reports board secretary position of Working Group of Genomics and Proteomics of the European Academy of Allergy and Clinical Immunology (EAACI). JRC reports Pre-doctoral grant FPI from Universidad CEU San Pablo, Swiss European Mobility Program grant from University of Zurich, EAACI Mid-term Fellowship. MiSo reports research grants from Swiss National Science Foundation, GSK, Novartis and speakers fee from AstraZeneca and board secretary position of the Basic and Clinical Immunology Section of the European Academy of Allergy and Clinical Immunology (EAACI). SS reports funding from National Center of Competence in Research (NCCR) on RNA and Disease to VT (https://nccr-rna-and-disease.ch/). VT reports grant from Swiss National Science Foundation. All other authors report no conflict of interest regarding this work.

### Author Declarations

The in vivo experimental infection with rhinovirus study received ethical approval from the St. Marys Hospital Research Ethics Committee (09/H0712/59). The cohort SIBRO was granted ethical permission from Switzerland and Poland (KEK/ZH/Nr. 20212/0043 Kantonale Ethik Kommission Zurich; KB/70/2013 and KB/567/2014 Bioethical Committee, Wroclaw Medical University), the cohort A got a permission from the Jagiellonian University Bioethics Committee (KBET/68/B/2008 and KBET/209/B/2011).

