## Supplementary Tables for "Rhinovirus-induced epithelial RIG-I inflammasome activation suppresses antiviral immunity and promotes inflammatory responses in virus-induced asthma exacerbations and COVID-19"

&Senior Co-Authorship

* Corresponding Author

[1] Swiss Institute of Allergy and Asthma Research (SIAF), University of Zurich, Davos, Switzerland

[2] Christine Kühne – Center for Allergy Research and Education (CK-CARE), Davos, Switzerland

[3] Department of Regenerative Medicine and Immune Regulation, Medical University of Bialystok, Bialystok, Poland

[4] Functional Genomics Center Zurich, ETH Zurich/University of Zurich, Zurich, Switzerland

[5] Institute of Virology and Immunology (IVI), Bern, Switzerland

[6] Department of Infectious Diseases and Pathobiology, Vetsuisse Faculty, University of Bern, Bern, Switzerland

[7] Graduate School for Cellular and Biomedical Sciences, University of Bern, Bern, Switzerland

[8] Division of Clinical Chemistry and Biochemistry, University Children`s Hospital Zurich and Children`s Research Center, University Children`s Hospital Zurich, Zurich, Switzerland

[9] IMMA, Department of Basic Medical Sciences, Facultad de Medicina, Universidad San Pablo-CEU, CEU Universities Madrid, Spain; Centre for Metabolomics and Bioanalysis (CEMBIO), Department of Chemistry and Biochemistry, Facultad de Farmacia, Universidad San Pablo-CEU, CEU Universities Madrid, Spain

[10] Department of Internal Medicine, Jagiellonian University Medical College, Cracow, Poland

[11] Department of Allergology and Internal Medicine, Medical University of Bialystok, Bialystok, Poland

[12] Department of Medicine and School of Microbiology, APC Microbiome Ireland, University College Cork, Ireland

[13] National Heart and Lung Institute Imperial College London, United Kingdom; Asthma UK Centre in Allergic Mechanisms of Asthma, London, United Kingdom

[14] Guy's Severe Asthma Centre, School of Immunology & Microbial Sciences, King's College London, London, United Kingdom

[15] Guy's & St Thomas' NHS Trust SE1 9RT; GSTT & South Thames Asthma Network Guy's Severe Asthma Centre, Guy's Hospital; Faculty of Life Sciences & Medicine King's College London, London, United Kingdom

[16] National Heart and Lung Institute Imperial College, London, United Kingdom; Asthma UK Centre in Allergic Mechanisms of Asthma, London, United Kingdom; Imperial College Healthcare NHS Trust, London, United Kingdom

**Correspondence:**

Milena Sokolowska, MD, PhD

Head of Immune Metabolism

Swiss Institute of Allergy and Asthma Research (SIAF)

Herman-Burchard-Strasse 9

CH-7265 Davos Wolfgang

### Supplementary Tables

### Supplementary Table S1. Enrichment analysis of the most significant process networks in bronchial brushings after in vivo rhinovirus A16 (RV-A16) infection in patients with asthma when compared with control individuals.

| Genes changed in Asthma after RV-A16 infection (RV-A16 infection vs baseline) VS genes changed in Control after RV-A16 infection (RV-A16 infection vs baseline) | | | | | |
|---|---|---|---|---|---|
| | Networks | Total | p-value | In data | Network objects from active data |
| 1 | Immune response_Phagosome in antigen presentation | 243 | 6.154E-22 | 124 | NF-kB p50/p50, Profilin I, RhoA, TAP1 (PSF1), I-kB, Beta-2-microglobulin, ITGB1, SHPS-1, PSMD3, PSMD1, PSMD2, ROCK, PSMA1, MHC class II, PI3K cat class IA, Hck, ELMO2, GRP78, Profilin, CD74, TRAM1, PSMB4, Slp76, p38beta (MAPK11), CD14, NCK1, GRB2, NF-kB1 (p105), PSMB6, alpha-M/beta-2 integrin, NF-kB1 (p50), ROCK2, PSMA4, c-Cbl, PSME2, Vinculin, PKC-alpha, MSN (moesin), PI3K cat class IA (p110-beta), ERM proteins, PSMD7, IKK-alpha, VASP, Fc epsilon RI gamma, C3, Lyn, N-WASP, TLR4, Calreticulin, IKK-beta, PSMB2, DOCK1, ROCK1, TAP2 (PSF2), BLNK, FGR, ELMO1, SEC61 beta, PSMB1, MHC class I, PSMB9, HLA-DPA1, PSMD11, Calnexin, PSMB3, alpha-5/beta-1 integrin, WASP, Actin cytoskeletal, PSMF1, HLA-DQB1, JNK(MAPK8-10), PSMD14, IKK (cat), SHP-1, PA28 (11S regulator), PSMB7, PSMD5, VCP, JNK3(MAPK10), iC3b, PSMD12, Fc epsilon RI beta, Legumain, NF-kB, Actin cytoplasmic 2, RDX (radixin), C3dg, FYB1, PSMA7, SEC61 gamma, PSMA3, CD21, HSP90, NF-kB p50/p65, Tapasin, PSMA5, JMJD6, HSP70, LAT, ITGA5, HLA-DQA1, RalA, Sec10, TAP, JNK1(MAPK8), Actin, Derlin1, p38 MAPK, Btk, HSP90 beta, PSMA6, ERp72, HLA-DPB1, PSMA2, NFKBIA, PSME1, FPR, ACTB, Paxillin, Cofilin, Rac1, IP3 receptor, Immunoproteasome (20S core), NF-kB p65/p65 |
| 2 | Immune response_Antigen presentation | 197 | 3.137E-18 | 101 | STAT3, NF-kB p50/p50, TAP1 (PSF1), IFNGR1, I-kB, Beta-2-microglobulin, JAK2, PSMD3, PSMD1, IP-30, HLA-DRB1, PSMD2, STAT2, PSMA1, CD80, MHC class II, NFYC, GRP78, CD74, CD45, PSMB4, NF-kB1 (p105), PSMB6, alpha-M/beta-2 integrin, NF-kB1 (p50), PSMA4, JAK1, ICAM1, PSME2, PSMD8, PSMD7, IKK-alpha, Fc epsilon RI gamma, TRAP-1, Calreticulin, TRAF6, IKK-beta, PSMB2, IFN-gamma, ECM29, TAP2 (PSF2), SEC61 beta, HLA-DMA, ICAM3, PSMB1, CREB1, MHC class I, PSMB9, HLA-DPA1, PSMD11, Calnexin, PSMB3, HLA-DOA, HLA-F, PSMF1, HLA-DQB1, IFNGR2, PSMD14, IKK (cat), PA28 (11S regulator), IFN-gamma receptor, PSMB7, PSMD5, CD40(TNFRSF5), CEACAM1, PSMD12, Legumain, NF-kB, PSMA7, SEC61 gamma, PSMA3, SP1, HSP90, NF-kB p50/p65, ITGAM, Tapasin, MHC class II beta chain, PSMA5, RFX5, HSP70, TNF-R1, ICOS, HLA-DQA1, KLRK1 (NKG2D), HLA-DMB, ULBP1, LFA-3, TAP, AP-3 beta subunits, HSP90 beta, PSMA6, HLA-DPB1, PSMA2, CD3 delta, CD86, NFKBIA, PSME1, STAT1, Immunoproteasome (20S core), NF-kB p65/p65, TNF-alpha |
| 3 | Inflammation_Interferon signaling | 110 | 7.886E-17 | 66 | STAT3, IRF8, IL-1 beta, IFI17, TAP1 (PSF1), IFNGR1, JAK2, MIG, IDO1, MxA, STAT2, MxB, Pyrin (MEFV), IFI27, iNOS, PML, c-Fos, ISG15, JAK1, ICAM1, PKR, CCL2, IFI6, I-TAC, C/EBPbeta, CCL8, ISGF3, ISG54, IL18RAP, ISG20, SSAT, SOCS3, IFN-gamma, TAP2 (PSF2), FasR(CD95), MNDA, IFNAR2, STAT1/STAT2, SERPINB9, IRF7, IRF2, IFP 35, MIP-1-beta, STAT5, IRF9, IFNGR2, IFITM2, SHP-1, IFN-gamma receptor, CD40(TNFRSF5), Apo-2L(TNFSF10), IFN-alpha/beta receptor, IFI44, IRF1, IFNAR1, Caspase-1, Caspase-8, ILT4, TLR3, KLF4, IFI56, CD86, ILT3, TIMP1, STAT1, GBP2 |
| 4 | Immune response_Phagocytosis | 223 | 1.195E-13 | 101 | NF-kB p50/p50, Profilin I, RhoA, Dectin-1, alpha-X/beta-2 integrin, I-kB, PKC-beta, ITGB1, SHPS-1, ROCK, PI3K cat class IA, IL-15RA, ILT2, Hck, PRK2, RelA (p65 NF-kB subunit), ELMO2, Profilin, PKC-beta1, Slp76, p38beta (MAPK11), CD14, NCK1, GRB2, NF-kB1 (p105), alpha-M/beta-2 integrin, c-Fos, NF-kB1 (p50), ROCK2, ITGB2, c-Cbl, Myosin I, Vinculin, PKC-alpha, p22-phox, MSN (moesin), PI3K cat class IA (p110-beta), ERM proteins, IKK-alpha, VASP, Fc epsilon RI gamma, C3, CORO1A(CLABP, TACO), Lyn, N-WASP, C3b, TLR4, Calreticulin, IKK-beta, MRLC, DOCK1, ROCK1, gp91-phox, c-Jun, BLNK, FGR, ELMO1, C/EBP, Fc gamma RII beta, Myosin II, c-Jun/c-Fos, alpha-5/beta-1 integrin, WASP, Actin cytoskeletal, JNK(MAPK8-10), IKK (cat), SHP-1, PKC-gamma, JNK3(MAPK10), VAV-3, iC3b, Fc epsilon RI beta, NF-kB, Actin cytoplasmic 2, RDX (radixin), C3dg, p40-phox, FYB1, p67-phox, CD21, NF-kB p50/p65, JMJD6, MELC, APOLPA, LAT, ITGA5, ILT4, MyHC, PKC-epsilon, JNK1(MAPK8), Actin, p38 MAPK, Btk, NFKBIA, CD47, ACTB, Paxillin, Cofilin, Rac1, IP3 receptor, NF-kB p65/p65 |
| 5 | Inflammation_Neutrophil activation | 215 | 7.577E-11 | 92 | Syntaxin 6, STAT3, NF-kB p50/p50, GRO-2, FPRL1, RhoA, I-kB, PKC-beta, TNF-R2, ROCK, PI3K cat class IA, AP-1, G-protein alpha-15, PA2G6, iNOS, p38beta (MAPK11), GRB2, alpha-M/beta-2 integrin, TRAF3, NF-kB1 (p50), ROCK2, ITGB2, JAK1, ICAM1, Cytochrome b-558, MEKK1(MAP3K1), CCL2, PKC-alpha, IL-10, p22-phox, PI3K cat class IA (p110-beta), Apaf-1, IKK-alpha, IL-8, PSGL-1, cPLA2, c-Jun/c-Jun, G-protein alpha-i family, tBid, CX3CR1, H-Ras, Adenylate cyclase, IKK-beta, PAK2, IFN-gamma, Caspase-3, ROCK1, gp91-phox, c-Jun, SODD, NSGPeroxidase, Adenylate cyclase type VI, IL-6, G-protein beta/gamma, Galpha(i)-specific peptide GPCRs, c-Jun/c-Fos, Actin cytoskeletal, Rac2, VTI1B, IKK (cat), PA24A, VAV-3, NF-kB, p40-phox, p67-phox, G-protein alpha-q/11, Syntaxin 1A, NF-kB p50/p65, ITGAM, PLA2, TNF-R1, IL8RA, Caspase-8, SNAP-23, Adenylate cyclase type IX, Actin, GRO-3, p38 MAPK, Btk, G-protein alpha-i2, PKC-beta2, NFKBIA, FPR, ACTB, Cofilin, Rac1, IP3 receptor, ATF-2/c-Jun, IL8RB, NF-kB p65/p65, Bid, TNF-alpha |
| 6 | Inflammation_Amphoterin signaling | 118 | 4.657E-10 | 58 | NF-kB p50/p50, RhoA, IL-1 beta, I-kB, ROCK, PI3K cat class IA, AP-1, RelA (p65 NF-kB subunit), p38beta (MAPK11), NF-kB1 (p105), alpha-M/beta-2 integrin, NF-kB1 (p50), ROCK2, ITGB2, Calgranulin A, ICAM1, MyD88, CCL2, PI3K cat class IA (p110-beta), IKK-alpha, Calgranulin B, IL-8, S100B, c-Jun/c-Jun, TLR4, Calgranulin C, H-Ras, TRAF6, IKK-beta, MRLC, ROCK1, c-Jun, IL-6, c-Jun/c-Fos, |

| | | | | | |
|---|---|---|---|---|---|
| | | | | | WASP, Actin cytoskeletal, S100P, JNK(MAPK8-10), IKK (cat), IL1RN, JNK3(MAPK10), NF-kB, SP1, NF-kB p50/p65, ITGAM, MELC, MyHC, JNK1(MAPK8), Actin, p38 MAPK, NFKBIA, ACTB, TLR2, Cofilin, Rac1, NF-kB p65/p65, TNF-alpha, RAGE |
| 7 | Cytoskeleton_Regulation of cytoskeleton rearrangement | 183 | 2.707E-09 | 78 | Plectin 1, Tubulin beta, Profilin I, Tubulin gamma, RhoA, RAP-1A, Desmin, 14-3-3 beta/alpha, RhoGDI beta, ROCK, Galpha(i)-specific amine GPCRs, CAPZA, Tubulin beta 1, Tubulin alpha, Cofilin, non-muscle, Profilin, PKC, CAPZ beta, NCK1, ARP3, CD44, PTEN, Vinculin, MSN (moesin), Tubulin gamma 1, ERM proteins, DIA1, Filamin B (TABP), CAPZA1, ACK1, N-WASP, Beta-fodrin, G-protein alpha-i family, DAL1, Zyxin, Nebulin, DOCK2, 14-3-3 zeta/delta, MRLC, G-protein alpha-o, DOCK1, ROCK1, VAV-2, GRAF, Thymosin beta-10, ELMO1, 14-3-3, ARPC1B, Myosin II, SPTBN(spectrin1-4), G-protein beta/gamma, RAP-2A, WASP, Actin cytoskeletal, ARF3, Filamin A, VAV-3, RDX (radixin), Spectrin beta 4, 14-3-3 eta, ARF1, ARPC5, MELC, ARPC1, MyHC, FGD1, PLK1, Actin, ECT2, Tubulin beta 2, ARPC2, ACTB, Paxillin, Cofilin, Actin muscle, Rac1, WaspIP, Tubulin (in microtubules) |
| 8 | Inflammation_Innate inflammatory response | 180 | 7.200E-09 | 76 | STAT3, NF-kB p50/p50, COX-2 (PTGS2), IL-1 beta, TLR1, I-kB, DMBT1, PKC-beta, APOBEC3G, G-protein alpha-15, PGRP-S, IRAK4, iNOS, p38beta (MAPK11), CD14, IP10, NF-kB1 (p50), ECSIT, IL-18, MyD88, Beta-defensin 1, MEKK1(MAP3K1), RIPK2, I-TAC, IKK-alpha, C3, IL-8, TLR6, PLAP, Factor I, cPLA2, C3b, G-protein alpha-i family, TLR4, SP-D, TRAF6, IKK-beta, IP3R3, IRAKM, IL1RAP, RIPK1, c-Jun, TBK1, MD-2, IRF7, IL-6, G-protein beta/gamma, SP-C, JNK(MAPK8-10), IKK (cat), IRF3, TANK, PLUNC, JNK3(MAPK10), NF-kB, TAB2, NF-kB p50/p65, Tissue factor, PLA2, TLR3, IP3R2, C3a, C5 convertase (C3b2Bb), JNK1(MAPK8), p38 MAPK, Btk, G-protein alpha-i2, PKC-beta2, NFKBIA, C3aR, sCD14, TLR2, ERAP1, IP3 receptor, NF-kB p65/p65, TNF-alpha |
| 9 | Cell adhesion_Leucocyte chemotaxis | 180 | 7.200E-09 | 76 | Tubulin beta, Tubulin gamma, GRO-2, RhoA, RAP-1A, ITGB1, MIG, ROCK, CCL22, CD80, G-protein alpha-12 family, MHC class II, PI3K cat class IA, Tubulin alpha, Profilin, CCL17, Calmodulin, Slp76, CCR7, NCK1, GRB2, IP10, CCL13, GCP2, ROCK2, ITGB2, alpha-4/beta-1 integrin, c-Cbl, ICAM1, PTEN, CCL2, Vinculin, CCL14, PI3K cat class IA (p110-beta), Tubulin gamma 1, I-TAC, VASP, IL-8, N-WASP, G-protein alpha-i family, CX3CR1, DOCK2, CCR2, ROCK1, CXCL13, ARHGEF1 (p115RhoGEF), ICAM3, MIP-1-beta, G-protein beta/gamma, Galpha(i)-specific peptide GPCRs, WASP, Fyn, CCL19, Rac2, CCR1, Galpha(q)-specific peptide GPCRs, FYB1, CCBP2 (CCR9), IL8RA, LAT, ICAM5, SKAP55, LFA-3, GRO-3, Btk, G-protein alpha-i2, CD86, CXCR4, Paxillin, Cofilin, Rac1, IP3 receptor, CCL15, G-protein alpha-13, IL8RB, Tubulin (in microtubules) |
| 10 | Immune response_Innate immune response to RNA viral infection | 84 | 1.808E-08 | 43 | I-kB, IDO1, MxA, STAT2, WARS, IRAK4, iNOS, p38beta (MAPK11), IP10, TRAF3, JAK1, MyD88, PKR, MDA-5, I-TAC, IL-8, ISGF3, RNaseL, ADAR1, TRAF6, IKK-beta, IRAKM, RIPK1, c-Jun, TBK1, IRF7, IL-6, IRF9, IKK (cat), IRF3, TANK, TLR7, TLR8, eIF2S1, NF-kB, TAB2, IFN-alpha/beta receptor, IRF1, Caspase-8, TLR3, 2'-5'-oligoadenylate synthetase, STAT1, RLI |
| 54 | Inflammation_Inflammasome | 118 | 2.680E-03 | 41 | IL-1 beta, Caspase-5, I-kB, AP-1, IRAK4, p38beta (MAPK11), NF-kB1 (p105), ISG15, IL-18, MyD88, PKR, MDA-5, RIPK2, IKK-alpha, ISGF3, CARD5, TLR4, TRAF6, IKK-beta, RIPK1, TBK1, IRF7, Nod2 (CARD15), JNK(MAPK8-10), IKK (cat), IRF3, TLR7, TLR8, eIF2S1, NF-kB, TAB2, Pannexin-1, TNF-R1, Caspase-1, TLR3, p38 MAPK, Btk, eIF2S2, NFKBIA, CARD8, TNF-alpha |

**Supplementary Table S2. Enrichment analysis of the ten most significant process networks in the top one hundred upregulated genes in human bronchial epithelial cells (HBECs) from control individuals and patients with asthma after rhinovirus A16 infection. Analyzed from GSE61141[66].**

| HBECs from control individuals | | | | | |
|---|---|---|---|---|---|
| | **Networks** | **Total** | **p-value** | **In data** | **Network objects from active data** |
| 1 | Inflammation_Interferon signaling | 110 | 3.409E-37 | 27 | IL29, PKR, CCL5, IFI17, IRF1, IL28A, MxB, ISG20, TAP1 (PSF1), IFI44, GBP1, I-TAC, Apo-2L(TNFSF10), STAT1/STAT2, SOCS1, IDO1, ISG54, STAT1, IRF7, MxA, IFP 35, IFI56, ISG15, IL28B, TLR3, PML, STAT2 |
| 2 | Immune response_Innate immune response to RNA viral infection | 83 | 1.840E-16 | 14 | PKR, MDA-5, RIG-I, IP10, IRF1, I-TAC, WARS, 2'-5'-oligoadenylate synthetase, IDO1, STAT1, IRF7, MxA, TLR3, STAT2 |
| 3 | Inflammation_Jak-STAT Pathway | 185 | 4.596E-07 | 10 | IL29, CCL5, IL28A, PLAUR (uPAR), LIFR, STAT1/STAT2, SOCS1, STAT1, IL28B, STAT2 |
| 4 | Inflammation_IFN-gamma signaling | 109 | 9.450E-06 | 7 | PKR, CCL5, K12, IP10, IRF1, SOCS1, STAT1 |
| 5 | Inflammation_Inflammasome | 120 | 1.756E-04 | 6 | PKR, MDA-5, RIG-I, IRF7, ISG15, TLR3 |
| 6 | Chemotaxis | 139 | 2.765E-03 | 5 | CCL5, IP10, CX3CL1, PLAUR (uPAR), I-TAC |
| 7 | Inflammation_Innate inflammatory response | 181 | 8.446E-03 | 5 | APOBEC3G, IP10, I-TAC, IRF7, TLR3 |
| 8 | Cell adhesion_Leucocyte chemotaxis | 180 | 3.694E-02 | 4 | CCL5, IP10, CX3CL1, I-TAC |
| 9 | Proliferation_Negative regulation of cell proliferation | 183 | 3.889E-02 | 4 | PKR, IFI17, WARS, STAT1 |
| 10 | Immune response_Antigen presentation | 194 | 4.655E-02 | 4 | TAP1 (PSF1), STAT1, CEACAM1, STAT2 |

| HBECs from patients with asthma | | | | | |
|---|---|---|---|---|---|
| | **Networks** | **Total** | **p-value** | **In data** | **Network objects from active data** |
| 1 | Inflammation_Interferon signaling | 110 | 1.008E-32 | 25 | IL29, CCL5, IFI17, IL28A, MxB, ISG20, TAP1 (PSF1), IFI44, GBP1, I-TAC, Caspase-1, Apo-2L(TNFSF10), MIG, STAT1/STAT2, IDO1, ISG54, STAT1, IRF7, MxA, IFP 35, IFI56, IL28B, TLR3, PML, STAT2 |
| 2 | Immune response_Innate immune response to RNA viral infection | 83 | 1.227E-14 | 13 | MDA-5, RIG-I, IP10, IKK-epsilon, I-TAC, WARS, 2'-5'-oligoadenylate synthetase, IDO1, STAT1, IRF7, MxA, TLR3, STAT2 |
| 3 | Inflammation_Jak-STAT Pathway | 185 | 6.247E-06 | 9 | IL29, IL-15RA, CCL5, IL28A, LIFR, STAT1/STAT2, STAT1, IL28B, STAT2 |
| 4 | Inflammation_Inflammasome | 120 | 2.189E-04 | 6 | MDA-5, RIG-I, IKK-epsilon, Caspase-1, IRF7, TLR3 |
| 5 | Inflammation_IFN-gamma signaling | 109 | 1.126E-03 | 5 | CCL5, K12, IP10, MIG, STAT1 |
| 6 | Inflammation_Innate inflammatory response | 181 | 1.918E-03 | 6 | APOBEC3G, IP10, IKK-epsilon, I-TAC, IRF7, TLR3 |
| 7 | Chemotaxis | 139 | 3.287E-03 | 5 | CCL5, IP10, I-TAC, IL-16, MIG |
| 8 | Immune response_Th17-derived cytokines | 98 | 5.518E-03 | 4 | MMP-13, I-TAC, MIG, STAT1 |
| 9 | Inflammation_NK cell cytotoxicity | 163 | 3.063E-02 | 4 | IL-15RA, Apo-2L(TNFSF10), STAT1, STAT2 |
| 10 | Cell adhesion_Platelet-endothelium-leucocyte interactions | 174 | 3.063E-02 | 4 | MMP-13, CCL5, STAT1, CD68 |

**Supplementary Table S3. All antiviral response genes significantly changed after rhinovirus A16 (RV-A16) infection in bronchial brushings from control individuals and patients with asthma after in vivo RV-A16 infection or in differentiated human bronchial epithelial cells from control individuals and patients with asthma after in vitro RV-A16 infection (GSE61141)[66].**

| Bronchial brushings from control individuals in vivo | | |
|---|---|---|
| **Gene** | **p-value** | **$Log_2FC$** |
| LYST | 7.65E-05 | -0.443907 |
| IVNS1ABP | 1.08E-04 | -0.4197461 |
| MAPK11 | 3.59E-04 | 0.28467409 |
| BECN1 | 4.00E-04 | -0.3110455 |
| CCT5 | 4.43E-04 | -0.412418 |
| TBK1 | 4.93E-04 | -0.5302973 |
| ACTA2 | 5.67E-04 | -0.4198554 |
| ELMOD2 | 6.05E-04 | -0.4646418 |
| ABCE1 | 7.70E-04 | -0.4356744 |
| CD207 | 0.00120396 | 0.46706636 |
| DNAJC3 | 0.00121386 | -0.38085 |
| BNIP3 | 0.00162649 | -0.4523524 |
| UNC93B1 | 0.00165717 | 0.29162879 |
| SPACA3 | 0.00186858 | 0.29907682 |
| IKBKB | 0.00253497 | 0.31654719 |
| IL33 | 0.00257272 | -0.5662286 |
| IFNAR1 | 0.00277058 | -0.3379496 |
| SLFN11 | 0.00304737 | -0.5547823 |
| IRF3 | 0.00309228 | 0.26590699 |
| APOBEC3C | 0.00450924 | -0.4267123 |
| CHRM2 | 0.00499786 | 0.2406417 |
| IFNAR2 | 0.00502243 | -0.3416906 |
| CXADR | 0.00512015 | -0.2713458 |
| MAPK14 | 0.00614791 | -0.2494267 |
| HSPB1 | 0.00753452 | -0.2903693 |
| CXCR4 | 0.00848183 | -0.9033424 |
| KCNJ8 | 0.01022031 | 0.19106938 |
| CCDC130 | 0.01152673 | 0.26813823 |
| POLR3G | 0.01328435 | 0.29856694 |
| DDX1 | 0.01405911 | -0.2643058 |
| RELA | 0.01479098 | -0.2935714 |
| TRIM56 | 0.01547338 | 0.1881259 |
| TLR8 | 0.01552932 | -0.7104081 |
| PSMA2 | 0.01595628 | -0.4307298 |
| CHUK | 0.01748839 | -0.3588789 |
| FCN3 | 0.0184118 | 0.27715324 |
| IL12B | 0.0188253 | 0.208578 |
| POLR3F | 0.02210039 | -0.2253792 |
| HYAL2 | 0.02246593 | 0.1740213 |
| CFL1 | 0.02444936 | -0.3017413 |
| IFNGR2 | 0.02559222 | -0.3396424 |
| UNC93B1 | 0.02568665 | 0.28795446 |
| FGR | 0.02652297 | -0.7025369 |
| DDX21 | 0.02830701 | -0.3674024 |
| RNASEL | 0.02979535 | -0.1934832 |
| PTPRC | 0.03030047 | -0.8586604 |
| DHX36 | 0.03121596 | -0.3011299 |
| TNF | 0.0315715 | -0.5356796 |
| TPT1 | 0.0326018 | 0.45659592 |
| DDX3X | 0.033224 | -0.2460331 |
| XCL1 | 0.03392249 | 0.68704931 |
| MICA | 0.03469049 | 0.24172981 |
| BANF1 | 0.03533612 | 0.21109478 |
| MEF2C | 0.03647154 | -0.4306598 |
| CD86 | 0.03691275 | -0.8744841 |
| CYP1A1 | 0.03713228 | 0.25314154 |
| BANF1 | 0.03814064 | -0.2129742 |
| CCL4 | 0.04035564 | -0.703145 |
| CDK6 | 0.04161901 | -0.1921755 |
| PRKRA | 0.04265319 | -0.1935144 |
| IL6 | 0.044015 | -0.6248495 |

| | | |
|---|---|---|
| BNIP3L | 0.0445931 | -0.2107504 |
| GBP3 | 0.0463936 | -0.4441241 |

| Bronchial brushings from patients with asthma in vivo | | |
|---|---|---|
| **Gene** | **p-value** | **$Log_2FC$** |
| IFI44L | 2.7513864 | 2.44E-07 |
| IFITM1 | 2.26654724 | 8.93E-07 |
| IFI44 | 1.78485617 | 1.86E-06 |
| IFITM3 | 1.49792001 | 2.05E-06 |
| BNIP3 | -0.4879504 | 2.91E-06 |
| MX1 | 1.79778192 | 6.72E-06 |
| BST2 | 1.5213133 | 7.38E-06 |
| IRF9 | 0.5769468 | 8.53E-06 |
| OAS2 | 1.44133293 | 9.67E-06 |
| STAT1 | 1.26202938 | 1.22E-05 |
| OAS1 | 1.54613253 | 1.25E-05 |
| MX2 | 2.04970628 | 1.52E-05 |
| IFIT1 | 2.16478729 | 1.85E-05 |
| PLSCR1 | 1.12390665 | 2.00E-05 |
| DDX60 | 1.41443322 | 2.87E-05 |
| TRIM22 | 1.02773215 | 3.26E-05 |
| OAS3 | 1.54432653 | 5.23E-05 |
| IFIT3 | 1.81597424 | 5.49E-05 |
| DHX58 | 0.70446222 | 9.76E-05 |
| DDX58 | 1.47047202 | 9.79E-05 |
| EIF2AK | 0.7781584 | 1.71E-04 |
| FCN3 | -0.3050202 | 1.99E-04 |
| IKBKE | 0.36925583 | 2.21E-04 |
| STAT2 | 0.82594415 | 2.36E-04 |
| GPAM | -0.2662866 | 2.96E-04 |
| IFI16 | 0.66718059 | 3.25E-04 |
| ISG15 | 0.96431722 | 3.28E-04 |
| IFIT2 | 1.98331393 | 3.34E-04 |
| DUOX2 | 1.62111069 | 3.40E-04 |
| CXCL10 | 2.51578782 | 3.53E-04 |
| PSMB9 | 0.94885053 | 3.74E-04 |
| ISG20 | 0.59236245 | 4.44E-04 |
| ADAR | 0.52414375 | 4.48E-04 |
| RSAD2 | 1.58864248 | 4.60E-04 |
| NLRC5 | 0.61071381 | 6.45E-04 |
| TRIM5 | 0.68854983 | 6.60E-04 |
| IFITM2 | 0.87811185 | 7.01E-04 |
| IRF7 | 0.47976395 | 7.04E-04 |
| IRAK3 | 0.5721565 | 8.88E-04 |
| HERC5 | 1.15893187 | 0.00107361 |
| PML | 0.66965248 | 0.00129139 |
| CLU | -0.2599909 | 0.00156214 |
| IFIT5 | 0.78368937 | 0.0017862 |
| IFIH1 | 1.05426744 | 0.00181852 |
| CXCL9 | 1.63985158 | 0.00260229 |
| IRF1 | 0.72633033 | 0.00279675 |
| PMAIP1 | 0.78214465 | 0.003265 |
| GBP3 | 0.43999573 | 0.00331372 |
| PYCARD | 0.26990835 | 0.00334091 |
| ZNF175 | -0.2148014 | 0.00442761 |
| CCL22 | 0.23745083 | 0.00563918 |
| CCL19 | 0.24199143 | 0.0057422 |
| APOBEC3G | 0.59811982 | 0.00594614 |
| ZC3H12A | 0.27829253 | 0.00721255 |
| OASL | 0.89995485 | 0.00761047 |
| APOBEC3F | 0.34347153 | 0.0082971 |
| IL6 | 0.53328796 | 0.00912099 |
| ENO1 | 0.23331382 | 0.00934609 |
| ABCC9 | -0.2962732 | 0.00937097 |
| BATF3 | 0.20857068 | 0.00957246 |
| FOSL1 | 0.17740254 | 0.00977377 |
| LGALS9 | 0.49906318 | 0.009962 |
| HYAL1 | 0.15341526 | 0.0102313 |
| APOBEC3H | 0.21874676 | 0.01147115 |
| TLR3 | 0.47466227 | 0.01234421 |

| | | |
|---|---|---|
| SERINC3 | -0.1090338 | 0.01379487 |
| SAMHD1 | 0.25655864 | 0.0140242 |
| TLR8 | 0.46085583 | 0.01454437 |
| PTPRC | 0.62254589 | 0.01553325 |
| TRIM11 | 0.10106083 | 0.01588222 |
| CCL8 | 1.08815918 | 0.01736971 |
| BNIP3L | -0.1621839 | 0.01771446 |
| DCLK1 | -0.1757575 | 0.01996567 |
| LILRB1 | 0.63895374 | 0.02009985 |
| CXADR | -0.1408438 | 0.02034223 |
| F2RL1 | 0.20734859 | 0.02125084 |
| TICAM1 | 0.18010247 | 0.02131712 |
| APOBEC3D | 0.22884253 | 0.02143983 |
| IL33 | -0.2644045 | 0.02260751 |
| MST1R | 0.19118748 | 0.02307895 |
| HMGA1 | 0.13802169 | 0.02337107 |
| ATG7 | 0.19313255 | 0.02563914 |
| CDK6 | 0.13288052 | 0.02909091 |
| CD86 | 0.5879905 | 0.02938831 |
| HNRNPUL1 | -0.1108471 | 0.03039986 |
| CD207 | -0.188618 | 0.03084029 |
| IFNG | 0.62195628 | 0.03202974 |
| AGBL4 | -0.2163882 | 0.03219032 |
| IFNE | 0.16956428 | 0.03676611 |
| PRF1 | 0.4861052 | 0.04035228 |
| CD40 | 0.30119882 | 0.0404023 |
| ELMOD2 | -0.1612959 | 0.04506776 |
| SRC | 0.13429267 | 0.04968988 |

| **Human bronchial epithelial cells from control individuals in vitro** | | |
|---|---|---|
| **Gene** | **p-value** | **$Log_2FC$** |
| IFIT2 | 7.149E-19 | 6.556 |
| RSAD2 | 1.321E-18 | 5.049 |
| OASL | 3.391E-18 | 5.383 |
| DDX58 | 1.106E-17 | 4.451 |
| IFIT3 | 2.416E-17 | 5.408 |
| MX2 | 1.33E-16 | 4.293 |
| IFIH1 | 1.62E-16 | 3.93 |
| CXCL10 | 1.72E-16 | 7.211 |
| IFIT1 | 5.28E-16 | 5.359 |
| OAS3 | 1.49E-15 | 3.911 |
| OAS1 | 2.49E-15 | 3.849 |
| CCL5 | 7.93E-15 | 3.8 |
| HERC5 | 1.40E-14 | 3.05 |
| ISG15 | 1.77E-14 | 4.998 |
| DDX60 | 2.11E-14 | 3.328 |
| APOBEC3A | 2.28E-14 | 3.66 |
| MX1 | 3.55E-14 | 4.369 |
| C19orf66 | 6.97E-14 | 2.55 |
| TRIM22 | 1.05E-13 | 2.948 |
| IRF7 | 2.27E-13 | 2.788 |
| ISG20 | 2.99E-13 | 3.233 |
| OAS2 | 3.19E-13 | 3.4 |
| STAT1 | 5.49E-13 | 3.245 |
| IFI44L | 9.02E-13 | 3.161 |
| GBP1 | 1.03E-12 | 2.973 |
| LGALS9 | 1.14E-12 | 2.699 |
| NLRC5 | 1.88E-12 | 2.954 |
| IFITM1 | 2.24E-12 | 3.642 |
| STAT2 | 4.70E-12 | 2.322 |
| PML | 4.98E-12 | 2.81 |
| IFI44 | 5.01E-12 | 2.485 |
| TLR3 | 8.37E-12 | 2.098 |
| TRIM25 | 9.38E-12 | 2.307 |
| IRF1 | 9.84E-12 | 2.275 |
| EIF2AK2 | 1.68E-11 | 2.167 |
| APOBEC3G | 2.40E-11 | 2.338 |
| CXCL9 | 4.86E-11 | 3.759 |
| BST2 | 5.97E-11 | 2.546 |
| IFITM3 | 1.34E-10 | 2.711 |

| | | |
|---|---|---|
| DHX58 | 1.63E-10 | 2.137 |
| IFITM2 | 2.30E-10 | 3.114 |
| TRIM5 | 4.98E-10 | 2.133 |
| PLSCR1 | 5.15E-10 | 2.003 |
| TNF | 7.26E-10 | 1.634 |
| ZC3HAV1 | 2.38E-09 | 1.784 |
| PSMB9 | 2.94E-09 | 1.885 |
| TICAM1 | 4.18E-09 | 1.55 |
| DUOX2 | 8.29E-09 | 2.733 |
| APOBEC3F | 8.74E-09 | 1.714 |
| IKBKE | 1.91E-08 | 1.756 |
| IFIT5 | 2.91E-08 | 1.548 |
| IFI16 | 3.20E-08 | 1.766 |
| APOBEC3B | 7.72E-08 | 2.314 |
| CCL22 | 1.51E-07 | 1.697 |
| ADAR | 1.67E-07 | 1.552 |
| PMAIP1 | 2.48E-07 | 1.58 |
| IRF9 | 5.61E-07 | 1.231 |
| IL23A | 6.12E-07 | 1.446 |
| PSMA2 | 1.28E-06 | 1.173 |
| GBP3 | 3.34E-06 | 1.523 |
| AGBL5 | 1.41E-05 | -1.63 |
| BCL3 | 2.21E-05 | 1.001 |
| IRAK3 | 2.73E-05 | 1.001 |
| ZC3H12A | 5.23E-05 | 1.283 |
| TBK1 | 5.63E-05 | 0.781 |
| APOBEC3D | 7.83E-05 | 0.8212 |
| SRC | 0.000167 | 0.8428 |
| MST1R | 0.0002878 | 0.9134 |
| SAMHD1 | 0.0003501 | 0.8651 |
| UNC13D | 0.0003652 | 0.8952 |
| PIM2 | 0.0004463 | -0.7108 |
| IFNGR2 | 0.0004815 | 0.79 |
| HMGA1 | 0.0005404 | 0.8514 |
| FOSL1 | 0.000623 | 0.7987 |
| UNC93B1 | 0.0007337 | 1.011 |
| TRIM56 | 0.002255 | 0.5678 |
| APOBEC3C | 0.002863 | 0.6992 |
| GTF2F1 | 0.007381 | 0.5849 |
| MAVS | 0.007572 | -0.5188 |
| DHX36 | 0.00758 | 0.535 |
| RELA | 0.007593 | 0.5051 |
| IVNS1ABP | 0.01132 | -0.5954 |
| EIF2AK4 | 0.01289 | -0.4502 |
| HNRNPUL1 | 0.0142 | -0.5764 |
| STMN1 | 0.01626 | -0.4616 |
| TRIM34 | 0.01815 | 0.4168 |
| BCL2L1 | 0.01992 | -0.5224 |
| BNIP3 | 0.021 | -0.554 |
| IKBKG | 0.0211 | 0.4076 |
| CXADR | 0.02114 | -0.4977 |
| SERINC5 | 0.02395 | -0.4672 |
| TPT1 | 0.02416 | 0.53 |
| LSM14A | 0.03128 | -0.4094 |
| SLFN11 | 0.0375 | 0.4628 |
| ILF3 | 0.03774 | -0.4139 |
| DCLK1 | 0.03914 | -0.5191 |
| CD40 | 0.04965 | 0.412 |

| **Human bronchial epithelial cells from patients with asthma in vitro** | | |
|---|---|---|
| **Gene** | **p-value** | **$Log_2FC$** |
| OASL | 7.85E-24 | 5.906 |
| CXCL10 | 1.53E-23 | 8.047 |
| IFIT1 | 6.74E-22 | 5.723 |
| IFIT2 | 3.04E-20 | 7.015 |
| CXCL9 | 8.69E-20 | 4.486 |
| IFIH1 | 2.55E-19 | 4.319 |
| ISG20 | 7.46E-19 | 3.936 |
| DDX58 | 1.02E-18 | 4.937 |
| RSAD2 | 1.12E-18 | 5.618 |

| | | |
|---|---|---|
| HERC5 | 1.69E-18 | 3.359 |
| IFIT3 | 2.12E-18 | 5.715 |
| C19orf66 | 2.33E-18 | 3.042 |
| MX2 | 3.91E-18 | 4.787 |
| PML | 5.07E-18 | 3.02 |
| OAS1 | 6.75E-18 | 4.169 |
| DHX58 | 1.16E-17 | 2.529 |
| OAS2 | 1.38E-17 | 3.88 |
| NLRC5 | 1.54E-17 | 3.176 |
| CCL5 | 1.91E-17 | 4.491 |
| GBP1 | 2.11E-17 | 3.626 |
| MX1 | 2.96E-17 | 4.603 |
| STAT2 | 5.46E-17 | 2.636 |
| IFI44 | 5.47E-17 | 2.745 |
| TRIM22 | 5.79E-17 | 2.969 |
| APOBEC3G | 6.16E-17 | 2.688 |
| OAS3 | 6.39E-17 | 4.22 |
| APOBEC3A | 7.85E-17 | 4.408 |
| TRIM5 | 1.06E-16 | 2.389 |
| STAT1 | 1.50E-16 | 3.396 |
| IFI44L | 1.91E-16 | 3.4 |
| APOBEC3F | 2.71E-16 | 2.283 |
| DDX60 | 3.08E-16 | 3.367 |
| IRF7 | 3.30E-16 | 2.946 |
| IFITM1 | 6.70E-16 | 3.543 |
| IKBKE | 1.36E-15 | 2.104 |
| TLR3 | 1.49E-15 | 2.351 |
| LGALS9 | 2.32E-15 | 2.614 |
| PLSCR1 | 2.52E-15 | 2.191 |
| IRF1 | 5.70E-15 | 2.376 |
| IFITM2 | 8.08E-15 | 3.223 |
| EIF2AK2 | 1.37E-14 | 2.326 |
| ISG15 | 2.03E-14 | 5.013 |
| TRIM25 | 2.11E-14 | 2.397 |
| IFITM3 | 7.08E-14 | 2.617 |
| APOBEC3B | 2.09E-13 | 2.488 |
| TICAM1 | 5.63E-13 | 2.069 |
| PSMB9 | 5.84E-13 | 2 |
| ZC3HAV1 | 6.16E-13 | 2.085 |
| BST2 | 1.05E-12 | 2.952 |
| IFI16 | 2.85E-12 | 2.035 |
| TNF | 3.32E-12 | 2.265 |
| DUOX2 | 3.70E-12 | 3.288 |
| ADAR | 5.35E-12 | 1.839 |
| IFIT5 | 2.19E-11 | 1.601 |
| IRF9 | 3.05E-11 | 1.428 |
| PMAIP1 | 1.83E-10 | 1.51 |
| IL23A | 8.16E-10 | 2.161 |
| SAMHD1 | 1.19E-09 | 1.413 |
| ZC3H12A | 2.16E-09 | 1.723 |
| CCL22 | 4.67E-09 | 1.918 |
| MST1R | 2.12E-08 | 1.137 |
| BCL3 | 6.60E-08 | 1.057 |
| CD40 | 3.82E-07 | 0.9531 |
| SRC | 8.37E-07 | 0.8584 |
| IL6 | 1.11E-06 | 2.266 |
| TRIM34 | 1.49E-06 | 0.8456 |
| APOBEC3C | 3.56E-06 | 0.9637 |
| UNC93B1 | 4.02E-06 | 1.062 |
| IRAK3 | 1.09E-05 | 0.9916 |
| PSMA2 | 1.76E-05 | 0.8836 |
| HMGA1 | 2.06E-05 | 0.8408 |
| APOBEC3D | 3.79E-05 | 0.8388 |
| RELA | 4.69E-05 | 0.6723 |
| EIF2AK4 | 5.24E-05 | -0.734 |
| TRIM56 | 6.39E-05 | 0.6574 |
| ABCE1 | 8.10E-05 | -0.7558 |
| AGBL5 | 0.0001097 | -0.926 |
| IFNGR2 | 0.0001243 | 0.6204 |
| RNASEL | 0.0001512 | 0.6969 |

| | | |
|---|---|---|
| FOSL1 | 0.0001995 | 1.182 |
| GBP3 | 0.0003406 | 1.763 |
| UNC13D | 0.0003743 | 0.6806 |
| BANF1 | 0.0005562 | -0.618 |
| SERINC5 | 0.001265 | -0.5607 |
| POLR3E | 0.001463 | -0.5625 |
| GTF2F1 | 0.00148 | 0.5035 |
| DCLK1 | 0.001586 | -0.7357 |
| STMN1 | 0.001603 | -0.6112 |
| CXADR | 0.001645 | -0.5167 |
| BCL2L1 | 0.002159 | -0.5074 |
| DDX3X | 0.002533 | 0.4623 |
| IRF5 | 0.002761 | 0.5806 |
| BAD | 0.004124 | -0.543 |
| MAVS | 0.00597 | -0.4032 |
| TBK1 | 0.006174 | 0.4936 |
| FAM111A | 0.009602 | 0.4343 |
| LSM14A | 0.01183 | -0.4332 |
| IKBKG | 0.01247 | 0.3916 |
| ILF3 | 0.01367 | -0.3934 |
| SPON2 | 0.01699 | 0.53 |
| DDX21 | 0.0178 | -0.3879 |
| IFNAR1 | 0.01898 | -0.3322 |
| IVNS1ABP | 0.02507 | -0.3771 |
| XPR1 | 0.02681 | 0.4449 |
| ELMOD2 | 0.02688 | -0.4107 |
| PYCARD | 0.03129 | 0.3599 |
| LYST | 0.03498 | 0.351 |
| HNRNPUL1 | 0.03777 | -0.3541 |
| PRKRA | 0.0379 | -0.3664 |
| BNIP3L | 0.04307 | -0.319 |
| DHX36 | 0.0456 | 0.3483 |
| TMEM173 | 0.04837 | 0.3204 |

**Supplementary Table S4. KEGG pathways enriched in significantly changed proteins secreted from the human bronchial epithelial cells of control subjects and patients with asthma after house dust mite stimulation and rhinovirus A16 (RV-A16) infection, as compared to RV-A16 infection alone.**

| KEGG pathways downregulated (controls) | | | | | | |
|---|---|---|---|---|---|---|
| #term ID | term description | observed gene count | background gene count | strength | false discovery rate | matching proteins in your network (labels) |
| map04060 | Cytokine-cytokine receptor interaction | 21 | 143 | 0.79 | 0.0000000000139 | CX3CL1, TGFB1, CCL2, CXCL6, KITLG, IL12B, TNFSF10, IL18, CXCL5, CXCL11, CCL20, CCL28, CCL7, CCL8, IL6, TNF, TNFSF14, FLT3LG, CCL3, VEGFA, CCL4 |
| map04062 | Chemokine signaling pathway | 11 | 40 | 1.06 | 0.000000202 | CX3CL1, CCL2, CXCL6, CXCL5, CXCL11, CCL20, CCL28, CCL7, CCL8, CCL3, CCL4 |
| map05323 | Rheumatoid arthritis | 10 | 36 | 1.06 | 0.000000784 | TGFB1, CCL2, CXCL6, IL18, CXCL5, CCL20, IL6, TNF, CCL3, VEGFA |
| map04620 | Toll-like receptor signaling pathway | 7 | 22 | 1.12 | 0.0000553 | IL12B, CXCL11, CASP8, IL6, TNF, CCL3, CCL4 |
| map04657 | IL-17 signaling pathway | 8 | 36 | 0.97 | 0.0000725 | CCL2, CXCL6, CXCL5, CASP8, CCL20, CCL7, IL6, TNF |
| map05142 | Chagas disease (American trypanosomiasis) | 7 | 28 | 1.02 | 0.00014 | TGFB1, CCL2, IL12B, CASP8, IL6, TNF, CCL3 |
| map04668 | TNF signaling pathway | 7 | 31 | 0.97 | 0.00022 | CX3CL1, CCL2, CXCL5, CASP8, CCL20, IL6, TNF |
| map05134 | Legionellosis | 5 | 12 | 1.24 | 0.00036 | IL12B, IL18, CASP8, IL6, TNF |
| map05200 | Pathways in cancer | 10 | 98 | 0.63 | 0.001 | TGFB1, KITLG, IL12B, AXIN1, TGFA, FGF5, CASP8, IL6, FLT3LG, VEGFA |
| map05168 | Herpes simplex infection | 6 | 31 | 0.91 | 0.0015 | CCL2, IL12B, CASP8, IL6, TNF, TNFSF14 |
| map05164 | Influenza A | 6 | 33 | 0.88 | 0.0019 | CCL2, IL12B, TNFSF10, IL18, IL6, TNF |
| map04621 | NOD-like receptor signaling pathway | 5 | 21 | 1 | 0.0022 | CCL2, IL18, CASP8, IL6, TNF |
| map05133 | Pertussis | 5 | 21 | 1 | 0.0022 | CXCL6, IL12B, CXCL5, IL6, TNF |
| map05152 | Tuberculosis | 6 | 36 | 0.84 | 0.0023 | TGFB1, IL12B, IL18, CASP8, IL6, TNF |
| map05321 | Inflammatory bowel disease (IBD) | 5 | 23 | 0.96 | 0.0025 | TGFB1, IL12B, IL18, IL6, TNF |
| map04933 | AGE-RAGE signaling pathway in diabetic complications | 5 | 25 | 0.92 | 0.0033 | TGFB1, CCL2, IL6, TNF, VEGFA |
| map05144 | Malaria | 5 | 26 | 0.9 | 0.0037 | TGFB1, CCL2, IL18, IL6, TNF |
| map05132 | Salmonella infection | 4 | 14 | 1.08 | 0.0038 | IL18, IL6, CCL3, CCL4 |
| map05143 | African trypanosomiasis | 4 | 15 | 1.05 | 0.0044 | IL12B, IL18, IL6, TNF |
| map04010 | MAPK signaling pathway | 7 | 71 | 0.61 | 0.0086 | TGFB1, KITLG, TGFA, FGF5, TNF, FLT3LG, VEGFA |
| map04932 | Non-alcoholic fatty liver disease (NAFLD) | 4 | 19 | 0.94 | 0.0086 | TGFB1, CASP8, IL6, TNF |
| map05161 | Hepatitis B | 4 | 20 | 0.92 | 0.0097 | TGFB1, CASP8, IL6, TNF |
| map04623 | Cytosolic DNA-sensing pathway | 3 | 9 | 1.14 | 0.0109 | IL18, IL6, CCL4 |
| map05146 | Amoebiasis | 4 | 22 | 0.88 | 0.0121 | TGFB1, IL12B, IL6, TNF |
| map05145 | Toxoplasmosis | 4 | 24 | 0.84 | 0.0154 | TGFB1, IL12B, CASP8, TNF |
| map04622 | RIG-I-like receptor signaling pathway | 3 | 11 | 1.06 | 0.0155 | IL12B, CASP8, TNF |
| map05212 | Pancreatic cancer | 3 | 11 | 1.06 | 0.0155 | TGFB1, TGFA, VEGFA |
| map01521 | EGF5R tyrosine kinas5e inhibitor resistance | 4 | 26 | 0.81 | 0.0177 | TGFA, EIF4EBP1, IL6, VEGFA |
| map05211 | Renal cell carcinoma | 3 | 12 | 1.02 | 0.0177 | TGFB1, TGFA, VEGFA |
| map04014 | Ras signaling pathway | 5 | 45 | 0.67 | 0.0189 | KITLG, TGFA, FGF5, FLT3LG, VEGFA |

| | | | | | | |
|---|---|---|---|---|---|---|
| map05165 | Human papillomavirus infection | 5 | 46 | 0.66 | 0.02 | AXIN1, EIF4EBP1, CASP8, TNF, VEGFA |
| map04151 | PI3K-Akt signaling pathway | 7 | 92 | 0.5 | 0.0215 | KITLG, TGFA, FGF5, EIF4EBP1, IL6, FLT3LG, VEGFA |
| map05210 | Colorectal cancer | 3 | 14 | 0.95 | 0.0218 | TGFB1, AXIN1, TGFA |
| map05225 | Hepatocellular carcinoma | 3 | 14 | 0.95 | 0.0218 | TGFB1, AXIN1, TGFA |
| map05410 | Hypertrophic cardiomyopathy (HCM) | 3 | 14 | 0.95 | 0.0218 | TGFB1, IL6, TNF |
| map04068 | FoxO signaling pathway | 3 | 17 | 0.87 | 0.0319 | TGFB1, TNFSF10, IL6 |
| map04218 | Cellular senescence | 3 | 19 | 0.82 | 0.0407 | TGFB1, EIF4EBP1, IL6 |
| map01523 | Antifolate resistance | 2 | 6 | 1.14 | 0.0408 | IL6, TNF |
| map04672 | Intestinal immune network for IgA production | 3 | 21 | 0.77 | 0.0492 | TGFB1, CCL28, IL6 |
| map05140 | Leishmaniasis | 3 | 21 | 0.77 | 0.0492 | TGFB1, IL12B, TNF |
| map05226 | Gastric cancer | 3 | 21 | 0.77 | 0.0492 | TGFB1, AXIN1, FGF5 |

Pathways marked in red demonstrate proteins included in viral infection pathway on the Figure 7C

| **KEGG pathways upregulated (controls)** |
|---|
| No significant enrichment |

| **KEGG pathways downregulated (asthma)** | | | | | | |
|---|---|---|---|---|---|---|
| **#term ID** | **term description** | **observed gene count** | **background gene count** | **strength** | **false discovery rate** | **matching proteins in your network (labels)** |
| map04060 | Cytokine-cytokine receptor interaction | 21 | 143 | 0.79 | 0.0000000000132 | CX3CL1, TGFB1, CCL2, CXCL6, KITLG, TNFSF10, IL17C, IL7, CXCL5, CXCL10, CXCL11, CCL20, CCL28, CCL7, IL6, TNF, TNFSF14, FLT3LG, CCL3, VEGFA, CCL4 |
| map04062 | Chemokine signaling pathway | 11 | 40 | 1.06 | 0.000000192 | CX3CL1, CCL2, CXCL6, CXCL5, CXCL10, CXCL11, CCL20, CCL28, CCL7, CCL3, CCL4 |
| map04657 | IL-17 signaling pathway | 10 | 36 | 1.06 | 0.000000742 | CCL2, CXCL6, IL17C, CXCL5, CXCL10, MMP1, CCL20, CCL7, IL6, TNF |
| map05323 | Rheumatoid arthritis | 10 | 36 | 1.06 | 0.000000742 | TGFB1, CCL2, CXCL6, CXCL5, MMP1, CCL20, IL6, TNF, CCL3, VEGFA |
| map04668 | TNF signaling pathway | 7 | 31 | 0.97 | 0.00029 | CX3CL1, CCL2, CXCL5, CXCL10, CCL20, IL6, TNF |
| map04620 | Toll-like receptor signaling pathway | 6 | 22 | 1.06 | 0.00045 | CXCL10, CXCL11, IL6, TNF, CCL3, CCL4 |
| map05200 | Pathways in cancer | 10 | 98 | 0.63 | 0.0012 | TGFB1, KITLG, AXIN1, IL7, TGFA, FGF5, MMP1, IL6, FLT3LG, VEGFA |
| map04933 | AGE-RAGE signaling pathway in diabetic complications | 5 | 25 | 0.92 | 0.0063 | TGFB1, CCL2, IL6, TNF, VEGFA |
| map05142 | Chagas disease (American trypanosomiasis) | 5 | 28 | 0.87 | 0.0088 | TGFB1, CCL2, IL6, TNF, CCL3 |
| map04010 | MAPK signaling pathway | 7 | 71 | 0.61 | 0.015 | TGFB1, KITLG, TGFA, FGF5, TNF, FLT3LG, VEGFA |
| map04151 | PI3K-Akt signaling pathway | 8 | 92 | 0.56 | 0.015 | KITLG, IL7, TGFA, FGF5, EIF4EBP1, IL6, FLT3LG, VEGFA |
| map05164 | Influenza A | 5 | 33 | 0.8 | 0.015 | CCL2, TNFSF10, CXCL10, IL6, TNF |
| map04623 | Cytosolic DNA-sensing pathway | 3 | 9 | 1.14 | 0.0181 | CXCL10, IL6, CCL4 |
| map05133 | Pertussis | 4 | 21 | 0.9 | 0.0181 | CXCL6, CXCL5, IL6, TNF |
| map04640 | Hematopoietic cell lineage | 5 | 41 | 0.71 | 0.0248 | KITLG, IL7, IL6, TNF, FLT3LG |
| map05212 | Pancreatic cancer | 3 | 11 | 1.06 | 0.0248 | TGFB1, TGFA, VEGFA |
| map01521 | EGFR tyrosine kinase inhibitor resistance | 4 | 26 | 0.81 | 0.0277 | TGFA, EIF4EBP1, IL6, VEGFA |
| map04014 | Ras signaling pathway | 5 | 45 | 0.67 | 0.0277 | KITLG, TGFA, FGF5, FLT3LG, VEGFA |

| | | | | | | |
|---|---|---|---|---|---|---|
| map04926 | Relaxin signaling pathway | 3 | 13 | 0.98 | 0.0277 | TGFB1, MMP1, VEGFA |
| map05144 | Malaria | 4 | 26 | 0.81 | 0.0277 | TGFB1, CCL2, IL6, TNF |
| map05211 | Renal cell carcinoma | 3 | 12 | 1.02 | 0.0277 | TGFB1, CCL2, IL6, TNF |
| map05132 | Salmonella infection | 3 | 14 | 0.95 | 0.031 | IL6, CCL3, CCL4 |
| map05210 | Colorectal cancer | 3 | 14 | 0.95 | 0.031 | TGFB1, AXIN1, TGFA |
| map05225 | Hepatocellular carcinoma | 3 | 14 | 0.95 | 0.031 | TGFB1, AXIN1, TGFA |
| map05410 | Hypertrophic cardiomyopathy (HCM) | 3 | 14 | 0.95 | 0.031 | TGFB1, IL6, TNF |
| map05168 | Herpes simplex infection | 4 | 31 | 0.73 | 0.0319 | CCL2, IL6, TNF, TNFSF14 |
| map04068 | FoxO signaling pathway | 3 | 17 | 0.87 | 0.0403 | TGFB1, TNFSF10, IL6 |

Pathways marked in red demonstrate proteins included in viral infection pathway on the Figure 7C

| KEGG pathways upregulated (asthma) | | | | | | |
|---|---|---|---|---|---|---|
| **#term ID** | **term description** | **observed gene count** | **background gene count** | **strength** | **false discovery rate** | **matching proteins in your network (labels)** |
| map04060 | Cytokine-cytokine receptor interaction | 8 | 143 | 0.7 | 0.0021 | IL4, IL1A, IL10RB, TNFRSF11B, CD40, IL24, TNFSF11, TNFRSF9 |

Pathways marked in red demonstrate proteins included in cytokine-mediated signaling pathway on the Figure 7C

**Supplementary Table S5. Protein concentrations in apical compartment of human bronchial epithelial cells from control individuals and patients with asthma in indicated conditions.**

| **Control (n=4)** | **Medium** | **RV-A16** | **SARS-CoV-2** | **RV-A16 + SARS-CoV-2** | **HDM** | **HDM+ RV-A16** | **HDM + SARS-CoV-2** | **HDM+RV-A16+ SARS-CoV-2** |
|---|---|---|---|---|---|---|---|---|
| **TWEAK**, pg/mL (mean, SEM) | 70.61 (10.11) | 81.12 (25.90) | 91.16 (30.74) | 103.3 (24.18) | 106.9 (31.01) | 72.73(17.71) | 83.42 (24.71) | 58.33 (10.82) |
| **IL-7**, pg/mL (mean, SEM) | 11.67 (4.517) | 11.68 (0.4573) | 12.33 (2.145) | 20.02 (9.446) | 18.29 (8.381) | 13.35 (4.343) | 17.52 (7.691) | 12.91 (3.969) |
| **IL-6**, pg/mL (mean, SEM) | 838.9 (461.0) | 1603 (761.4) | 895.8 (503.8) | 1542 (837.9) | 411.5 (182.0) | 637.9 (395.5) | 447.2 (271.5) | 642.7 (332.4) |
| **TRAIL**, pg/mL (mean, SEM) | 1246 (279.0) | 1879 (276.4) | 1089 (439.8) | 2416 (923.1) | 806.8 (162.3) | 1012 (253.8) | 1106 (442.6) | 1122 (306.4) |
| **IL-18**, pg/mL (mean, SEM) | 1.13 (0.09892) | 4.767 (0.4784) | 2.427 (0.3510) | 6.238 (1.634) | 2.261 (0.9410) | 5.52 (0.4008) | 3.224 (1.174) | 6.117 (1.489) |
| **IL-15**, pg/mL (mean, SEM) | 13.64 (3.601) | 14.61 (2.332) | 11.81 (3.049) | 18.45 (4.596) | 10.66 (1.038) | 13.91 (2.131) | 14.02 (2.787) | 15.36 (2.302) |
| **CXCL9**, pg/mL (mean, SEM) | 26.57 (5.050) | 75.36 (5.172) | 26.41 (5.236) | 64.17 (12.07) | 29.64 (6.358) | 68.07 (26.22) | 28.67 (4.519) | 70.48 (34.03) |
| **IL-17C**, pg/mL (mean, SEM) | 57.13 (14.14) | 131.7 (65.29) | 63.66 (35.79) | 91.39 (30.62) | 75.52 (30.22) | 75.64 (25.73) | 47.46 (8.440) | 57.80 (8.351) |
| **CCL8**, pg/mL (mean, SEM) | 0.2323 (0.02910) | 5.146 (2.076) | 0.4479 (0.1871) | 3.085 (1.220) | 0.3860 (0.1017) | 2.052 (1.083) | 0.4622 (0.2222) | 2.112 (0.5764) |

*RV-A16*, rhinovirus A16; *SARS-CoV-2*, severe acute respiratory syndrome coronavirus 2; *HDM*, house dust mite

| **Asthma (n=5)** | **Medium** | **RV-A16** | **SARS-CoV-2** | **RV-A16 + SARS-CoV-2** | **HDM** | **HDM+ RV-A16** | **HDM + SARS-CoV-2** | **HDM+RV-A16+ SARS-CoV-2** |
|---|---|---|---|---|---|---|---|---|
| **TWEAK**, pg/mL (mean, SEM) | 59.38 (16.12) | 99.62 (35.25) | 62.92 (13.09) | 43.90 (7.524) | 148.0 (57.71) | 108.3 (35.22) | 143.5 (60.21) | 148.7 (55.08) |
| **IL-7**, pg/mL (mean, SEM) | 13.36 (3.353) | 21.49 (9.893) | 14.12 (3.772) | 9.822 (2.986) | 27.67 (14.72) | 20.58 (9.203) | 24.80 (14.36) | 29.50 (15.03) |
| **IL-6**, pg/mL (mean, SEM) | 1004 (323.7) | 1795 (558.7) | 959.6 (280.9) | 1147 (392.5) | 658.3 (255.2) | 953.3 (406.1) | 625.9 (250.9) | 1558 (604.9) |
| **TRAIL**, pg/mL (mean, SEM) | 724.6 (59.55) | 2069 (520.7) | 945.7 (179.2) | 1144 (238.4) | 1153 (360.4) | 1266 (136.6) | 941.0 (256.9) | 2167 (177.7) |
| **IL-18**, pg/mL (mean, SEM) | 1.603 (0.2176) | 7.817 (1.117) | 2.076 (0.2509) | 10.25 (3.334) | 2.336 (0.6292) | 9.263 (2.571) | 2.825 (0.3171) | 9.956 (2.112) |
| **IL-15**, pg/mL (mean, SEM) | 10.32 (1.243) | 16.20 (1.749) | 11.94 (1.113) | 15.80 (1.542) | 7.976 (1.722) | 18.68 (3.937) | 11.13 (1.307) | 21.62 (1.733) |
| **CXCL9**, pg/mL (mean, SEM) | 22.10 (7.700) | 53.18 (12.46) | 20.93 (5.088) | 35.83 (10.81) | 19.97 (6.450) | 64.72 (27.70) | 17.19 (4.945) | 67.16 (26.03) |
| **IL-17C**, pg/mL (mean, SEM) | 53.18 (26.43) | 196.5 (103.5) | 45.89 (17.83) | 94.16 (28.81) | 66.09 (25.44) | 156.8 (74.68) | 64.77 (30.70) | 195.0 (110.2) |
| **CCL8**, pg/mL (mean, SEM) | 0.4266 (0.1867) | 6.139 (4.612) | 0.3114 (0.1229) | 1.687 (0.5324) | 0.4799 (0.1937) | 6.693 (5.944) | 0.3721 (0.1516) | 14.40 (12.91) |

*RV-A16*, rhinovirus A16; *SARS-CoV-2*, severe acute respiratory syndrome coronavirus 2; *HDM*, house dust mite

**Supplementary Table S6. House dust mite extracts characteristics.**

| | **HDM extract (Allergopharma)** | **HDM extract B (Citeq)** |
|---|---|---|
| Protein content | 0.525 mg protein/mg extract | 0.216 mg protein/ mg extract |
| Endotoxin content | 6.27 EU/mg extract | 51 EU/mg extract |
| Der p 1 | 46.66 µg/mg extract | 21 µg/mg extract |
| Der p 2 | - | 2.9 µg/mg extract |

*EU*, endotoxin units, *Der p 1*, house dust mite Der p1 allergen; *Der p 2*, house dust Mite Der p 2 allergen.

**Supplementary Table S7. Commercially available reagents used in the study.**

| Reagent | Catalog number | Company |
|---|---|---|
| Human Bronchial Epithelial cells | EP51AB | Epithelix |
| Human Bronchial Epithelial cells | CC-2540 | Lonza |
| THP-1 cells | thpx-sp | Invivogen |
| House dust mite extract | 02.01.64 | Citeq |
| Human Rhinovirus A16 | C1404B | Virapur |
| Ac-YVAD-cmk caspase-1 inhibitor | inh-yvad | Invivogen |
| MCC950, NLRP3 inflammasome inhibitor | AV02509 | Avistron |
| TBK1/IKKε inhibitor BX795 | 702675-74-9 | Sigma-Aldrich |
| LPS | tlrl-3pelps | Invivogen |
| ATP | tlrl-atp | Invivogen |
| BEGM Bronchial Epithelial Growth Medium BulletKit | CC3171+CC-4175 | Lonza |
| DMEM | 41965-039 | Gibco |
| MucilAir Medium | EP03MD | Epithelix |
| RPMI-1640 | R8758-500ml | Sigma-Aldrich |
| Opti-MEM® I Reduced Serum Medium | 31985070 | LifeTechnologies |
| Corning Inserts in 24-well culture plates, sterile | 3470-COR | Corning |
| Retinoic Acid | R2625-50mg | Sigma-Aldrich |
| Trypsin-EDTA (0.5%) | 15400-054 | Thermofisher Scientific |
| Human IL1B/IL-1F2 duo set | DY201 | R&D systems |
| TMB Substrate Reagent Kit | 555214 | BD |
| V-Plex Human IL1B kit | K151QPD-1 | MSD |
| RIPA Lysis and Extraction Buffer | 89901 | ThermoFisher Scientific |
| cOmplete™, Mini, EDTA-free Protease Inhibitor Cocktail | 4693159001 | Merc (Roche) |
| Pierce BCA Protein Assay Kit | 23225 | ThermoFisher Scientific |
| 4–20% Mini-PROTEAN® TGX™ Precast Protein Gels, 10-well, 50 µl | 4561094 | Biorad |
| Trans-Blot® Turbo™ Mini Nitrocellulose Transfer Packs | 1704158 | Biorad |
| 10x Tris/Glycine/SDS Running Buffer | 161-0772 | Biorad |
| SurePAGE™, Bis-Tris, 10x8, 4-20%, 12 wells | M00656 | GenScript |
| Nitrocellulose Transfer Membrane, 0.22um | L-08006-001 | Advansta |
| WesternBright Sirius HRP substrate | K-12043-C20 | Advansta |
| Restore™ PLUS Western Blot Stripping Buffer | 46428 | ThermoFisher Scientific |
| SuperSignal West Femto Maximum Sensitivity Substrate | 34095 | ThermoFisher Scientific |
| 2-mercaptoetanol | 63689-100ml-F | Sigma-Aldrich |
| RNAeasy Plus Micro Kit | 74034 | Qiagen |
| RevertAid RT Reverse Transcription Kit | K1691 | ThermoFisher Scientific |
| Maxima SYBR Green/ROX qPCR Master Mix (2X) | K0221 | ThermoFisher Scientific |
| RNAlater Stabilization Solution | 1018087 | Qiagen |
| SuperScript IV VILO Master Mix | 11756050 | ThermoFisher Scientific |
| RecoverAll | AM1975 | ThermoFisher Scientific |
| FSC22 Frozen Section Media | 3801480 | Leica |
| ProLong Diamond Antifade Mountant with DAPI | P36962 | ThermoFisher Scientific |
| TaqMan™ 2019-nCoV Assay Kit v1* | A47532 | ThermoFisher Scientific |
| RV_20f2 TaqMan Assay - Vi99990017_po | A41333 | ThermoFisher Scientific |
| **Western Blot analyses** | | |
| Mouse anti-NLRP3 | AG-20B-0014-C100 | Adipogen |
| Goat anti-IL1B | AF-201-NA | R&D systems |
| Mouse anti-ASC | sc-514414 | Santa Cruz Biotechnology |
| Rabbit anti-Caspase-1 | 2225 | Cell Signaling |
| Goat anti-RIG-I | sc-48929 | Santa Cruz Biotechnology |

| | | |
|---|---|---|
| HRP Goat Anti Mouse IgG | 111-035-146 | Jackson Laboratory |
| HRP Mouse anti-goat IgG | sc2354 | Santa Cruz Biotechnology |
| HRP AffiniPure Goat Anti Rabbit IgG | 111-035-003 | Jackson Laboratory |
| HRP Anti-beta Actin | ab49900 | Abcam |
| **Co-immunoprecipitation analyses** | | |
| Rabbit anti-ASC | sc22514-R | Santa Cruz Biotechnology |
| Protein A Beads | 161-0413 | Biorad |
| Mouse anti-RIG-I | sc376845 | Santa Cruz Biotechnology |
| Rabbit anti-MDA5 | ab126630 | Abcam |
| HRP Goat Anti Mouse IgG | 115-035-146 | Jackson Laboratory |
| HRP AffiniPure Goat Anti Rabbit IgG | 111-035-003 | Jackson Laboratory |
| **Western Blot from apical compartments analyses** | | |
| Goat anti-IL1B | AF-201-NA | R&D systems |
| HRP Mouse anti-goat IgG | sc2354 | Santa Cruz Biotechnology |
| **Confocal staining in HBECs** | | |
| Mouse IgG1 anti-IL1B | ab156791 | Abcam |
| Mouse IgG1 anti-RIG-I | sc-376845 | Santa Cruz Biotechnology |
| Mouse IgG1 anti-ASC | sc-514414 | Santa Cruz Biotechnology |
| Goat anti-mouse IgG, Alexa Fluor 546 | A11003 | Invitrogen |
| Goat anti-mouse IgG, Alexa Fluor 488 | A11001 | Invitrogen |
| Mouse IgG1 Control | X0931 | Dako |
| Goat serum (Normal) | X0907 | Dako |
| **Confocal staining in bronchial brushings** | | |
| Rabbit anti-Caspase-1 | 2225 | Cell Signalling |
| Mouse IgG1 anti-IL1B | ab156791 | Abcam |
| Mouse IgG1 anti-RIG-I | sc-376845 | Santa Cruz Biotechnology |
| Goat anti-Rabbit IgG, Alexa Fluor 488 | A11034 | Invitrogen |
| Goat anti-mouse IgG, Alexa Fluor 546 | A11003 | Invitrogen |
| Goat anti-mouse IgG, Alexa Fluor 546 | A11003 | Invitrogen |
| Rabbit Immunoglobin Fraction | X0936 | Dako |
| Mouse IgG1 Control | X0931 | Dako |
| Goat Serum (Normal) | X0907 | Dako |
| **Confocal staining for NLRP3 and SARS-CoV-2 experiment** | | |
| Anti ACE2 | ab15348 | Abcam |
| Anti NLRP3 | AG-20B-0014-C100 | Adipogen |
| Anti Occludin | OC-3F10 | ThermoFisher |
| Anti N-Protein | MA1-7404 | ThermoFisher |
| Rabbit Ig Fraction | X0936 | DAKO |
| Mouse IgG1 | X0931 | DAKO |
| Mouse IgG2b | X0944 | DAKO |
| Goat anti-Mouse IgG Alexa 488 | A11001 | Invitrogen |
| Goat anti-Rabbit IgG Alexa 546 | A11010 | Invitrogen |
| Goat anti mouse IgG2b | A21143 | Invitrogen |
| DAPI | 10236276001 | Sigma Aldrich |

**Supplementary Table S8. Clinical characteristics of study participants. In vivo and in vitro cohorts.**

| In vivo RV-A16 infection cohort | | | | | | | | | |
|---|---|---|---|---|---|---|---|---|---|
| **Condition** | **Gina status** | **ACQ status at baseline$** | **Age (years, age range)** | **Gender** | **Drugs** | **FeV1 (%) day 0 (% predicted)** | **Total serum IgE (IU/mL)** | **Total SPT weal size (mm)** | **HDM SPT weal size (mm)** |
| Control | Healthy | Healthy | 21-25 | Male | na | 113 | 22 | 0 | 0 |
| Control | Healthy | Healthy | 21-25 | Male | na | 97 | 16 | 0 | 0 |
| Control | Healthy | Healthy | 21-25 | Male | na | 97 | 16 | 0 | 0 |
| Control | Healthy | Healthy | 46-50 | Female | na | 88 | 19 | 0 | 0 |
| Control | Healthy | Healthy | 31-35 | Female | na | 103 | 13 | 0 | 0 |
| Control | Healthy | Healthy | 26-30 | Male | na | 105 | 3 | 0 | 0 |
| Control | Healthy | Healthy | 26-30 | Female | na | 105 | 38 | 0 | 0 |
| Control | Healthy | Healthy | 51-55 | Male | na | 105 | 16 | 0 | 0 |
| Control | Healthy | Healthy | 16-20 | Male | na | 100 | 14 | 0 | 0 |
| Asthma | mild | well | 26-30 | Female | SABA | 113 | 207 | 5 | 0 |
| Asthma | moderate | poor | 31-35 | Male | ICS | 70 | 870 | 11 | 3 |
| Asthma | mild | well | 21-25 | Male | SABA | 91 | 121 | 14 | 4 |
| Asthma | moderate | well | 16-20 | Female | ICS | 92 | 64 | 16 | 0 |
| Asthma | moderate | well | 46-50 | Female | ICS | 88 | 57 | 8 | 0 |
| Asthma | mild | partial | 31-35 | Male | SABA | 79 | 806 | 13 | 5 |
| Asthma | moderate | partial | 31-35 | Female | ICS | 65 | 507 | 8 | 0 |
| Asthma | moderate | poor | 31-35 | Male | ICS | 84 | 146 | 12 | 3 |
| Asthma | moderate | partial | 46-50 | Male | SABA | 73 | 1204 | 7 | 4 |
| Asthma | mild | partial | 31-35 | Female | SABA | 78 | 119 | 6 | 6 |
| Asthma | moderate | poor | 26-30 | Male | SABA | 73 | 19 | 11 | 4 |
| Asthma | moderate | poor | 51-55 | Male | ICS | 83 | 157 | 26 | 5 |
| Asthma | moderate | partial | 31-35 | Female | ICS | 77 | 67 | 7 | 0 |
| Asthma | moderate | poor | 41-45 | Female | ICS | 68 | 1593 | 3 | 4 |
| Asthma | mild | well | 21-25 | Female | SABA | 115 | 213 | 10 | 4 |
| Asthma | moderate | poor | 41-45 | Male | ICS | 74 | 228 | 17 | 0 |
| Asthma | moderate | well | 46-50 | Male | ICS | 81 | 87 | 9 | 3 |
| Asthma | moderate | partial | 46-50 | Female | ICS | 82 | 2106 | 29 | 9 |
| Asthma | mild | well | 51-55 | female | SABA | 101 | 69 | 14 | 5 |

*RV-A16;* rhinovirus A16*; GINA*; global initiative for asthma; *ACQ*, asthma control questionnaire; *FeV1*, forced expiratory volume in 1 second; *IgE*, immunoglobulin E; *SPT*, skin prick test; *HDM*, house dust mite; *na*, not applicable; *SABA*, short acting beta agonists; *ICS*, inhaled corticosteroids. $ Well controlled=ACQ score ≤0.75, partially controlled=ACQ score 0.76-1.49, poorly controlled=ACQ score ≥1.50, *=summation of all positive individual allergen weal sizes

| Cohort SIBRO (In vitro) | | | | |
|---|---|---|---|---|
| **Obesity Status** | **Asthma Status** | **Age (years, age range)** | **Gender** | **BMI ($kg/m^2$)** |
| Non-Obese | Non-asthma | 41-45 | Male | 24.61 |
| Non-Obese | Non-asthma | 26-30 | Male | 21.62 |
| Non-Obese | Non-asthma | 26-30 | Female | 21.01 |
| Non-Obese | Non-asthma | 46-50 | Male | 23.57 |
| Non-Obese | Non-asthma | 41-45 | Male | 22.86 |
| Non-Obese | Non-asthma | 41-45 | Female | 22.21 |
| Non-Obese | Non-asthma | 51-55 | Female | 24.14 |
| Non-Obese | Asthma | 31-35 | Male | 21.74 |
| Non-Obese | Asthma | 26-30 | Female | 24.8 |
| Non-Obese | Asthma | 21-25 | Male | 23.94 |
| Non-Obese | Asthma | 26-30 | Male | 22.69 |
| Non-Obese | Asthma | 51-55 | Female | 21.56 |
| Non-Obese | Asthma | 36-40 | Female | 19.92 |
| Non-Obese | Asthma | 46-50 | Female | 21.1 |
| Non-Obese | Asthma | 46-50 | Female | 23.42 |
| Non-Obese | Asthma | 41-45 | Male | 25.21 |
| Non-Obese | Asthma | 51-55 | Female | 21.56 |
| Non-Obese | Asthma | 51-55 | Female | 21.83 |
| Non-Obese | Asthma | 56-60 | Female | 24.68 |

*BMI*, body max index.

| Cohort A (In vitro) | | |
|---|---|---|
| **Diagnosis** | **Gender** | **Age (years, age range)** |
| Control | Female | 26-30 |
| Control | Male | 26-30 |
| Control | Male | 61-65 |
| Control | Male | 41-45 |
| Control | Female | 31-35 |
| Asthma | Female | 26-30 |
| Asthma | Female | 61-65 |
| Asthma | Female | 26-30 |
| Asthma | Female | 61-65 |
| Asthma | Female | 51-55 |
| Asthma | Female | 41-45 |
| Asthma | Female | 36-40 |
| Asthma | Female | 61-65 |

| | | |
|---|---|---|
| Asthma | Male | 51-55 |
| Asthma | Male | 56-60 |
| Asthma | Male | 31-35 |
| Asthma | Male | 51-55 |

**Supplementary Table S9. Primary human bronchial epithelial cells (HBECs) used in the study. Commercially available and obtained from the study subjects.**

| HBECs source | Condition | Age (years, age range) | Gender | Smoker |
|---|---|---|---|---|
| Epithelix | Healthy | 71-75 | Male | No |
| Epithelix | Healthy | 51-55 | Male | No |
| Epithelix | Healthy | 76-80 | Male | No |
| Epithelix | Healthy | 51-55 | Male | No |
| Epithelix | Healthy | 61-65 | Male | No |
| Epithelix | Healthy | 56-61 | Male | No |
| Lonza | Healthy | 66-70 | Female | No |
| Lonza | Healthy | 21-25 | Female | No |
| Epithelix | Healthy | 51-55 | Female | No |
| Epithelix | Healthy | 16-20 | Male | No |
| Epithelix | Healthy | 61-65 | Female | No |
| Epithelix | Healthy | 16-20 | Male | No |
| Epithelix | Asthma | 36-40 | Male | No |
| Epithelix | Asthma | 51-55 | Male | No |
| Epithelix | Asthma | 41-45 | Male | No |
| Epithelix | Asthma | 56-60 | Male | No |
| Lonza | Asthma | 66-70 | Male | No |
| Lonza | Asthma | 61-65 | Female | No |
| Lonza | Asthma | 11-15 | Female | No |
| Lonza | Asthma | 21-25 | Male | No |
| Epithelix | Asthma | 16-20 | Female | No |
| Epithelix | Asthma | 46-50 | Male | No |
| Epithelix | Asthma | 76-80 | Female | No |
| Epithelix | Asthma | 51-55 | Female | No |
| Epithelix | Asthma | 46-50 | Female | No |

| Cell source | Condition | Age (years, age range) | Gender |
|---|---|---|---|
| Cohort A | Control | 51-55 | Female |
| Cohort A | Control | 71-75 | Male |
| Cohort SIBRO | Non-asthma | 31-35 | Female |
| Cohort A | Asthma | 46-50 | Female |
| Cohort A | Asthma | 36-40 | Female |
| Cohort A | Asthma | 51-55 | Male |
| Cohort A | Asthma | 61-65 | Male |
| Cohort A | Asthma | 46-50 | Female |
| Cohort A | Asthma | 36-40 | Male |
| Cohort A | Asthma | 21-25 | Female |
| Cohort A | Asthma | 61-65 | Female |
| Cohort A | Asthma | 26-30 | Female |
| Cohort A | Asthma | 66-70 | Female |
| Cohort A | Asthma | 61-65 | Female |
| Cohort A | Asthma | 51-55 | Female |
| Cohort SIBRO | Asthma | 41-45 | Male |
| Cohort SIBRO | Asthma | 31-35 | Male |
| Cohort SIBRO | Asthma | 51-55 | Male |

**Supplementary Table S10. Sequences of the primers used in the study.**

| Gene | Forward primer sequence | Reverse primer sequence |
|---|---|---|
| IL1B | CTCTTCGAGGCACAAGGCA | GGCTGCTTCAGACACTTGAG |
| RV-A16 POSITIVE STRAND | CGGGACTGCAAACACTACCT | CACCACGTGTGTCCCTAACA |
| DDX58 | TGATTGCCACCTCAGTTGCT | TCCTCTGCCTCTGGTTTGGA |
| NLRC5 | GCTGGAGGAGGTCAGTTTGC | TGTTTCGGCTCAGGTCAAGT |
| IFIH1 | AGATGCAACCAGAGAAGATCCA | TGGCCCATTGTTCATAGGGT |
| CASP1 | GCCCACCACTGAAAGAGTGA | TTCACTTCCTGCCCACAGAC |
| IFNL2/3 | CTGGGAGACAGCCCAGTTCA | AGAAGCGACTCTTCTAAGGCATCTT |
| IFNB1.1 | CGCCGCATTGACCATCTA | GACATTAGCCAGGAGGTTCTC |
| IFNB1.2 | AGGCCAAGGAGTACAGTCAC | GAGGTAACCTGTAAGTCTGTTAATG |
| EEFA1 | TGAAGTCTGGTGATGCTGCC | CAAAGCGACCCAAAGGTGGA |
| **Gene** | **Primer pair ID** | **Company** |
| IFNL1 | H_IL29_1 | Sigma Aldrich |

**Supplementary Table S11. List of genes included in inflammasome-mediated immune responses and antiviral responses gene sets. Curated from the gene sets available at the GSEA and MSigDB Databases.**

| | |
|---|---|
| Inflammasome-mediated immune responses | TXNIP, NLRP3, NLRP1, PANX1, PYCARD, HSP90AB1, APP, MEFV, P2RX7, NLRC4, BCL2, BCL2L1, TXN, CASP1, PSTPIP1, AIM2, CASP4, CASP5, CASP8, NLRC5, DDX58, IFIH1, IL18, NLRP7, NOD1, TAB1, IRAK3, CHUK, MAP3K8, TAB2, PELI3, TAB3, IKBKB, IL1A, IL1B, IL1R1, IL1RAP, IL1RN, IRAK1, IRAK2, TMEM189-UBE2V1, MAP3K3, MYD88, IRAK4, TOLLIP, MAP2K1, MAP2K6, PELI2, PELI1, MAP2K4, SKP1, MAP3K7, TRAF6, UBE2N, IL1R2, TNIP2, CUL1, IKBKG, RIPK2, SQSTM1, BTRC, RBX1, NAIP, CARD18, CD40LG, CTSB, HSP90AA1, HSP90B1, PYDC1, SUGT1, TNF, TNFSF11, TNFSF14, TNFSF4, IFNG, IL12A, IL12B, IL33, IRF1, TIRAP, CIITA, NLRP12, NLRP4, NLRP5, NLRP6, NLRP9, NLRX1, BIRC2, BIRC3, CARD6, CCL2, CCL5, CCL7, CFLAR, CXCL1, CXCL2, FADD, IFNB1, IL6, IRF2, MAPK1, MAPK11, MAPK12, MAPK13, MAPK3, MAPK8, MAPK9, NFKB1, NFKBIA, NFKBIB, PEA15, RELA, XIAP, NOD2. |
| Antiviral responses | ABCC9, ABCE1, ABCF3, ACTA2, ADAR, ADARB1, AGBL4, AGBL5, AIMP1, AP1S1, APOB, APOBEC1, APOBEC3A, APOBEC3B, APOBEC3C, APOBEC3D, APOBEC3F, APOBEC3G, APOBEC3H, ATG7, AZU1, BAD, BANF1, BATF3, BCL2, BCL2L1, BCL2L11, BCL3, BECN1, BNIP3, BNIP3L, BST2, BTBD17, C17orf85, C19orf2, C19orf66, CARD9, CCDC130, CCL11, CCL19, CCL22, CCL4, CCL5, CCL8, CCT5, CD207, CD40, CD86, CD8A, CDK6, CFL1, CHRM2, CHUK, CLU, CRCP, CREBZF, CXADR, CXCL10, CXCL12, CXCL9, CXCR4, CYP1A1, DCLK1, DDIT4, DDX1, DDX21, DDX3X, DDX41, DDX58, DDX60, DEFA1, DEFA1B, DEFA3, DHX36, DHX58, DMBT1, DNAJC3, DUOX2, EEF1G, EIF2AK2, EIF2AK4, ELMOD2, ENO1, EXOSC4, EXOSC5, F2RL1, FADD, FAM111A, FCN3, FGR, FOSL1, FOXP3, GATA3, GBP1, GBP3, GLI2, GPAM, GTF2F1, HBXIP, HERC5, HMGA1, HMGA2, HNRNPUL1, HSPB1, HYAL1, HYAL2, HYAL3, IFI16, IFI44, IFI44L, IFIH1, IFIT1, IFIT1B, IFIT2, IFIT3, IFIT5, IFITM1, IFITM2, IFITM3, IFNA1, IFNA10, IFNA13, IFNA14, IFNA16, IFNA17, IFNA2, IFNA21, IFNA4, IFNA5, IFNA6, IFNA8, IFNAR1, IFNAR2, IFNB1, IFNE, IFNG, IFNGR1, IFNGR2, IFNK, IFNW1, IKBKB, IKBKE, IKBKG, IL10RB, IL12A, IL12B, IL23A, IL28A, IL28B, IL28RA, IL29, IL33, IL6, ILF3, IRAK3, IRF1, IRF3, IRF5, IRF7, IRF9, ISG15, ISG20, ITCH, IVNS1ABP, KCNJ8, LGALS9, LILRB1, LSM14A, LYST, MAPK11, MAPK14, MAVS, MB21D1, MEF2C, MICA, MST1R, MX1, MX2, NLRC5, NLRP3, NPC2, OAS1, OAS2, OAS3, OASL, ODC1, OPRK1, PCBP2, PENK, PIM2, PLSCR1, PMAIP1, PML, POLR3A, POLR3B, POLR3C, POLR3D, POLR3E, POLR3F, POLR3G, POLR3H, POLR3K, PRF1, PRKRA, PSMA2, PSMB9, PTPRC, PYCARD, RELA, RNASEL, RPS15A, RSAD2, SAMHD1, SERINC3, SERINC5, SLFN11, SPACA3, SPON2, SRC, STAT1, STAT2, STMN1, TBK1, TBX21, TICAM1, TLR3, TLR7, TLR8, TMEM173, TNF, TNFSF4, TPT1, TRIM11, TRIM22, TRIM25, TRIM34, TRIM5, TRIM56, TRIM6, UNC13D, UNC93B1, XCL1, XPR1, ZC3H12A, ZC3HAV1, ZNF175. |

**Supplementary Table S12. All proteins available for PEA measurements used as statistical background for STRING analyses of targeted proteomics data.**

| UniprotID |
|---|
| P03950, Q9Y5C1, O95445, Q96KN2, P15907, P12830, P00915, P07451, P22748, Q9NQ79, P49747, Q16627, P55774, P13501, P13987, P08709, P03951, P39060, Q9BXJ1, P06681, Q9BXR6, P20023, P01034, P27487, Q12805, P17813, Q9UGM5, Q15485, Q16769, P22749, Q14393, P08581, P0DOY2, P17936, P24592, P11215, P05362, P32942, P16871, Q14767, Q8NHL6, Q8N423, O75023, P05451, P23141, P12318, O75015, P14151, Q9Y5Y7, P42785, P11226, P10721, P15529, Q16853, P01033, Q13361, Q9H1U4, P13591, O00533, P46531, O14786, P59665, P80188, P14543, Q99650, P19021, P55058, P05154, P07359, Q13093, O15031, Q15113, Q12884, Q13332, Q06141, P35542, Q14515, P00441, Q96H15, P24821, P22105, P35443, P05543, P20062, Q03167, Q15582, P07478, P35590, P07911, P19320, Q6EMK4, P04070, Q86SJ2, Q7Z5R6, Q8TD06, P15848, P08237, O43521, Q02742, P21810, Q06520, P11274, Q8N5S9, Q9BQT9, Q9NX58, P04637, Q49AH0, P02462, O43186, P0CG37, Q9UBG0, Q8WYN0, P16562, Q9UK85, P25685, P52564, Q13561, P22681, Q01543, P47929, A4D1B5, Q9NS71, O00451, P01275, P01242, Q9Y662, O60243, O75054, Q9NRM6, Q9UKR0, Q96I82, Q6UXK5, Q8N2G4, Q12912, Q6UB28, Q16653, O94760, O00221, Q969V3, Q92982, O95644, P23515, Q9UBM4, P30041, Q9NZ53, Q10471, Q96SM3, P58294, Q9HCU5, Q7Z5A7, O14904, O14917, Q13576, P57771, Q07960, Q9NZN5, Q6UXD5, Q9C0C4, O96013, Q96LC7, O43699, Q9P0V8, Q9H156, Q9H5Y7, Q86WV1, O14662, O43752, O95988, Q9Y6A5, Q9Y2W6, P09758, O95183, Q92558, Q9UPY6, Q96PQ0, Q13105, Q16698, Q76LX8, P35318, O00253, P35475, Q9BYF1, P22004, P35218, P31997, P07711, Q99895, O94907, P12104, P19883, P27352, P51161, P01241, Q9UK05, P04792, Q9UJM8, P18510, Q9HB29, Q8TAD2, Q8NEV9, Q14213, P24394, Q04760, P78380, P41159, P06858, P31994, P47992, Q9UEW3, Q9UKP3, P16860, Q9Y6K9, Q8IYS5, Q13219, P26022, P09874, P01833, Q99075, Q14005, P51888, Q16651, P02760, P25116, P21980, P12931, Q14242, Q15109, P00797, Q9BQR3, Q13043, Q8IW75, Q9UIB8, Q9NQ25, Q99523, Q9BUD6, P04179, P07204, P40225, P40225, P13726, O14763, Q9Y6Q6, O14836, Q12866, Q96IQ7, Q9BWV1, O00220, O43915, Q15389, Q02763, Q92583, P29965, P07585, O00182, P09601, P39900, P09237, P01127, Q9BQ51, P01730, P49763, Q96D42, P15144, P20160, Q13867, P33151, P15085, P15086, P42574, P07339, Q9UBR2, Q16663, O15467, O00175, Q13740, P36222, Q13231, P02452, Q9NPY3, Q12860, Q9H2A7, P04080, P19957, P54760, P00533, P16422, P16581, P15090, P17931, P56470, P28799, Q99988, Q9HCN6, P08833, P18065, Q16270, P05107, P13598, P14778, P27930, Q96F46, O95998, P01589, P08887, Q9Y624, Q92876, P01130, P36941, Q9NQ76, P08253, P08254, P14780, Q99727, P24158, P05164, P02144, Q9UM47, P10451, Q15166, O75594, P98160, P05121, P16284, P04085, Q8NBP7, P80370, P16109, P35247, Q9HD89, Q99969, Q86VB7, Q96PL1, Q9HCB6, Q01638, P13686, P10646, P00750, P02786, Q07654, Q5T2D2, Q9Y275, P19438, P20333, O14798, Q92956, P25445, P30530, P78324, Q03405, P04275, P16860, Q10588, Q9UKK9, P16112, P05067, Q9BY76, P15289, P50895, P15291, O43505, P08236, P08118, P00918, P23280, Q9UBX1, P11717, Q6YHK3, Q8N6Q3, Q9NNX6, P48960, Q8TCZ2, Q4KMG0, Q6UXG3, O43405, Q5KU26, Q8IWV2, P24387, Q9Y240, Q86T13, Q15828, Q9NZV1, Q6UXH1, P47712, Q02487, Q9UBP4, Q14118, Q07108, Q13822, Q96AP7, Q96RD9, P30043, O95633, P21217, Q11128, P01215, P55808, Q8TDQ0, P04233, Q6UXH9, P55103, P08648, P05556, P78552, O43278, O43291, Q08431, Q16363, Q14696, Q99538, Q6GTX8, P06734, P19256, P14174, O00339, Q99972, Q14112, Q99983, Q9UKJ1, P23284, P30086, Q5VY43, P09619, Q99497, P07237, P48745, P10586, Q9Y6N7, Q14162, Q96QR1, O75326, O43464, P00995, Q9NQ38, O00241, P31948, O95721, P63313, P04066, O14773, Q969Z4, P29350, Q8TEU8, Q9Y279, Q01151, P09038, P42701, P18627, Q9UQV4, O76036, Q13241, P48061, P10747, O15123, P05089, P40933, Q9Y653, P43629, P29474, P21246, Q15116, Q8WXI7, Q5T4W7, O15169, O15444, Q9NRJ3, Q9H5V8, P80162, P28325, Q8NFT8, Q13541, O95750, P12034, P49771, Q13651, Q08334, P29460, Q13261, Q16552, Q9P0M4, Q13478, P14784, Q9NYY1, Q9UHF4, Q8N6P7, Q13007, P15018, P42702, P03956, P09238, P20783, Q99748, P13725, P80511, Q13291, Q8IXJ6, O95630, P50225, P30203, Q969D9, P01374, O14788, Q9GZV9, Q9NSA1, P10147, Q14116, P21583, O00300, P00749, P13500, P00813, Q14790, Q99731, P78556, P55773, P13236, P25942, P02778, O14625, P42830, Q07325, P78423, P01579, P01583, P35225, P60568, O95760, P05112, P13232, P10145, P01137, P09603, P80075, P80098, Q99616, Q9BZW8, Q9NZQ7, P06127, P01732, O43508, P01375, O43557, Q07011, P22301, P05113, P51671, P01138, P39905, P01135, P50591, P05231, P14210, P15692, P09341, Q7Z6M3, P27540, Q13490, P16278, P78410, Q9UHC6, Q15517, P28845, P78310, Q9UMR7, Q8WTT0, Q8WXI8, Q6UXB4, Q6EIG7, Q9BXN2, Q07065, Q13574, O00273, Q9UN19, Q14203, P19474, Q9GZT9, Q04637, P63241, Q96P31, Q6DN72, Q96DB9, P14317, P50135, P52294, Q8N608, O43736, Q9UKX5, P23229, P18564, Q8IU57, Q00978, P51617, Q9NWZ3, Q05084, P08727, Q8NHJ6, O60449, P48740, P35240, P16455, Q96SB3, P34130, Q12968, Q03431, O75475, Q06830, P30044, Q6ZUJ8, Q9HCM2, Q14435, O95786, P58499, O94992, Q04759, O43597, Q9Y2J8, O60880, Q9UQQ2, Q9Y3P8, P78362, P52823, P30048, Q12933, Q92844, O14867, Q9NP99, Q9C035, Q15661, Q9UNE0, Q96PD2, P05412, Q05516, P23526, Q9UHX3, Q8IZP9, P51693, O95841, O43827, P50995, P09525, Q9UBU3, P20711, P40259, P19022, Q9HBB8, Q9H4D0, Q8N1Q1, P21964, P43234, Q9Y5K6, Q6WN34, Q15846, Q76M96, P46109, Q9NY25, Q9H6B4, Q9NR28, P09417, Q9UHL4, P98082, P27695, O75356, Q6UWV6, P12724, Q96LA6, P09467, P22466, P09104, P35754, O75791, P51858, Q01973, Q8WX77, P26010, O43240, Q16773, P46379, Q96JA1, A6NI73, Q86VZ4, Q9NPH0, Q16820, Q641Q3, Q8NI22, O95544, Q92692, Q9NQX5, O95502, Q15155, Q9UKJ0, Q02790, Q9NWQ8, P09668, Q92520, P41236, P25815, Q9BYZ8, Q9BZR6, P00352, P16083, Q8WTU2, Q9BQB4, Q13275, P35237, P50452, Q9Y286, Q04900, Q8WVQ1, Q8NBJ7, O00161, P31431, P29017, P52888, Q8NBS9, P19971, P01222, Q03403, Q9GZM7, Q06418, P40818, P13611, P28907, P30533, Q16620, P12644, Q96GW7, Q2TAL6, P22223, P55285, P48052, P14384, P25774, Q8N126, Q8TD46, Q8TDQ1, Q08708, O94779, Q9P126, Q8IUN9, Q9UBT3, P53634, Q9P0K1, O75077, Q8NBI3, O15197, P52798, Q08345, P15311, Q96LA5, O00214, P56159, O60609, P78333, P09919, P15509, O14793, Q16775, Q01344, P57087, Q16719, Q9BS40, Q6UX15, O43155, Q6ISS4, O43561, Q14108, P21757, Q8NFP4, O15232, P55145, P10636, Q02083, P08473, O95185, P41271, O14594, Q92823, O60462, Q9NR71, Q9HAN9, Q9BZM5, Q16288, P41217, P16234, O43157, Q9ULL4, P15151, Q2VWP7, Q96B86, Q6NW40, Q9HCK4, Q2MKA7, Q6ZMJ2, Q96GP6, Q92765, P37023, Q9Y336, Q9BZZ2, Q9H3U7, P17405, Q92752, Q08629, P04216, Q9H3S3, Q9HAV5, Q96NZ8, P29460, P29459, O95727, P12544, Q9NP84, O75509, Q03393, O60242, Q9BTE6, Q9BYC5, P19801, Q9UJ72, P07306, O15263, Q86Z14, P49789, Q10589, P55291, Q12864, P63098, Q9Y2V2, O94985, P40198, Q16619, Q9Y4X3, Q8IX05, P08962, P41208, Q9Y5P4, P52943, P78560, P32926, P16444, Q9H4A9, Q13444, P51452, Q96EP0, P14625, P42892, Q5JZY3, P23588, P21802, P13284, O76070, P36269, P09211, P09466, O15496, P32456, P30519, P24071, Q6UXK2, Q9UK53, P29218, Q9H0C8, Q8IU54, P26951, P24001, Q96EK5, P43628, Q6UWL6, Q9NS15, P48357, Q9Y6D9, P20138, P41227, O00308, Q969M7, P58417, P07196, P06748, Q13451, Q9Y680, Q8TCT1, Q15126, Q9NRG1, Q9UHV9, P11464, O14944, Q96B36, Q06323, Q96IU4, Q92597, Q9H477, P23443, Q8N474, P37108, Q9H4F8, P13385, Q9BW30, Q99426, Q96RJ3, P18031, Q6UX27, Q16864, Q9UKS7, Q9P0J1, Q9Y478, P30838, O95994, O95831, Q8NDB2, P55957, Q13145, P01258, P27797, O43570, Q9ULX7, P48730, Q9HAW4, Q11201, O00748, Q02246, P06850, Q8NC01, P23582, Q86SJ6, P42658, Q02880, Q9Y5L3, O43854, P98073, Q9UHF1, Q96RT1, P01588, Q0Z7S8, Q12778, Q9NQ88, O60760, Q13308, O14713, O75569, P09960, Q9GZY6, P01229, Q7L5Y9, P40121, Q9Y5V3, P53582, Q03426, Q9Y4K4, Q15797, Q9NXA8, Q9Y5A7, P19878, O60934, P80303, P20472, P49023, P68106, O60240, O15357, Q9NRA1, Q8IUK5, Q86SR1, P35070, Q07954, P25786, Q9BXJ7, Q8N8S7, P53539, P61244, O75688, P50749, P20936, Q12913, O75787, Q9UKL0, P49788, Q7LG56, Q86WD7, Q15165, Q9UNK0, O00186, P19429, P07332, P09769, P07947, Q7L8A9, P42768, O75354, P21589, Q8TE58, P40222, P04083, P16870, P06731, P13688, O60911, O95971, P09326, Q9UBG3, Q9UJ71, P38936, O00548, P78325, Q9NQ30, P29317, Q13158, 6BAA4, Q14512, P15328, P41439, P09958, P35052, P08069, P06756, P18084, P15260, Q9UBX7, Q9UKR3, Q9P0G3, O60259, O95274, Q16674, Q13421, P50579, P21741, Q99717, Q96NY8, P01298, O00592, O00622, P31949, P26447, P07949, P04626, P21860, Q15303, Q9BXY4, O14828, Q9BYH1, P09486, P18827, P56279, P37173, P48307, Q9HBG7, O15455, O75888, Q9NS68, O95407, P00519, P07948, Q14508, P35916, Q6UXB2, P08670, Q9Y5W5, O95388, O43895, P15514, Q14956, Q16790, P26842, P32970, O43927, P48023, P09382, P10144, P20718, O75144, Q29983, Q29980, P01133, P43489, P35968, O00233, Q9UJY5, P15121, P05187, P55008, O43707, P02771, Q12904, P20273, Q9Y644, P06865, Q9BYE9, Q9P1Z2, Q9UDT6, P42575, P30260, Q99795, Q9NTU7, P0DN86, Q496F6, P20849, P02745, O95715, P28838, Q9H0P0, Q9H773, P55039, Q14241, Q9BS26, Q53H82, P55789, O60907, O43524, O60763, Q9HD26, Q14353, O14558, P09105, P13747, P01591, P49441, Q14773, Q9UMF0, P38484, P01584, Q96PD4, Q9Y5K2, P33241, Q9H8J5, Q86SF2, P32004, P62166, P43490, P09110, O60542, P08397, Q9H3G5, P12872, P01303, Q8WUW1, Q9Y2B0, Q92832, Q96FQ6, Q93096, O75695, Q08174, Q8IWL2, P20340, P36888, P28827, P42331, P35637, O76038, P13521, Q96I15, Q9UHD8, O60575, Q99536, P48643, P51580, Q96J42, P13693, O95379, Q9BSL1, Q13459, Q8NEZ2, Q5VIR6, P17948, Q7Z5L0, Q6PCB0, Q9Y5K8, Q7Z739. |
