## Supplementary Figures for "Rhinovirus-induced epithelial RIG-I inflammasome activation suppresses antiviral immunity and promotes inflammatory responses in virus-induced asthma exacerbations and COVID-19"

### Supplementary Figure S1

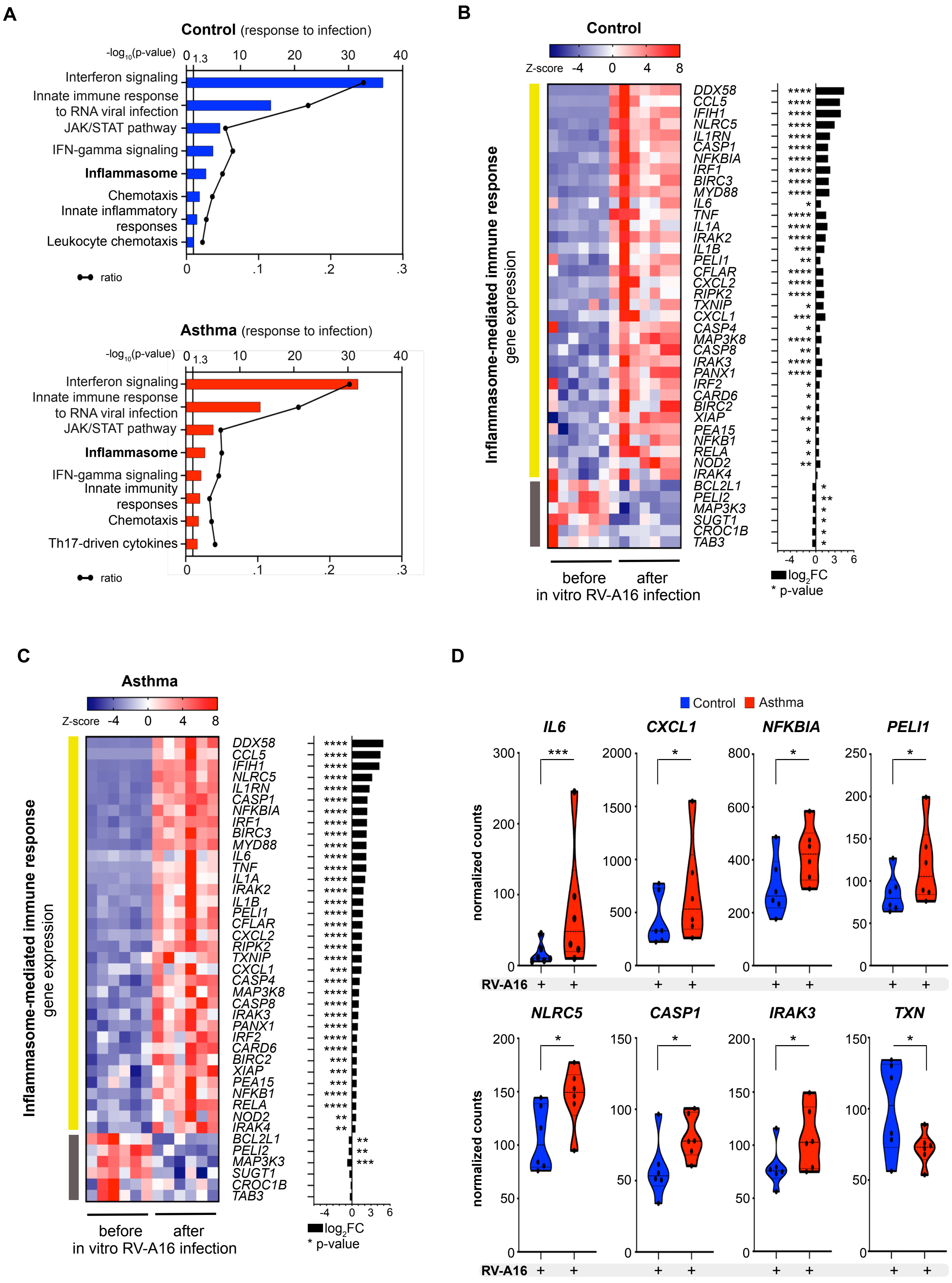

**Supplementary Figure S1**

**Inflammasome- and IL-1β-mediated immune responses are augmented in patients with asthma after infection with rhinovirus**

**A)** Top significantly enriched pathways within 100 most significantly upregulated genes in HBECs from controls (upper panel) and patients with asthma (lower panel) after RV-A16 infection in vitro (control n=6, asthma n=6; from GSE61141). Black line represents a ratio of genes in the experiment over the whole pathway set. **B-C)** Heatmap of genes encoding inflammasome-mediated immune responses in HBECS from **B)** control individuals and **C)** patients with asthma after in vitro RV-A16 infection presented together with the corresponding $log_2$ fold change (FC) (control n=6, asthma n=6; GSE61141). Yellow and grey left side color bars represent genes upregulated or downregulated, respectively. **D)** Genes encoding inflammasome-mediated immune responses significantly different between control individuals and patients with asthma after in vitro RV-A16 infection (control n=6, asthma n=6; from GSE61141). (*) represents a significant difference as indicated. p-value: *<0.05; **<0.005; ***<0.0005, ****<0.00005. Heatmap displays normalized gene expression across the groups (row normalization). Inflammasome-mediated immune responses gene set was curated based on databases and ontologies listed in the Molecular Signatures Database (MSigDB, Broad Institute, Cambridge). *HBECs*, differentiated human bronchial epithelial cells; *RV-A16*, rhinovirus A16; *UV-RV-A16*, UV-treated rhinovirus A16.

### Supplementary Figure S2

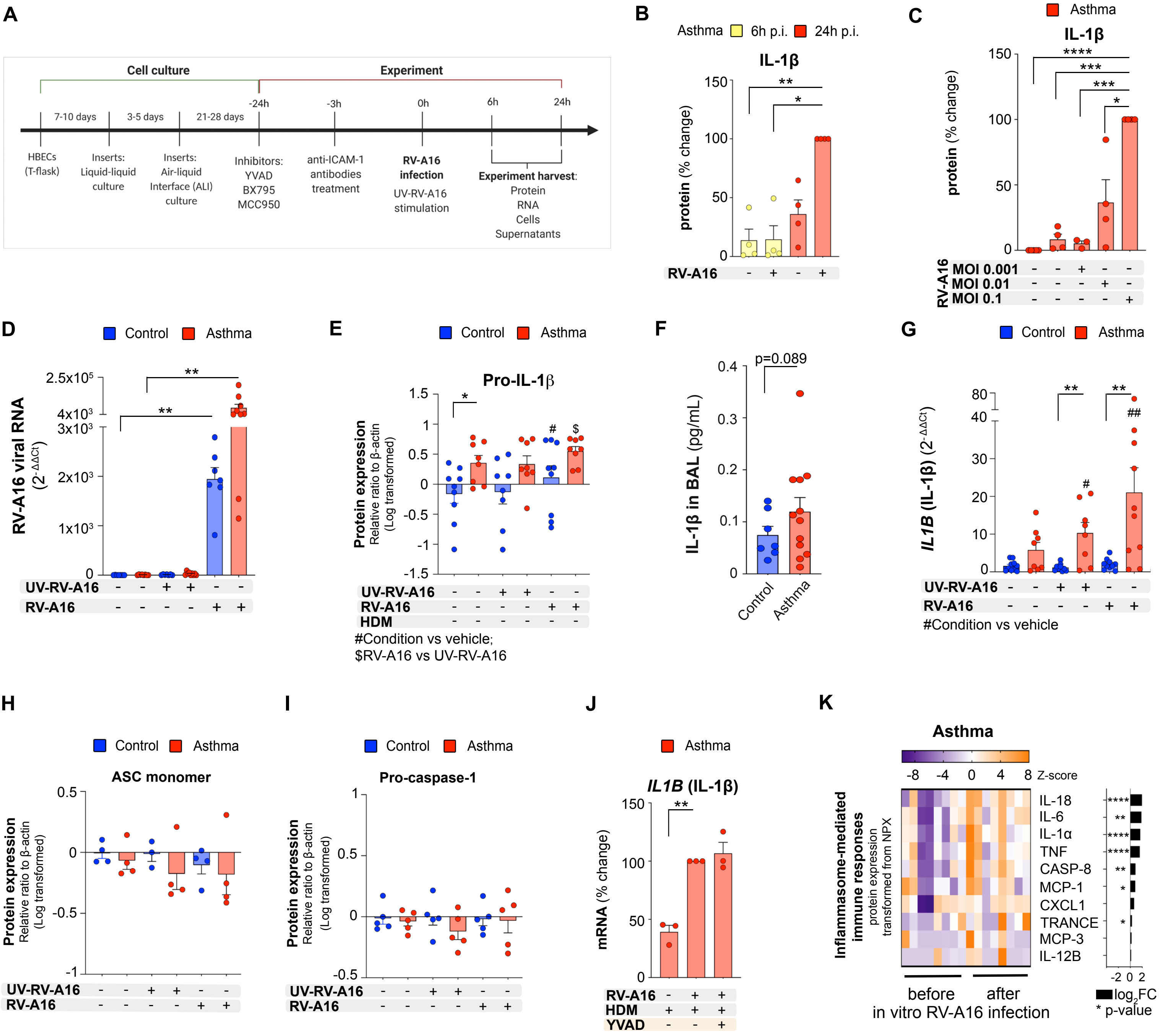

**Supplementary Figure S2**

**Augmented rhinovirus-induced inflammasome activation in airway epithelium from patients with asthma**

**A)** Detailed experimental in vitro model overview. Primary human bronchial epithelial cells (HBECs) from control individuals (blue) or patients with asthma (red) were differentiated for 21-28 days in the Air-Liquid Interface (ALI) culture. 24h prior rhinovirus A16 (RV-A16) infection, cells were stimulated with caspase-1 inhibitor (YVAD), TBK1/IKKε inhibitor (BX795) and NLRP3 inflammasome inhibitor (MCC950) or vehicle. To block cell entry of RV-A16, HBECs were incubated with anti-ICAM-1 antibodies 3h before the infection. Cells were infected with RV-A16 at the multiplicity of infection (MOI) 0.1, 0.01 and 0.001 or the respective UV-RV-A16 controls. Protein, RNA, cells, and supernatants were harvested at 6h and 24h after infection. **B)** IL-1β release to the apical compartment was assessed by ELISA for 6h (yellow) and 24h p.i. (red) (n=4). Data are presented as the percentage of the response after 24h p.i. **C)** IL-1β release to the apical compartment assessed by ELISA 24h after RV-A16 infection in the MOI 0.1, 0.01, and 0.001, combined with HDM pre-stimulation (n=4). Data are presented as the percentage of the response after RV-A16 infection in the MOI 0.1. **D)** expression of *RV-A16 positive strand* (RV-A16 viral RNA) was assessed using RT-PCR, and presented as the relative quantification ($RQ=2^{-\Delta\Delta Ct}$) as compared to the vehicle condition in the HBECs from control individuals (control n=7, asthma n=7-9). **E)** Quantification of densitometry results of pro-IL-1β protein assessed by Western Blot, presented as a log-transformed ratio relative to β-actin and normalized to the vehicle condition in control individuals (control n=9, asthma n=8). **F)** IL-1β protein in the bronchoalveolar lavage (BAL) fluid of control subjects and patients with asthma (cohort SIBRO), assessed using the mesoscale platform and Welsh`s t-test. (control n=8, asthma n=12). **G)** mRNA expression of *IL1B* (IL1β) was assessed using RT-PCR, and presented as a relative quantification ($RQ=2^{-\Delta\Delta Ct}$) compared to the vehicle from the controls (control n=9-10, asthma n=8-10). **H-I)** Quantification of densitometry results from **H)** ASC and **I)** pro-caspase-1 protein expression assessed by Western Blot, presented as a log-transformed ratio relative to β-actin expression and normalized to the vehicle condition in the HBECs from the control individuals (ASC: control n=4, asthma n=4; caspase-1: control n=5, asthma n=5). **J)** mRNA expression of *IL1B* (IL1β) (n=3), in HBECs from patients with asthma in the presence or absence of caspase-1 inhibitor (YVAD). Data are presented as the percentage of the response after HDM+RV-A16 treatment. **K)** Heatmap of proteins associated with the inflammasome-mediated immune responses after in vitro RV-A16 infection in HBECs from patients with asthma analyzed with the Proximity Extension Assay (PEA) proteomics, transformed from the normalized protein expression (NPX), and presented together with the $log_2$ fold change (FC) (black bars) (control n=5, asthma n=8). Proteins were measured in the apical compartment of the ALI cultures. HBECs from patients with asthma are presented in red, HBECs from control individuals are presented in blue. (*) represents a significant difference as indicated. (#) represents a significant difference of indicated condition as compared to the vehicle from the same group. ($) represents a significant difference between RV-A16 and UV-RV-A16 condition. Bar graph data show mean ± SEM analysed with one-way ANOVA (Kruskal-Wallis test), RM one-way ANOVA (Friedman test) or mixed-effects model, as appropriate, depending on the data relation (paired or unpaired) and distribution (if not mentioned differently), *p-value≤0.05, **p-value≤0.01, ***p-value≤0.001, ****p-value≤0.0001. Figure S2A was prepared with Biorender.com. *ALI*; Air-liquid interface cultures*; BAL*; Bronchoalveolar lavage, *HBECs*, differentiated human bronchial epithelial cells; *HDM*, house dust mite; *RV-A16*, rhinovirus A16; *UV-RV-A16*, UV-treated rhinovirus A16; *YVAD*, ac-YVAD-cmk (caspase-1 inhibitor); *anti-ICAM-1*, anti-ICAM-1 antibody; *BX795*, TBK1/IKKε inhibitor; *MCC950*, NLRP3 inflammasome inhibitor; *MOI*, multiplicity of infection; *p.i.*, post-infection; *NPX*, normalized protein expression.

### Supplementary Figure S3

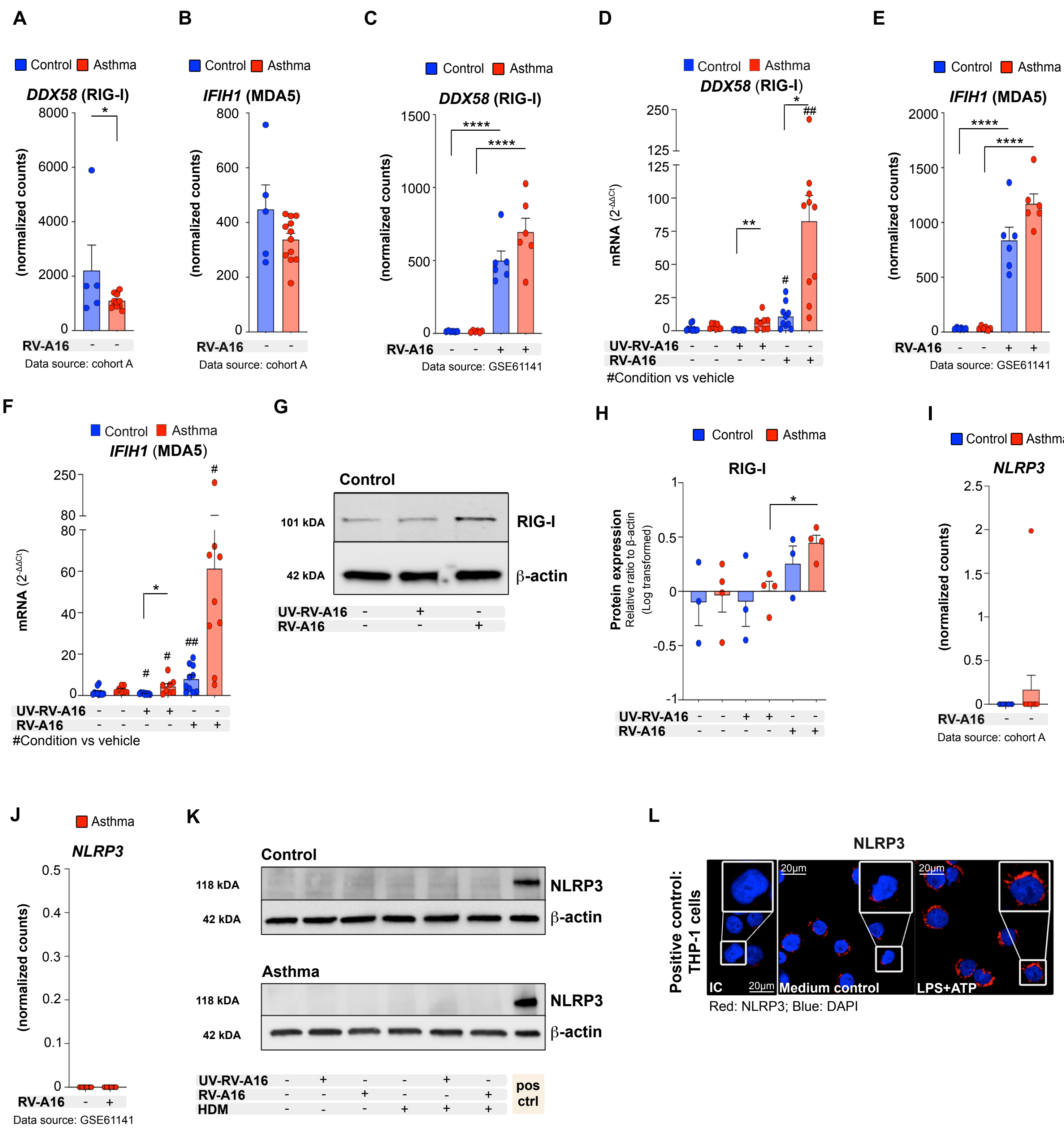

**Supplementary Figure S3**

**Rhinovirus-induced RIG-I, but not NLRP3 or MDA5 inflammasome activation in bronchial epithelium in asthma**

**A-B)** Expression of **A)** *DDX58* (RIG-I) and **B)** *IFIH1* (MDA5) in HBECs at baseline (control n=5, asthma n=12). **C)** Expression of *DDX58* (RIG-I) after RV-A16 infection (control n=6; asthma n=6). **D)** mRNA expression of *DDX58* (RIG-I) was assessed using RT-PCR and presented as a relative quantification ($RQ=2^{-\Delta\Delta Ct}$) as compared to the vehicle condition in HBECs from control subjects (control n=9-10, asthma n=9-10). **E)** Expression of *IFIH1* (MDA5) after RV-A16 infection (control n=6; asthma n=6). **F)** mRNA expression of *IFIH1* (MDA5) was assessed using RT-PCR and presented as a relative quantification ($RQ=2^{-\Delta\Delta Ct}$) as compared to the vehicle condition in HBECs from control subjects (control n=9-10, asthma n=9-10). **G)** Representative Western Blot images of RIG-I protein expression in HBECs from control individuals (n=4). **H)** Quantification of densitometry of RIG-I protein expression, presented as a log-transformed ratio relative to β-actin and normalized to the vehicle condition in HBECs from control subjects (control n=3, asthma n=4). **I-J)** Expression of *NLRP3* in **I)** in HBECs at baseline (control n=5, asthma n=12), and **J)** HBECs from patients with asthma after RV-A16 infection (n=6). **K)** Representative Western Blot images of NLRP3 protein in the HBECs from control subjects (upper panel) and patients with asthma (lower panel) in all analyzed conditions, showed next to the positive control of lipopolysaccharide (LPS)-stimulated monocytes. Control n=3, asthma n=4, LPS-stimulated primary human monocytes n=7. **L)** Representative confocal images of THP-1 cells used as positive controls: (n=2); scale bars: 20μm. HBECs from patients with asthma are presented in red, HBECs from control individuals are presented in blue. Transcriptome data are presented as normalized counts. (*) represents a significant difference between indicated conditions. (#) represents a significant difference of indicated condition as compared to the vehicle from the same group. Bar graph data present mean ± SEM, p-value: *<0.05; **<0.005; ***<0.0005, ****<0.00005. Data presented on the figure S3A, B, I: cohort A, S3C, E, J: GSE61141. *IC*; Isotype control; *HBECs*, differentiated human bronchial epithelial cells; *RV-A16*, rhinovirus A16; *UV-RV-A16*, UV-treated rhinovirus A16; *pos ctrl*, positive control; *LPS*, lipopolysaccharide; *ATP*, adenosine triphosphate.

### Supplementary Figure S4

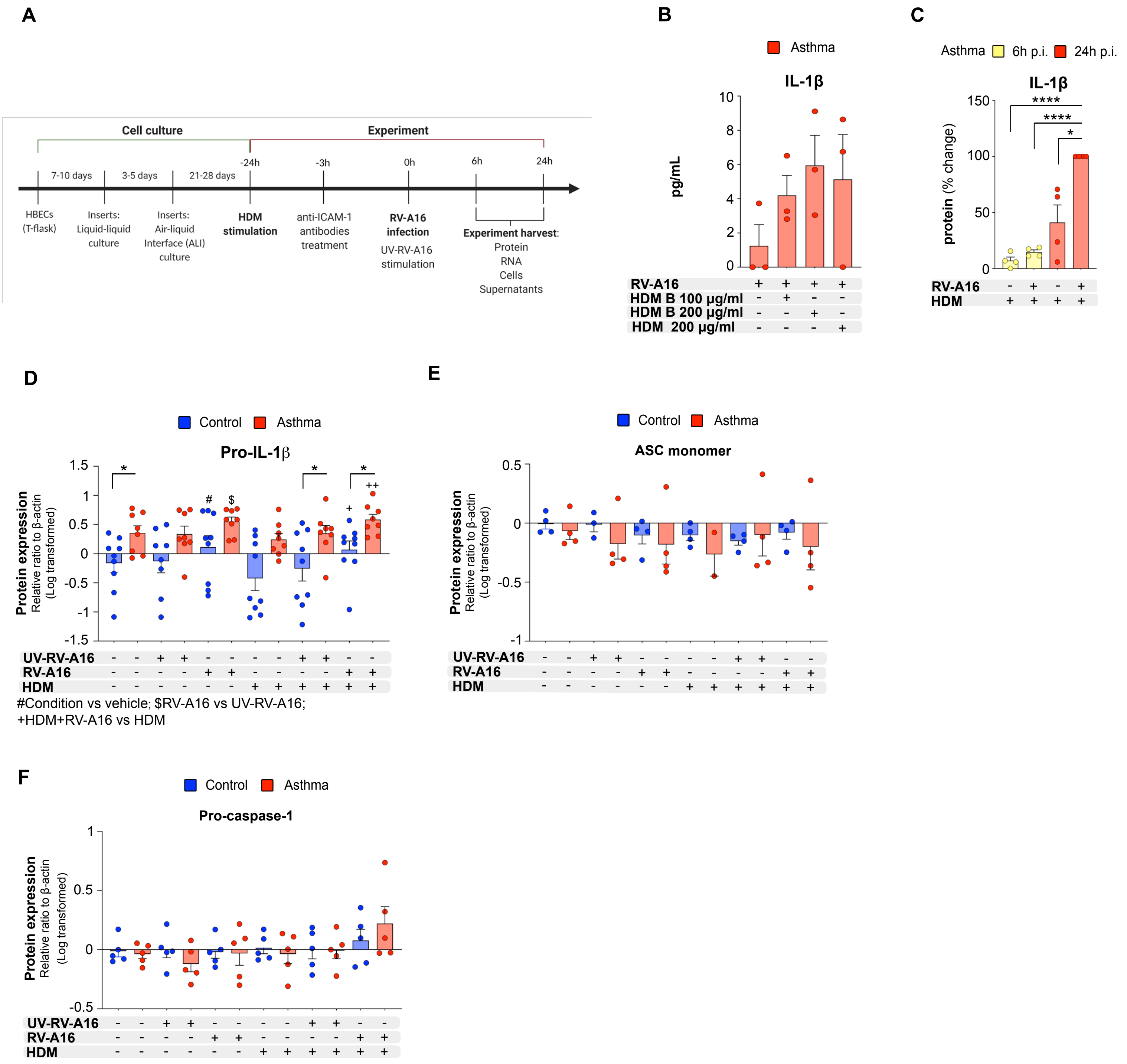

**Supplementary Figure S4**

**House dust mite enhanced rhinovirus-induced inflammasome activation in bronchial epithelium in asthma**
**A)** Detailed experimental in vitro model overview. Primary human bronchial epithelial cells (HBECs) from control individuals (blue) or patients with asthma (red) were differentiated for 21-28 days in the Air-Liquid Interface (ALI) culture. 24h prior rhinovirus A16 (RV-A16) infection, cells were stimulated with house dust mite (HDM) (200μg/mL) or vehicle. Cells were infected with RV-A16 at the multiplicity of infection (MOI) 0.1 or the respective UV-RV-A16 controls. Protein, RNA, cells, and supernatants were harvested at 6h and 24h after infection. **B-C)** IL-1β release to the apical compartment was assessed by ELISA **B)** two independent HDM extracts, HDM (the main one used in the manuscript) and HDM B (similar extract from a different company), were tested 24h after RV-A16 infection (n=3); and **C)** for 6h (yellow) and 24h p.i. (red) (n=4). Data are presented as the percentage of the response after 24h p.i. **D)** Quantification of densitometry results of pro-IL-1β protein assessed by Western Blot, presented as a log-transformed ratio relative to β-actin and normalized to the vehicle condition in control individuals (control n=9, asthma n=8). **E-F)** Quantification of densitometry results from **E)** ASC and **F)** pro-caspase-1 protein expression assessed by Western Blot, presented as a log-transformed ratio relative to β-actin expression and normalized to the vehicle condition in the HBECs from the control individuals (ASC: control n=4, asthma n=4; caspase-1: control n=5, asthma n=5. HBECs from patients with asthma are presented in red, HBECs from control individuals are presented in blue. (*) represents a significant difference as indicated. (#) represents a significant difference of indicated condition as compared to the vehicle from the same group. (&) represents a significant difference upon HDM treatment when compared to the respective condition without HDM. ($) represents a significant difference between RV-A16 and UV-RV-A16 condition. (+) represents a significant difference between HDM+RV-A16 and HDM condition. Bar graph data show mean ± SEM analysed with one-way ANOVA (Kruskal-Wallis test), RM one-way ANOVA (Friedman test) or mixed-effects model, as appropriate, depending on the data relation (paired or unpaired) and distribution (if not mentioned differently), *p-value≤0.05, **p-value≤0.01, ***p-value≤0.001, ****p-value≤0.0001. Figure S4A was prepared with Biorender.com. *ALI*; Air-liquid interface cultures*; HBECs*, differentiated human bronchial epithelial cells; *HDM*, house dust mite; *RV-A16*, rhinovirus A16; *UV-RV-A16*, UV-treated rhinovirus A16; *MOI*, multiplicity of infection; *p.i.*, post-infection.

### Supplementary Figure S5

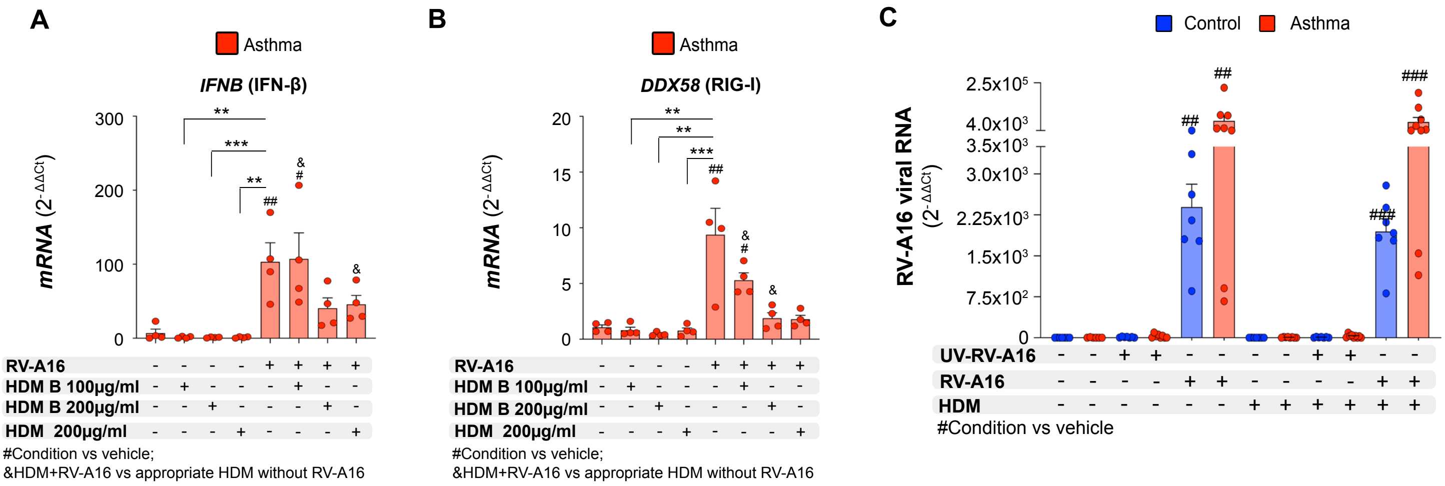

**Supplementary Figure S5**

**House dust mite impaired interferon responses in rhinovirus-infected bronchial epithelium of patients with asthma**

mRNA expression of **A)** *IFNB* (IFN-$\beta$) and **B)** *DDX58* (RIG-I) in HBECs from patients with asthma pre-treated with two HDM extracts from different manufacturers or vehicle for 24h followed by an infection with rhinovirus A16 (RV-A16) in the multiplicity of infection (MOI) 0.1, (n=4), assessed using RT-PCR. **C)** expression of *RV-A16 positive strand* (RV-A16 viral RNA) was assessed using RT-PCR, and presented as the relative quantification ($RQ=2^{-\Delta\Delta Ct}$) as compared to the vehicle condition in the HBECs from control individuals (control n=7, asthma n=7-9). (*) represents a significant difference between indicated conditions. (#) represents a significant difference of indicated condition as compared to the vehicle. (&) represents a significant difference between HDM+RV-A16 treatment and HDM only. Graph data present mean $\pm$ SEM analysed with one-way ANOVA (Kruskal-Wallis test), RM one-way ANOVA (Friedman test) or mixed-effects model, as appropriate, depending on the data relation and distribution, *p-value≤0.05, **p-value≤0.01, ***p-value≤0.001. *HBECs*, differentiated human bronchial epithelial cells; *RV-A16*, rhinovirus A16; *HDM*, house dust mite.

### Supplementary Figure S6

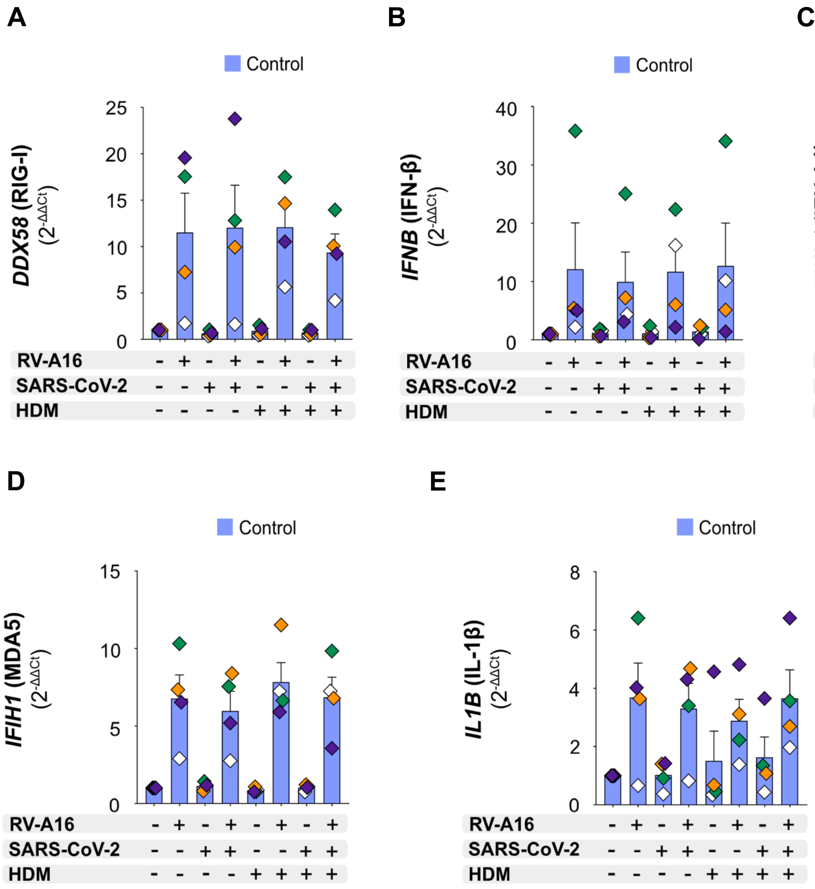

**Supplementary Figure S6**

**Pre-existing rhinovirus infection attenuated SARS-CoV-2 infection, but augmented epithelial inflammation in asthma**

mRNA expression of **A)** *DDX58* (RIG-I), **B)** *IFNB* (IFN-$\beta$), **C)** *IFNL1* (IFN-$\lambda$1), **D)** *IFIH1* (MDA5), and **E)** *IL1B* (IL-1$\beta$) was assessed using RT-PCR and presented as relative quantification (RQ=$2^{-\Delta\Delta Ct}$) compared to vehicle condition (control: n=4). Data were analysed with Friedman's test. Bars depict the mean $\pm$ SEM, whereas color-coded diamonds (control subjects) show individual data from the same donor. *RV-A16*, rhinovirus A16; *HDM*, House Dust Mite; *HBECs*, Human Bronchial Epithelial Cells; *SARS-CoV-2*, Severe Acute Respiratory Syndrome Coronavirus

**Supplementary Figure S7**

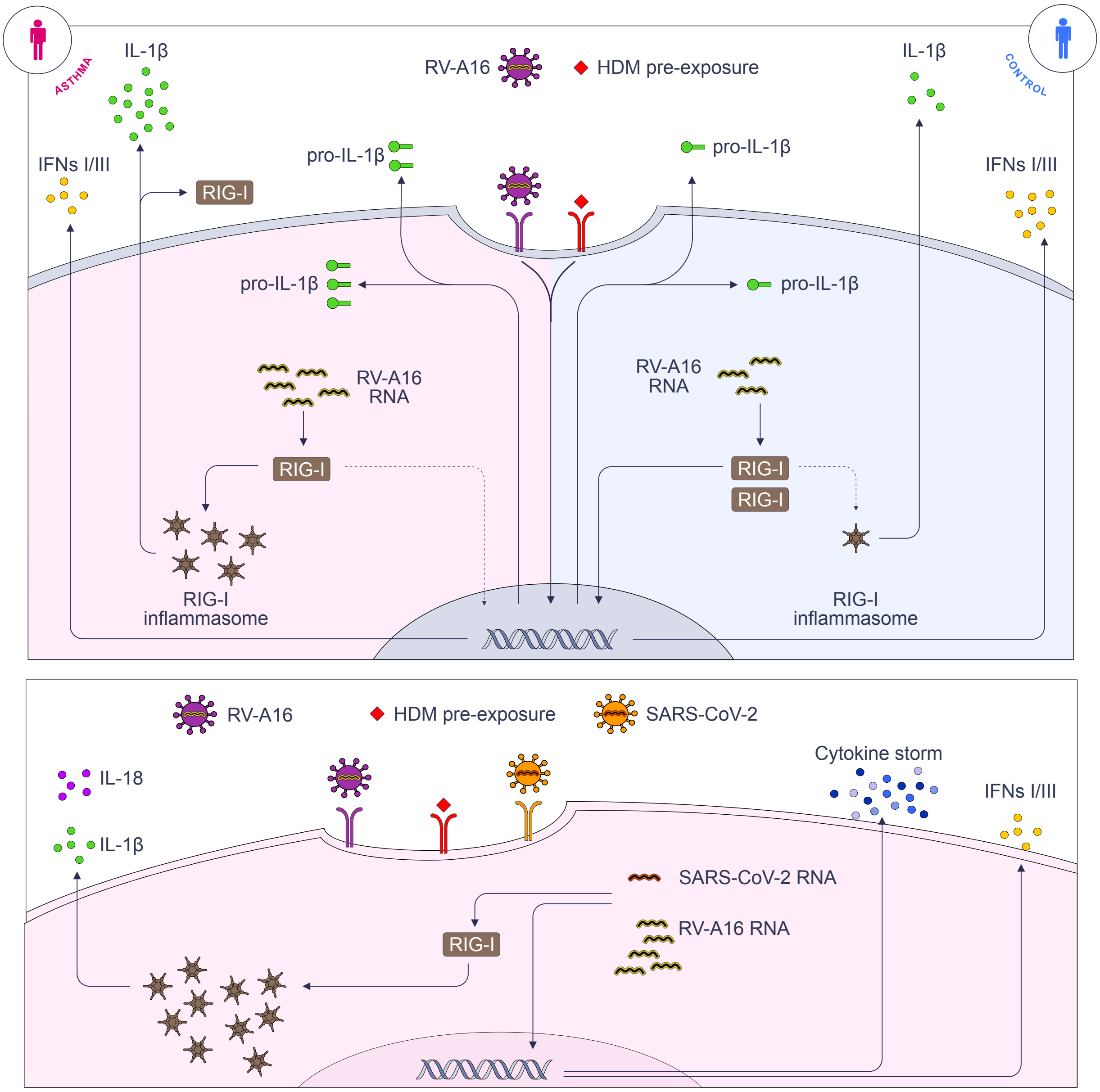

**Supplementary Figure S7**

**The role of epithelial RIG-I signaling in asthma**

**(Upper panel)** Rhinovirus infection in humans is sensed in bronchial airway epithelium via retinoic acid-inducible gene I (RIG-I) helicase. This leads to the recruitment of apoptosis-associated speck like protein containing a caspase recruitment domain (ASC), oligomerization and RIG-I inflammasome activation. Virus-induced RIG-I inflammasome activation- and IL-1β-mediated immune responses are highly augmented in patients with asthma, which is responsible for the functional impairment of the RIG-I-dependent antiviral response, prolonged viral clearance, and unresolved inflammation in asthma. Pre-exposure to house dust mite (HDM) amplifies rhinovirus-induced epithelial injury in patients with asthma via i) enhancement of non-mature pro-IL-1β release, ii) overactivation of RIG-I inflammasome and subsequent release of mature IL-1β, and RIG-I, iii) inhibition of type I/III IFNs and ISG-responses, and iv) activation of extra proinflammatory and pro-remodelling proteins. **(Lower panel)** Pre-existing rhinovirus infection attenuates SARS-CoV-2 infection, but such coinfection augments RIG-I inflammasome activation and epithelial inflammation in patients with asthma, especially in the presence of HDM.
